# Mapping heterogeneity in the neuroanatomical correlates of depression

**DOI:** 10.1101/2025.08.27.25334497

**Authors:** Devon Watts, Travis T Mallard, Lorenza Dall’ Aglio, Evan Giangrande, Chris Kennedy, Na Cai, Karmel Choi, Tian Ge, Jordan W. Smoller

**Author notes:** Corresponding author: Devon Watts.

## Abstract

Major depressive disorder (MDD) affects millions worldwide, yet its neurobiological underpinnings remain elusive. Neuroimaging studies have yielded inconsistent results, hindered by small sample sizes and heterogeneous depression definitions. We sought to address these limitations by leveraging the UK Biobank’s extensive neuroimaging data (n=30,122) to investigate how depression phenotyping depth influences neuroanatomic profiles of MDD. We examined 256 brain structural features, obtained from T1- and diffusion-weighted brain imaging, and nine depression phenotypes, ranging from self-reported symptoms (shallow definitions) to clinical diagnoses (deep). Multivariable logistic regression, machine learning classifiers, and feature transfer approaches were used to explore correlational patterns, predictive accuracy and the transferability of important features across depression definitions. For white matter microstructure, we observed widespread fractional anisotropy decreases and mean diffusivity increases. In contrast, cortical thickness and surface area were less consistently associated across depression definitions, and demonstrated weaker associations. Machine learning classifiers showed varying performance in distinguishing depression cases from controls, with shallow phenotypes achieving similar discriminative performance (AUC=0.807) and slightly higher positive predictive value (PPV=0.655) compared to deep phenotypes (AUC=0.831, PPV=0.456), when sensitivity was standardized at 80%. However, when shallow phenotypes were downsampled to match deep phenotype case/control ratios, performance degraded substantially (AUC=0.690). Together, these results suggest that while core white-matter alterations are shared across phenotyping strategies, shallow phenotypes require approximately twice the sample size of deep phenotypes to achieve comparable classification performance, underscoring the fundamental power-specificity tradeoff in psychiatric neuroimaging research.

## Introduction

Major depressive disorder (MDD) is a leading cause of disability worldwide, affecting over 300 million people[1]. Despite decades of research[2], the neurobiological underpinnings of MDD remain poorly understood. While neuroimaging studies have attempted to elucidate the brain basis of depression (e.g., collaborative efforts from the ENIGMA consortium[3]), findings have varied considerably and often lack replication. This inability to identify robust correlates of MDD poses a significant challenge for advancing our understanding of its pathophysiology, ultimately impeding our ability to improve patient outcomes. Recent research suggests that inconsistent results may be partially explained by limited statistical power. For example, Winter and colleagues[4] demonstrated that brain structural and functional differences between individuals with MDD and healthy controls were remarkably small. This underscores the challenge of detecting subtle but important brain-behavior relationships in MDD, highlighting the need for larger sample sizes to increase precision [5, 6]. While addressing the issue of sample size will likely require large-scale, collaborative efforts over time, another major contributor to inconsistent findings that may be more readily addressed is phenotypic heterogeneity[7, 8].

Across subfields of psychiatry, differences in how depression is measured and defined not only contribute to inconsistent findings but also shape the scope of biological discoveries. In the field of psychiatric genetics, Cai and colleagues[9] demonstrated that eight different definitions of depression, ranging from “shallow” self-report measures to “deep” clinical diagnoses, yielded varying heritability and genetic correlates. Similarly, in psychiatric neuroimaging, researchers have shown that phenotypic heterogeneity complicates efforts to identify reliable neurobiological markers. For instance, Harris and colleagues[10] found that associations between depression and brain structural anatomy varied depending on whether cases were defined with self-reports, questionnaire-derived ‘probable’ depression, or diagnostic phenotyping. Although the authors expected to find greater effect sizes for more in-depth phenotypes, they found stronger associations between depression and cortical thickness when using a minimal phenotyping approach.

While these findings represent important advances, they also highlight key gaps in our understanding of depression heterogeneity. First, it is unclear how associations with imaging-derived phenotypes may vary across a broader range of depression phenotypes, particularly those defined using more granular approaches[9]. Second, the degree to which brain-based predictive models of MDD are impacted by phenotyping considerations has yet to be explored. Understanding whether the discriminatory power of imaging-derived phenotypes (IDPs) depends on the measurement of MDD – and whether models built on “shallow” definitions can generalize to “deeper” definitions and vice versa – could provide critical insights for the clinical translation of psychiatric neuroimaging.

Here, we draw upon large-scale data from the UK Biobank (UKBB) to investigate associations between 256 IDPs and nine depression phenotypes of varying depth in 30,122 adults. These phenotypes, as defined by Cai et al[9] and detailed in **Table 1**, range from minimal self-reported measures to more clinically validated diagnoses. The phenotypes are stratified into two categories: shallow phenotypes based primarily on self-reporting and help-seeking behavior, and deep phenotypes reflecting stricter or clinically confirmed definitions of depression. By characterizing associations with a diverse set of IDPs (white matter microstructure, gray matter morphology, subcortical volume, and global volume), we systematically evaluate which structural features are consistently linked to depression. To contextualize these findings, we compare our results with meta-analytic results from the ENIGMA consortium, providing insight into the robustness and specificity of neuroanatomical alterations observed for MDD relative to other forms of serious mental illness. In addition to charting neuroanatomical associations, we use advanced machine learning techniques to evaluate the predictive power of neuroimaging features and whether this depends on the depth of depression phenotyping. Finally, via a novel feature transfer approach, we assess the generalizability of predictive neuroanatomical features across depression phenotypes. Collectively, this integrative framework enables us to identify both shared and distinct neural correlates across depression phenotypes, bridging the gap between clinical heterogeneity and neurobiological findings in MDD.

**Table 1.** Spectrum of depression phenotypes in UK Biobank: from minimal to comprehensive definitions. This table presents a categorization of depression phenotypes in the UK Biobank, focusing on individuals with structural neuroimaging data. The phenotypes are arranged hierarchically, from the most minimal definitions (e.g., GPNoDep) to structured diagnostic criteria based on DSM-5 with multiple episodes (e.g., MDDRecur), building upon the work of Nai et al. (2020). Data collection in the UK Biobank involved multiple stages. Initially, all participants completed the Touchscreen Questionnaire, a broad screening tool for various health conditions and behaviors. Subsequently, a subset of participants completed the Online Mental Health Questionnaire, a more targeted assessment employing validated diagnostic instruments such as the Composite International Diagnostic Interview Short Form (CIDI-SF) to provide more precise psychiatric phenotyping. For the purposes of machine learning classification, we dichotomized these phenotypes into ‘shallow’ and ‘deep’ depression categories. Shallow depression encompasses individuals who sought help from their general practitioner (GPpsy), or psychiatrist (Psypsy), those who self-reported depression during nurse-led interviews (SelfRepDep), and those who additionally reported at least one of two cardinal symptoms of depression (low mood or anhedonia) for a minimum of two weeks (DepAll). Deep depression was defined using more stringent criteria: either electronic medical records with ICD-10 codes for depressive disorders or meeting full diagnostic criteria on the CIDI-SF administered in the online mental health questionnaire.

| Phenotype | Description | Sample Size Cases | Sample Size Controls |
| --- | --- | --- | --- |
| **GPNoDep**<br>(shallow depression) | Individuals who sought help for nerves, anxiety, tension or depression from a GP (fields 2090/2100), but did not meet criteria for depression based on the touchscreen questionnaire (fields 4631/4598). | 683 | 4789 |
| **GPPsy**<br>(shallow depression) | Individuals who sought help for nerves, anxiety, tension, or depression from a GP (fields 2090/2100). | 9529 | 20477 |
| **Psypsy**<br>(shallow depression) | Individuals who sought help for nerves, anxiety, tension, or depression from a psychiatrist (fields 2090/2100). | 2706 | 27353 |
| **SelfRepDep**<br>(shallow depression) | Individuals who self-reported having depression (field 20002) during a verbal interview with a trained nurse. | 1944 | 20421 |
| **DepAll**<br>(shallow depression) | Individuals who reported at least one of the two cardinal symptoms of depression (low mood or apathy, fields 4631/4598) for at least two weeks on the Touchscreen questionnaire and sought help from either a GP or psychiatrist. This provides a more comprehensive, though still minimal, phenotyping approach. | 1994 | 5491 |
| **ICD10Dep**<br>(deep depression) | This phenotype includes individuals with primary (data field 41202) or secondary (data field 41204) ICD-10 diagnoses of mood or affective disorders (F32-F39) from linked hospital admission records in the UK Biobank. The specific diagnoses include depressive episodes (F32), recurrent depressive disorder (F33), persistent mood disorders (F34), other mood disorders (F38), and unspecified mood disorders (F39). This definition aligns with the "ICD10-coded depression" classification used in Howard et al. 2018. | 432 | 15812 |
| **ICD10Dep_exclpsych** | This phenotype includes individuals with primary or secondary ICD-10 diagnoses of | 392 | 15432 |
| (deep depression) | mood or affective disorders (F32-F39), specifically: depressive episodes (F32), recurrent depressive disorder (F33), persistent mood disorders (F34), other mood disorders (F38), and unspecified mood disorders (F39). Both cases and controls exclude individuals with diagnoses of schizophrenia, bipolar disorder, and other psychiatric conditions. |  |  |
| **LifetimeMDD**<br>(deep depression) | Individuals who met the DSM-V criteria for lifetime Major Depressive Disorder (MDD) based on responses to the Composite International Diagnostic Interview Short Form (CIDI-SF) in the UK Biobank's follow-up online questionnaire. This uses full DSM-5 criteria for diagnosing lifetime MDD, providing a stricter phenotyping definition. | 3019 | 9458 |
| **MDDRecur**<br>(deep depression) | Individuals from the LifetimeMDD group who reported having two or more depressive episodes (field 20442), indicating recurrent MDD. | 1869 | 9105 |

## Methods

### Study Design and Participants

This cross-sectional study utilized data from the UK Biobank, (UKB; http://www.ukbiobank.ac.uk), a large population-based cohort study that recruited approximately 500,000 volunteers across the UK between 2006 and 2010. A subset of participants underwent brain MRI scans starting in 2014. The UK Biobank received ethical approval from the North West Centre Research Ethics Committee (REC number 11/NW/0382), and the current analyses were conducted under the approved UKB application (32568). We investigated the neuroanatomical correlates of depression using data from 30,122 UK Biobank Participants. Nine depression phenotypes of varying depth were examined, as previously described[9]. These phenotypes ranged from “shallow” definitions based on minimal criteria to “deeper” definitions meeting Major Depressive Disorder (MDD) thresholds according to a structured self-report (CIDI-SF) or ICD-10 diagnoses from EMR. The essential characteristics of these phenotypes, taken from Cai et al. 2020 [9], and respective sample sizes of cases and controls across depression phenotypes can be found in **Table 1**, whereas a visual depiction can be found in **Supplementary Figure S2**. For machine learning analyses, we dichotomized “shallower” vs. “deeper” definitions, excluding the GPNoDep category. The deeper phenotypes were based on meeting MDD thresholds according to either the CIDI-SF or ICD-10 diagnoses, while the shallower phenotypes encompassed GPpsy, Psypsy, SelfRepDep, and DepAll. Although GPNoDep was excluded from machine learning analyses as these individuals did not meet criteria for depression, we retained it in other analyses to provide insights into the spectrum of mood-related help-seeking behaviors and their potential neurobiological correlates. Image acquisition and processing protocols for brain MRI data, including T1-weighted structural and diffusion-tensor imaging procedures are detailed in the Supplementary Methods. Briefly, we analyzed 256 brain imaging features: 68 cortical thickness, 68 surface area, 10 global cortical volumes, 14 subcortical volumes, 48 fractional anisotropy, and 48 mean diffusivity measurements. Population stratification control procedures using genetic ancestry principal components[11], visualization methods for mapping neuroanatomical correlates across depression phenotypes using 3D brain maps, and approaches for conducting pairwise comparisons of effect sizes between phenotypes are also described in the Supplementary Methods.

### Feature preprocessing and normalization

Prior to conducting univariate logistic regression models, all 256 candidate neuroimaging features were z-score normalized to ensure comparability across different measurement scales and to facilitate interpretation of the resulting odds ratios (ORs). Z-score normalization[12] was performed by subtracting the mean and dividing by the standard deviation of each feature, resulting in a standardized scale with a mean of 0 and a standard deviation of 1. ORs were calculated by exponentiating the coefficients of the logistic regression models, representing the change in the odds of depression associated with one standard deviation change in the brain measure, adjusting for confounders.

### Univariate and multivariable logistic regression analyses

Univariate logistic regression analyses were conducted using the *’glm’* function in R within the Stats package[13], with the presence or absence of depression as the outcome variable and neuroimaging features as predictor variables. Multivariable logistic regression models were adjusted for several covariates, including age, sex, assessment center, estimated total intracranial volume, head motion, and the top 20 genetic ancestry PCs to control for potential confounding due to population stratification. To calculate 95% confidence intervals (CIs) for the ORs, we used the profile likelihood method. Unlike Wald intervals, which assume that the uncertainty in parameter estimates follow a symmetric, bell-shaped distribution around the best estimate[14], profile likelihood intervals allow for asymmetry in this uncertainty[15]. This flexibility makes profile likelihood intervals more reliable when the true shape of the uncertainty distribution deviates from this ideal symmetric form. To evaluate overlap in case definitions across depression phenotypes, we computed pairwise Jaccard indices[16] as shown in **Supplementary Figure S13** (range: 0–1; higher values indicate greater overlap). These analyses revealed minimal overlap between shallow and deep depression phenotypes (e.g., GPNoDep vs. ICD10Dep: J = 0.014; DepAll vs. LifetimeMDD: J = 0.126), supporting their stratification into distinct analytical groups within ML analyses.

### UK Biobank and ENIGMA consortium effect size comparisons

To position our findings within the broader neuroimaging literature, we compared UK Biobank effect sizes with ENIGMA consortium meta-analytic estimates for major depressive disorder, bipolar disorder (BD), and schizophrenia (SCZ). We converted odds ratios from our analyses to Cohen’s d values [17] using the formula d = ln(OR) × √3 / π, and calculated Spearman rank correlations [18] between UK Biobank and ENIGMA effect sizes across imaging measures. Additionally, we conducted comparative analyses of the top 5 effect sizes between datasets, restricting analyses to white matter tracts common to both studies and averaging hemispheric values to align with ENIGMA’s bilateral reporting format. Complete methodological details for these effect size comparisons are provided in Supplementary Methods.

### Brain-based Prediction Models

To complement our examination of brain-based associations across depression phenotypes using case-control binary logistic regression analyses, we developed a series of machine learning classifiers to investigate the predictive power and generalizability of these associations. Our aim was to determine whether neuroimaging features could accurately discriminate between different depression phenotypes and controls, and to assess how both the most important predictive features and the model performance metrics varied across phenotypes. To achieve this, we implemented a range of binary and multi-class classification models to explore various comparisons: deep depression versus controls, shallow depression versus controls, combined depression versus controls, deep versus shallow depression, and a multi-class classifier distinguishing between deep depression, shallow depression, and controls. This approach allowed us to evaluate how predictive performance metrics (such as AUC-ROC, sensitivity, and specificity) and feature importance rankings varied across different depression phenotypes and classification tasks. By examining both the performance metrics and the most important features in each model, we sought to understand which neuroanatomical characteristics were most predictive of different depression phenotypes and how generalizable these patterns were across the phenotyping spectrum.

### Model development and implementation

To implement our classification approach, we primarily utilized random forest classifiers, leveraging the *mlr3[19]* and *ranger[20]* packages in R. Random forests were chosen for their ability to handle high-dimensional data, capture nonlinear relationships, and provide interpretable feature importance measures[21]. As a point of comparison and to assess the potential benefits of more complex modeling strategies, we also implemented stacked ensemble models, as further discussed in the **Supplementary Methods**. Briefly, stacked ensembles, also known as super learners, combine predictions from multiple base models to create a more robust meta-model with potential improvements in performance metrics[22]. Our ensemble consisted of five diverse base learners, including random forest, elastic net[23], *k*-nearest neighbors (kNN)[24], light gradient-boosting machine (lightGBM), and naïve bayes[25]. Random forest, already utilized as our primary classifier, excels in capturing non-linear relationships and providing feature importance measures[21]. LightGBM was incorporated for its ability to handle large-scale, high-dimensional data and capture nonlinear interactions through gradient boosting. Elastic net, a form of logistic regression combining L1 and L2 regularization, was chosen for its capacity to perform feature selection and handle multicollinearity, respectively, within a linear framework[23]. The k-nearest neighbors (kNN) algorithm was incorporated for its ability to model complex decision boundaries without making distributional assumptions, particularly useful for capturing local patterns in the data[24]. Lastly, naive bayes was included for its efficiency in high-dimensional spaces and effectiveness with smaller training sets, despite its assumption of feature independence[25]. A key feature of our stacked ensemble architecture was using cross-validated predictions from the base learners as input features for the meta-learner, rather than the original features[26]. This approach, implemented through a 5-fold cross-validation pipeline, allows the ensemble to potentially capture a wider range of probability distributions and decision boundaries than any individual base learner alone.

Features were z-score normalized prior to model development. To evaluate the impact of potential confounding factors on model performance, residualization was performed, where for each neuroimaging feature, we regressed out the effects of age, sex, assessment center, estimated total intracranial volume, head motion, and the top 20 genetic ancestry principal components using a linear model with the lm() function in R[13]. This process was performed separately for shallow and deep depression phenotypes. We implemented random forest classifiers using a 70-30 stratified split (*rsample[27]* package) to preserve outcome class distributions between training and testing sets. Hyperparameter tuning was performed using Bayesian optimization[28] via the *mlr3mbo* package with 5-fold cross-validation with 5 repeats, balancing exploration of the hyperparameter space against computational efficiency[29]. To mitigate class imbalance[30], we implemented inverse frequency class weighting rather than data resampling techniques. Model performance was assessed on the 30% holdout using AUC-ROC (primary metric), F1 score (harmonic mean of precision/recall), sensitivity, specificity, PPV, NPV, and PR-AUC (95% CIs via stratified bootstrapping). Sensitivity thresholds were standardized to 80% for cross-model comparison, with depression cases designated as the positive class in case-control models and deep depression cases in deep vs shallow comparisons, respectively. Detailed preprocessing procedures, hyperparameter optimization, model evaluation, and feature importance calculations are provided in Supplementary Methods.

### Feature transfer machine learning approach

To investigate the specificity and generalizability of brain-based associations across depression phenotypes, we implemented a novel feature transfer approach. We first trained separate random forest classifiers for deep vs controls and shallow vs controls classification tasks. Using recursive feature elimination[31] to rank features by importance, we identified the top 30% of features (76 features) from each model based on their predictive contribution to the respective classification task. We then trained two sets of models for comparison. Outcome-specific models used the top 30% of features derived from the same classification task, serving as our baseline comparison. Feature transfer models used the top 30% of features derived from the opposite classification task. Specifically, deep vs controls classifiers were trained using the 76 most important features identified by the shallow vs controls classifier, and shallow vs controls classifiers were trained using the 76 most important features identified by the deep vs control classifier, respectively. Model performance metrics, including AUC, sensitivity, specificity, PPV, NPV, and AUPRC were compared between outcome-specific features and feature transfer approaches This systematic comparison allowed for evaluating whether neuroanatomical features predictive of one depression phenotype were specific to deep or shallow definitions or exhibited generalizability across phenotyping depth.

### Model comparison and statistical testing

Furthermore, given the possibility of overlapping confidence intervals in performance metrics across outcome-specific and feature transfer classifiers, stratified bootstrap tests with 10,000 iterations for F1 score and AUC metrics were used to ascertain whether there were statistically significant differences between models. The stratified bootstrap test involves repeatedly sampling the data with replacement, stratifying by class to maintain the original class distribution[32]. The stratification was implemented by first calculating the proportion of each class in the original data, then separately resampling from each class with replacement to preserve these proportions in every bootstrap iteration. This approach ensures consistent class distributions across all bootstrap samples, reducing variability that could otherwise arise from random sampling in imbalance datasets. We implemented a two-tailed test by calculating p-values based on the proportion of bootstrap samples where the absolute difference in performance metrics (F1 score or AUC) between the two classifiers was greater than or equal to the observed difference. This approach accounts for performance differences favoring either classifier. Importantly, we performed simultaneous bootstrapping for both classifiers, calculating the difference in each iteration. This method preserves the paired nature of the observations and models the sampling distribution of the performance differences directly. We selected F1 score and AUC as informative metrics because the F1 score provides a balanced measure of precision and recall, while AUC evaluates the model’s ability to discriminate between classes across various threshold settings. Additionally, we employed McNemar’s test to compare the classification performance of the models. This test assesses the differences between two models’ classification errors on the same dataset, considering the paired nature of the data[33].

### Multi-class classification

Additionally, to capture the nuanced relationships between different depression phenotypes and healthy controls simultaneously, we implemented multi-class random forest classifiers. This approach allowed us to discriminate between shallow depression, deep depression, and healthy controls within a single model, potentially revealing patterns that may be obscured in binary classifications. We utilized the *mlr3[19]* and *ranger[20]* packages to maintain consistency with our binary classification methodology. Hyperparameter tuning was performed using Bayesian optimization with the *mlr3tuning* package, optimizing for multi-class AUC (MAUC) using the “classif.mauc_aunu” measure, which extends binary AUC to multi-class problems and provides a balanced assessment of the model’s discriminative ability across all classes[34]. We applied the same data splitting, stratified sampling, feature residualization, and feature importance strategies used in our binary classifiers to ensure comparability. To account for class imbalance in the multi-class setting, we weighted performance metrics by class prevalence.

### Code Availability

A static version of all custom code used for data analysis and visualization in this study is available upon request. This includes R scripts for logistic regression analyses, visualization of neuroanatomical correlates, pairwise comparisons of effect sizes across depression phenotypes, concordance analyses with meta-analytic estimates, and machine learning analyses. For the visualization of neuroanatomical correlates, we utilized the *ggseg3d* package, which allows for the creation of interactive 3D brain maps of effect sizes. This package is openly available and can be accessed at https://github.com/ggseg/ggseg3d.

## Results

### 1. White matter microstructure shows consistent alterations across depression phenotypes

We investigated the neuroanatomical correlates of depression across various phenotyping strategies with a series of multivariable logistic regression models, each including one of 256 z-score transformed brain structural variables as the primary predictor, alongside covariates (age, sex, intracranial volume, assessment center, and head motion). We derived odds ratios (ORs) for depression in association with brain white matter microstructure (fractional anisotropy and mean diffusivity at major fiber bundles), gray matter morphometry (cortical thickness and surface area), subcortical volumes, and global volumetric measures. All p-values reported were adjusted for multiple comparisons using the False Discovery Rate (FDR) method across all 256 tests. For detailed information on each variable, refer to **Supplementary Table S1**. Depression operationalization across phenotypes is described in **Table 1** and in the Methods section.

All brain structural correlates across IDPs and depression phenotypes are summarized in **Supplementary Table S2**. To illustrate these associations, we focused on specific examples comparing shallow (e.g. GPPsy and SelfRepDep) and deep (e.g. LifetimeMDD and MDDRecur) phenotypes as defined by Cai and colleagues[9], highlighting their most robust associations and the number of significant regions identified. As shown in **Supplementary Table S2**, white matter microstructure in fractional anisotropy and mean diffusivity demonstrated the most pronounced associations with depression status across phenotypes. The spatial distribution of associations across regions for each IDP is shown in **Supplementary Figure S1**.

Fractional anisotropy measures predominantly showed reduced values across depression phenotypes, with 85.88% of odds ratios below 1.0. Values ranged from 0.781 to 1.098 (median: 0.949, mean: 0.946, SD: 0.052). The most robust associations in fractional anisotropy were observed in major white matter bundles, including the corpus callosum, corona radiata, and principal association fibers. For instance, in the GPpsy phenotype, fractional anisotropy was significantly lower in 31 white matter tracts (FDR-adjusted p ≤ 0.05) out of the 48 examined, with the strongest association in the superior fronto-occipital fasciculus (right; OR = 0.915, 95% CI: 0.891-0.939, FDR-adjusted p = 6.65e-09). By comparison, the MDDRecur phenotype showed fractional anisotropy reductions in 11 white matter tracts (FDR-adjusted p ≤ 0.05). Complementing the fractional anisotropy findings, mean diffusivity showed widespread increases associated with depression across phenotypes, with 75.23% of the odds ratios above 1, indicating higher diffusivity in depression cases, compared to controls. Mean diffusivity measures also demonstrated a broader range of associations compared to fractional anisotropy, with odds ratios spanning 0.870 to 1.282 (median: 1.032, mean: 1.042, SD: 0.066). The SelfRepDep phenotype exhibited significantly higher mean diffusivity in 36 white matter tracts (FDR-adjusted p ≤ 0.05) out of the 48 examined, while MDDRecur demonstrated significantly higher mean diffusivity in 15 white matter tracts. White matter associations for both fractional anisotropy and mean diffusivity across phenotypes are illustrated in **Figure 1**, with individual odds ratios, 95% confidence intervals, and uncorrected and FDR-adjusted p-values across all imaging-derived phenotypes shown in **Supplementary Table S2**.

**Figure 1.**
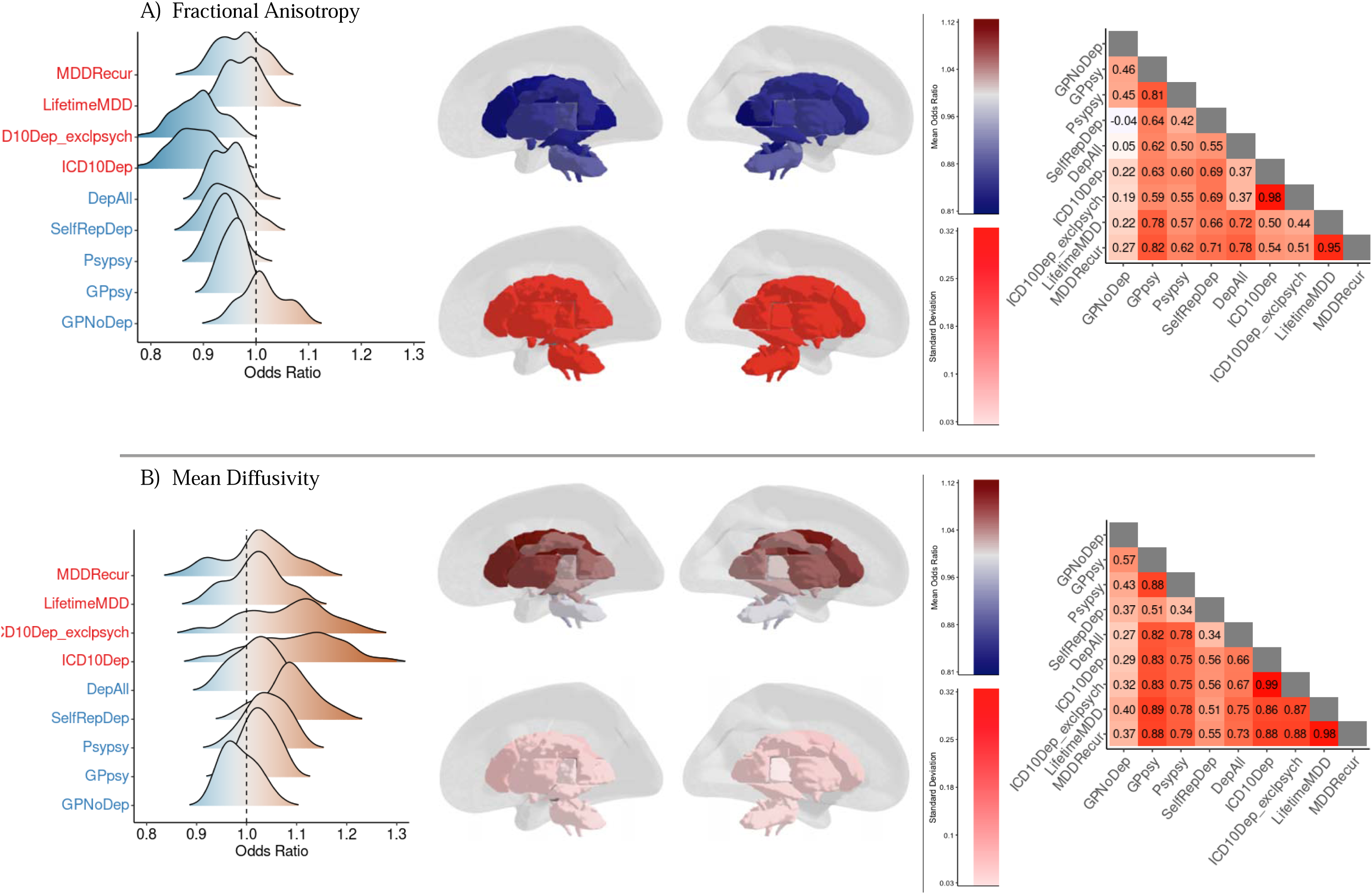
Associations between white matter imaging-derived phenotypes and depression definitions. This figure illustrates the relationship between white matter microstructure and depression across varying phenotyping strategies. These results are derived from multivariable logistic regression models, where each included one of 256 z-score transformed neuroanatomical variables as the primary predictor, along with covariates (age, sex, intracranial volume, assessment center, and head motion), with depression status (case/control) as the outcome. The neuroanatomical variables compass white matter microstructure (FA and MD), gray matter morphometry (cortical thickness and surface area), subcortical volumes, and global brain volumetric measures. **(A)** Fractional anisotropy (FA) and **(B)** mean diffusivity (MD) associations are presented through density plots, 3D brain maps, and correlation heatmaps. Density plots (left) display odds ratios (ORs) for FA and MD across depression phenotypes, ranging from strictly defined (top) to minimally defined (bottom). Deeper phenotypes are depicted in red, shallower in blue. ORs < 1 are shown in light blue, >1 in light brown. The 3D brain maps (center) visualize mean ORs (top) and standard deviations (bottom) across all nine depression definitions. For FA, cooler colors (blues) indicate regions where increased FA is associated with decreased odds of depression. For MD, warmer colors (red) show areas where increased MD correlates with higher depression odds. Standard deviation maps highlight regions with the most variance in effects across definitions. Heatmaps (right) display Spearman’s correlations of ORs across depression definitions, ranging from - 1 (perfect negative correlation) to 1 (perfect positive correlation). These heatmaps provide insight into the consistency of neuroanatomical associations across different depression phenotypes, with darker colors indicating stronger correlations. FA shows widespread reductions (ORs < 1) across white matter tracts, indicating that lower FA values are generally associated with higher odds of depression. The strongest associations are observed in the corpus callosum, corona radiata, and major association fibers. Concurrently, MD demonstrates widespread increases (ORs > 1), with the most pronounced effect in similar regions, particularly the corpus callosum, corona radiata, and internal capsule. Importantly, the variability in effects across phenotyping definitions differs between FA and MD. FA shows higher variability (standard deviations of ORs ranging from ∼0.29 to ∼0.32) compared to MD (standard deviations of ORs ranging from ∼0.03 to ∼0.09). This suggests that FA measures may be more sensitive to the depth and specificity of depression phenotyping strategies, while MD associations appear more consistent across different phenotyping approaches. These patterns suggest that altered white matter microstructure, characterized by both reduced FA and increased MD, is a pervasive feature across depression phenotypes, but with differing levels of consistency.

**Figure 2.**
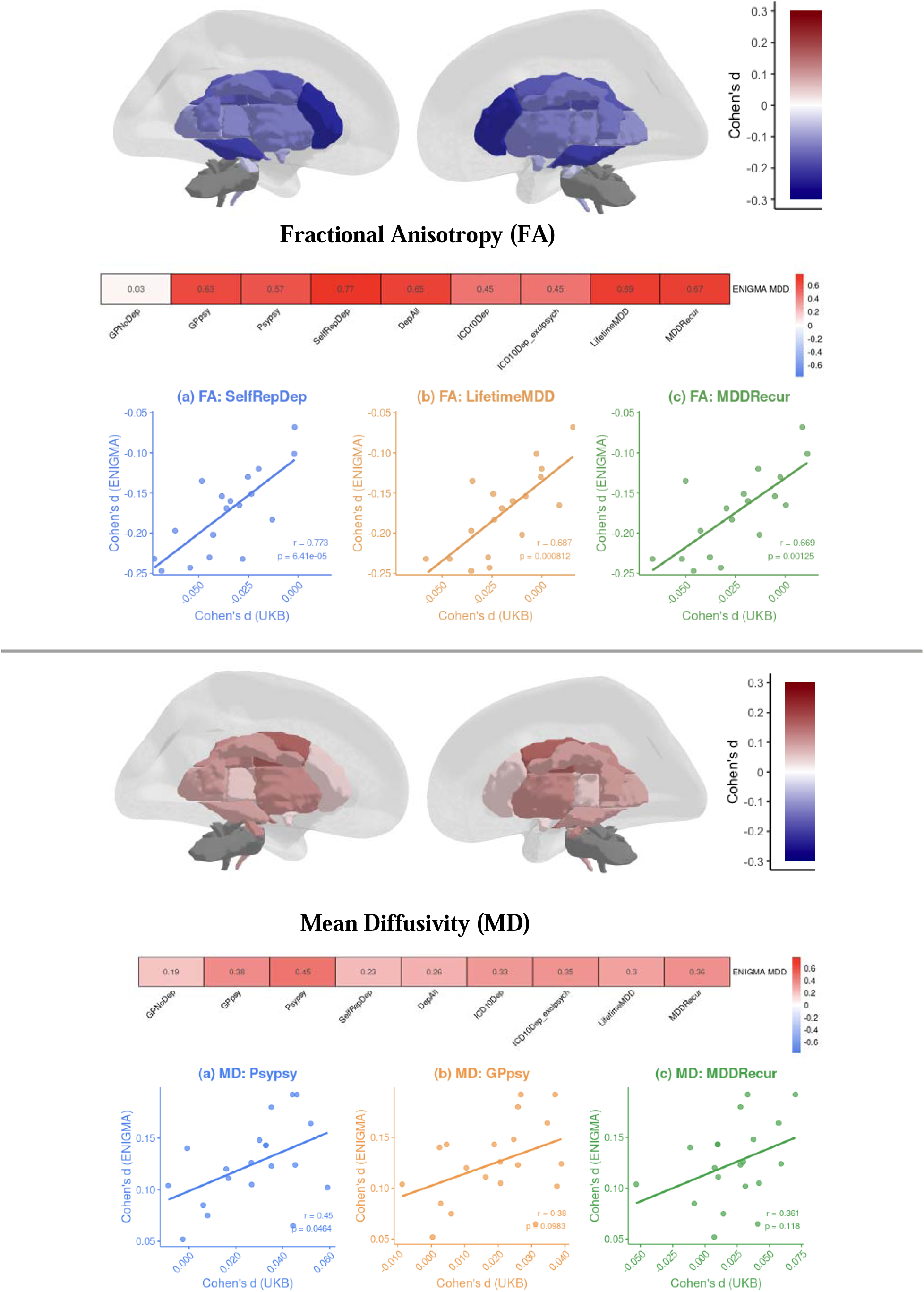
Relationships between brain-psychopathology associations in UK Biobank and the ENIGMA MDD consortium in white-matter tracts. This figure illustrates the comparison of neuroimaging effect sizes for white matter diffusion metrics between the UK Biobank (UKBB) and effect size estimates from the latest and largest meta-analysis from the ENIGMA MDD Working Group. The figure is divided into two main sections: (A) Fractional Anisotropy (FA) and Mean Diffusivity (MD). Each section comprises three panels. The upper panels present 3D brain maps illustrating ENIGMA MDD Working Group effect size estimates (Cohen’s d). For FA, cooler colors (blues) indicate regions where increased FA is associated with decreased odds of depression, while for MD, warmer colors (red) show areas where increased MD correlates with higher depression odds. The middle panels display heatmap vectors of Spearman’s rank correlations between ENIGMA MDD effect size estimates and UKBB results across various depression phenotypes. For FA, stronger correlations are generally observed for more ‘strictly’ defined phenotypes such as LifetimeMDD and MDDRecur. In contrast, among MD measures, weaker correlations were observed between effect size estimates and UKBB results relative to FA, with the strongest positive correlations observed among shallower phenotypes, including Psypsy and GPpsy. The lower panels feature scatter plots for the three depression phenotypes showing the strongest correlations with ENIGMA MDD effect size estimates. These plots compare Cohen’s d values from ENIGMA MDD (vertical axis) against those from UKBB (horizontal axis) across white matter tracts. For FA, the phenotypes displayed are SelfRepDep (blue, r = 0.773), LifetimeMDD (orange, r = 0.687), and MDDRecur (green, r = 0.669), demonstrating strong positive correlations across these phenotypes. For MD, the phenotypes are Psypsy (blue, r = 0.45), GPpsy (orange, r = 0.38), and MDDRecur (green, r = 0.361), showing moderate positive correlations. Each data point represents a specific white matter tract.

In contrast to the widespread associations for white matter microstructure, cortical thickness and surface area measures generally showed few significant associations with depression (**Supplementary Figure S1, Supplementary Figure S3, Supplementary Table S2)**. Cortical thickness odds ratios (ORs) ranged from 0.862 to 1.169 (median = 0.990, mean = 0.994, SD = 0.008), whereas surface area measures displayed a similarly narrow range (ORs 0.824–1.133; median = 0.981, mean = 0.991, SD = 0.019). Across depression phenotypes, significant associations (FDR-adjusted p ≤ 0.05) in surface area were limited to GPpsy (4 regions), Psypsy (2 regions), and ICD10Dep (1 region). Overall, MDDRecur exhibited the largest number of significant thickness associations (6 regions), followed by GPpsy, SelfRepDep, and LifetimeMDD (4 regions each), ICD10Dep (2 regions), and Psypsy and ICD10DepExclude (1 region each), with DepAll showing no significant cortical findings. In sum, across phenotyping approaches, fractional anisotropy and mean diffusivity showed stronger and more consistent associations with depression, whereas cortical thickness and surface area yielded fewer and more variable results.

#### 1.1. Self-reported depression shows stronger white-matter microstructural alterations than clinical definitions

To assess whether the strength of associations varied across phenotypic definitions, we conducted pairwise comparisons of odds ratios using the delta method. These comparisons revealed that the SelfRepDep phenotype yielded significantly stronger associations (i.e., larger odds ratios) with several mean diffusivity measures than the clinically-defined phenotypes. No significant differences in odds ratios between phenotypes were found for fractional anisotropy, cortical thickness, or surface area measures after multiple comparison correction (see Supplementary Results for detailed comparisons). These findings suggest that while white matter microstructure is more consistently implicated in depression than cortical morphology, the magnitude of this association can vary significantly depending on the phenotype used.

### 2. Comparison of Findings Between the UKB and ENIGMA Consortium

To contextualize our findings within the broader neuroimaging literature, we compared effect sizes from UK Biobank data with meta-analytic estimates from ENIGMA consortium working groups for MDD[35, 36], BD [37, 38], and SCZ [39, 40]. We calculated Spearman rank correlations between UKB Cohen’s d values (converted from odds ratios) and corresponding ENIGMA meta-analytic Cohen’s d estimates across brain regions for each neuroimaging measure (cortical thickness, surface area, fractional anisotropy, and mean diffusivity) and depression phenotype definition. None of the correlations between UKB and ENIGMA effect sizes were statistically significant for cortical measures (thickness: r = −0.01 to 0.31, p = 0.100-0.910; surface area: r = −0.08 to 0.22, p = 0.477-0.739). In contrast, greater concordance was observed for white matter microstructure measures, particularly fractional anisotropy, with the stronger correlations found in deeper phenotypes such as LifetimeMDD (r = 0.69), MDDRecur (r = 0.67), and the intermediary phenotype DepAll (r = 0.65).

We then compared Cohen’s d values for the five tracts with the largest reported effect sizes in ENIGMA MDD with their corresponding UKBB Cohen’s d values for those same tracts, and reciprocally examined the five tracts with the largest UKBB Cohen’s d values in ENIGMA MDD **(Figure 3)**. While UKBB effect sizes were consistently smaller than ENIGMA meta-analytic estimates, we observed significant effects in EMR-based depression definitions (ICD10Dep and ICD10Dep_exclpsych). Specifically, in ICD10Dep, we observed significant reductions (FDR-adjusted p < 0.05) in the genu of corpus callosum (d = −0.14), body of corpus callosum (d = −0.12), and bilateral anterior limb of internal capsule (d = −0.09), with similar magnitudes in ICD10Dep_exclpsych. Together, while these results demonstrate directional agreement with ENIGMA, the overall magnitudes of effect sizes in UKBB were smaller, which may be partly driven by sample sizes and demographic differences between UKBB and ENIGMA, rather than phenotyping depth alone. Comparisons of UKBB results with meta-analytic effect size estimates from ENIGMA BD and ENIGMA SCZ can be found in **Supplementary Figures S4-S7**. Complete details of these cross-consortium comparisons are provided in Supplementary Methods and Supplementary Results.

**Figure 3.**
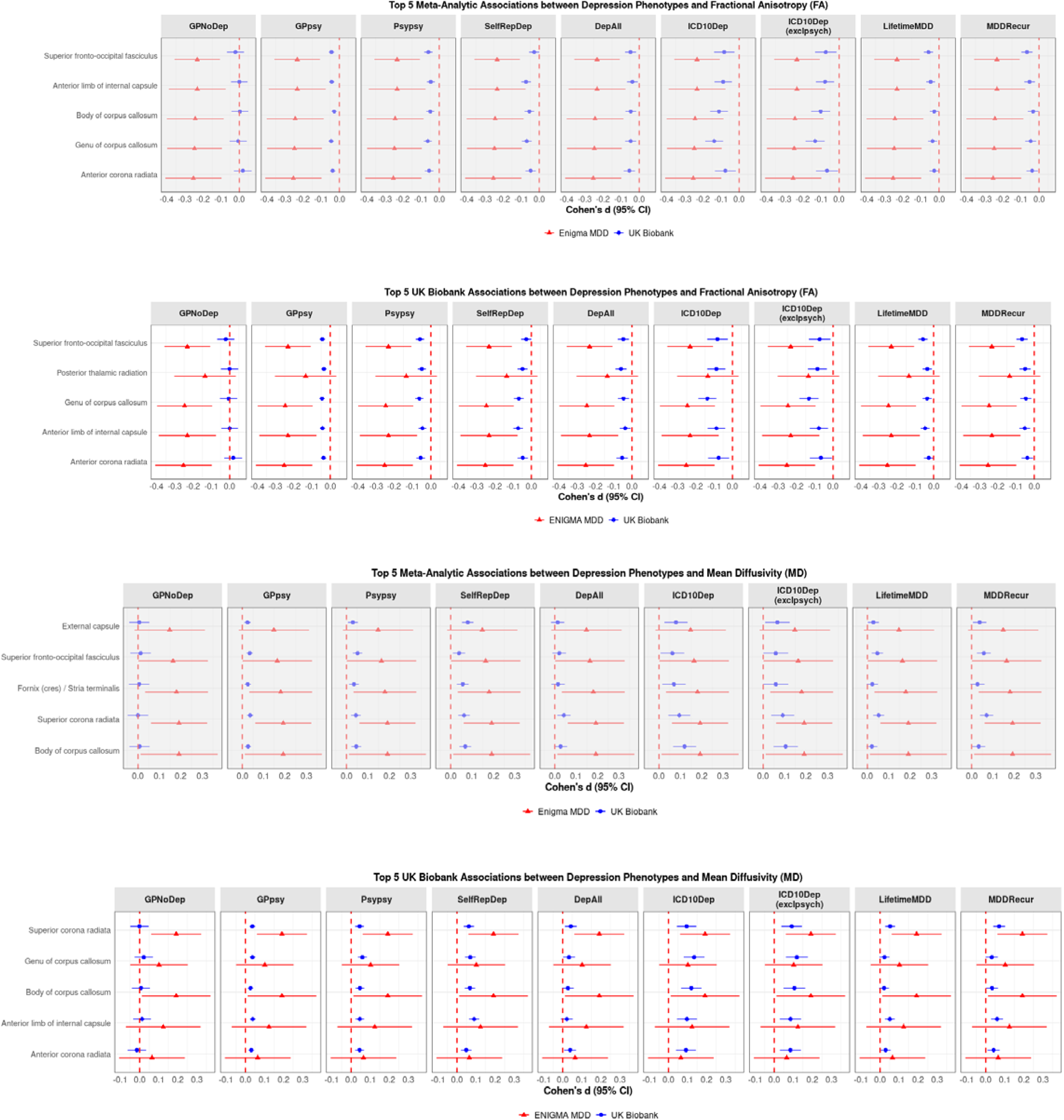
Comparison of effect sizes between UK Biobank depression phenotypes and ENIGMA MDD meta-analysis for fractional anisotropy (FA) and mean diffusivity (MD) measures. This figure presents a comparison of effect sizes (Cohen’s d) for white matter diffusion metrics between various depression phenotypes in the UK Biobank (UKBB) and the largest and most recent meta-analytic findings from the ENIGMA Major Depressive Disorder (MDD) Working Group. The first panel presents the five strongest associations between depression and FA from ENIGMA MDD, juxtaposed with corresponding effect sizes across nine depression phenotypes in the UK Biobank. The meta-analytic effect size estimates demonstrate small to moderate reductions in fractional anisotropy in individuals with depression, with Cohen’s d values ranging from −0.2 to −0.4. These reductions are observed in key white matter tracts, including the superior fronto-occipital fasciculus, anterior limb of internal capsule, body and genu of corpus callosum, and anterior corona radiata. In contrast, the UKBB data reveal substantially smaller effect sizes for most phenotypes among these tracts, clustering around zero. Notably, depression definitions based on electronic medical records (EMR) show slightly larger effect sizes, though still truncated relative to those reported in the meta-analysis. The second panel displays the five strongest associations between depression and fractional anisotropy in the UKBB, compared with their corresponding effect sizes in ENIGMA MDD. The UKBB effect sizes are generally small, with Cohen’s d values close to zero across most phenotypes. The ENIGMA MDD effect size estimates for these regions, while also small, are negative across all five regions, indicating subtle reductions in fractional anisotropy associated with depression. The third panel, mirroring the first, presents the five strongest associations between depression and mean diffusivity from the ENIGMA meta-analysis, alongside the corresponding UKBB phenotypes. The ENIGMA effect sizes range from 0.1 to 0.2, indicating small increases in mean diffusivity in regions such as the external capsule, superior fronto-occipital fasciculus, fornix/stria terminalis, superior corona radiata, and body of corpus callosum. As with fractional anisotropy, the UK Biobank effect sizes are smaller, with depression definitions based on EMR showing the closest alignment with the ENIGMA results. The fourth panel illustrates the five strongest associations between depression and mean diffusivity in the UK Biobank, compared with their corresponding effect size estimates in ENIGMA MDD. The UK Biobank effect sizes vary across phenotypes but are generally small. The ENIGMA MDD effect sizes for these regions are consistently positive but small, suggesting subtle increases in mean diffusivity associated with depression. Across all panels, a consistent pattern emerges where effect sizes in the UK Biobank are generally smaller and more variable across phenotypes compared to the ENIGMA Major Depressive Disorder meta-analysis. Depression phenotypes defined using EMR in the UK Biobank show the closest alignment with the ENIGMA results, particularly for fractional anisotropy measures.

### 3. Machine learning classification of depression phenotypes

For the purposes of our machine learning classification, we categorized eight UK Biobank depression phenotypes into two broader groups (“shallow” versus “deep”), as summarized in **Table 1**. GPNoDep was excluded from machine learning analyses as these individuals sought help for mental health concerns but did not meet depression criteria. Full methodological details are provided in Methods and Supplementary Methods.

#### 3.1.1. Neuroanatomical correlates yield divergent classification metrics across depression phenotyping depth

We developed multiple machine learning classifiers to discriminate between depression cases and controls using 256 IDPs. Our primary analysis compared deep versus shallow depression classifications, while additional analyses explored multi-class classification and stacked ensemble methods. When distinguishing cases from controls in the testing set (30%), shown within **Table 2**, the deep depression classifier achieved an area under the receiver operating characteristic curve (AUC) of 0.831 (95% CI: 0.819-0.844), while the shallow depression (broad) model showed a slightly lower AUC of 0.810 (95% CI: 0.802-0.819). At a standardized sensitivity of 80%, the deep depression model demonstrated a specificity of 68.2% (95% CI: 66.9%-69.4%), compared to 61.1% (95% CI: 59.7%-62.5%) for the shallow depression (broad) model. The positive predictive value (PPV) for the deep depression model was 45.6% (95% CI: 43.8%-47.4%), while the shallow depression (broad) model achieved a substantially higher PPV of 67.2% (95% CI: 66.0%-68.4%). Both models demonstrated high negative predictive values (NPV), with 91.1% (95% CI: 90.1%-91.9%) for deep depression and 75.4% (95% CI: 74.0%-76.8%) for shallow depression (broad). The shallow (broad) model also showed superior performance in F1 score (0.730 vs 0.581) and AUPRC (0.849 vs 0.716), indicating better precision-recall balance.

**Table 2.** Performance metrics of random forest classifiers for deep and shallow depression phenotypes. Random forest classifiers were trained on 256 neuroimaging features to discriminate deep depression (defined by DSM-V criteria for MDD or ICD-10 diagnoses) vs controls and shallow depression (based on self-reported symptoms) vs. controls. Models were developed using stratified sampling (70% training, 30% testing) with Bayesian optimization for hyperparameter tuning. In our depression phenotyping approach, individuals could meet multiple operational definitions across the spectrum. For the shallow vs control classifiers, we distinguished between broad and narrow definitions: the broad definition includes individuals who meet criteria for shallow depression regardless of whether they also meet criteria for deeper phenotypes, capturing the full spectrum as typically defined in population cohorts; the narrow definition includes only individuals who exclusively meet criteria for shallow depression without meeting deeper phenotype criteria. This broad approach reflects how depression is typically defined in large health systems and population cohorts, capturing the full spectrum of self-reported depression. For the deep vs controls model, the training set consisted of 11,978 controls and 3998 deep depression cases, with a test set of 5134 controls and 1714 deep depression cases. For the shallow (broad) versus controls classifier, the training set comprised 11,978 controls and 11,321 shallow depression cases, with a test set of 5134 controls and 4852 shallow depression cases. The shallow (narrow) versus controls classifier used a training set of 11,978 controls and 7996 shallow depression cases, with a test set of 5134 controls and 1714 shallow depression cases. Features were residualized for potential confounders including age, sex, assessment center, estimated total intracranial volume, head motion, and the top 20 genetic ancestry principal components. This residualization was performed using linear regression models for each feature, with the confounders as predictors. The residuals from these models were then used as input features for the classifiers. Sensitivity was standardized to 80% for all models to facilitate direct comparisons. Performance metrics are reported with 95% confidence intervals. As summarized in Table 2, the deep depression model showed a slightly higher AUC (0.831 vs 0.807), while the shallow depression (broad) classifier demonstrated stronger performance across several other key classification metrics, including a higher F1 score (0.720 vs 0.581) and PPV (0.655 vs 0.456) compared to the deep phenotype classifier. These differences reflect a more favorable balance between precision and recall for the shallow (broad) model at the standardized sensitivity of approximately 80%. The higher PPV indicates that when the shallow depression (broad) model predicts a positive case, it is correct 65.5% of the time, compared to 45.6% for the deep depression model, representing a lower false positive rate. Additionally, the shallow (broad) model achieved a higher AUPRC (0.849 vs 0.716), indicating better precision across different recall levels despite its slightly lower AUC. The shallow (narrow) model showed intermediate performance on most metrics (F1 score: 0.635, PPV: 0.526, AUC: 0.782, AUPRC: 0.792), suggesting that including individuals who also meet deep phenotype criteria enhances the discriminative ability of shallow depression classification.

|  | Deep vs Controls | Shallow vs Controls (Broad) | Shallow vs Controls (Narrow) |
| --- | --- | --- | --- |
| **F1 Score** | 0.581 (0.564, 0.597) | 0.720 (0.711, 0.729) | 0.635 (0.623, 0.646) |
| **Sensitivity** | 0.799 (0.780, 0.818) | 0.800 (0.788, 0.811) | 0.800 (0.786, 0.813) |
| **Specificity** | 0.682 (0.669, 0.694) | 0.655 (0.643, 0.667) | 0.519 (0.505, 0.533) |
| **PPV** | 0.456 (0.438, 0.474) | 0.655 (0.643, 0.667) | 0.526 (0.512, 0.540) |
| **NPV** | 0.911 (0.901, 0.919) | 0.761 (0.748, 0.774) | 0.795 (0.781 - 0.809) |
| **AUC** | 0.831 (0.819, 0.844) | 0.807 (0.799, 0.816) | 0.782 (0.772, 0.793) |
| **AUPRC** | 0.716 (0.696, 0.735) | 0.841 (0.832, 0.849) | 0.792 (0.781, 0.802) |

To further investigate neuroanatomical distinctions between depression phenotypes, we implemented multi-class random forest and Super Learner classifiers using all 256 candidate features. As shown in **Supplementary Table S3**, the multi-class random forest classifier achieved a macro average AUC of 0.803 (95% CI: 0.795-0.811), with deep depression showing the highest class-specific AUC of 0.841 (95% CI: 0.829-0.853), followed by shallow depression (AUC = 0.797, 95% CI: 0.787-0.806) and controls (AUC = 0.771, 95% CI: 0.762-0.780). Unlike the random forest models in **Table 2**, our super learner classifiers (**Supplementary Table S4**) using all 256 candidate features demonstrated superior performance for deep vs control classification (AUC = 0.911, 95% CI: 0.899 − 0.923) compared to shallow vs control (AUC = 0.765, 95% CI: 0.752-0.775). Separate random forest classifiers were also developed for individual depression phenotypes, as detailed in **Supplementary Tables S5-S8**. **Supplementary Figure S8** presents feature rankings for shallow and deep depression models using all 256 neuroimaging features, while **Supplementary Figure S9** examines feature importance patterns in multi-class versus binary case-case classifications (shallow versus deep). Feature importance analysis revealed that deep depression models relied more heavily on global volumetric measures (e.g., normalized gray matter volume and peripheral cortical gray matter volume), while shallow models placed greater emphasis on surface area measurements, though some features such as mean fractional anisotropy in the fornix maintained consistent importance across both phenotypes. Notably, volume-based measurements (peripheral cortical gray matter, total gray matter, and ventricular cerebrospinal fluid volumes) were most important for distinguishing depression cases from controls in the multi-class classifier, whereas differentiation between depression phenotype depths relied on a broader range of neuroanatomical features. In sum, the larger and less stringent depression definitions were easier to predict with neuroimaging features, while deeper definitions showed lower PPV across models, implying trade-off between clinical specificity and statistical power.

#### 3.1.2. Neuroanatomical features derived from broader depression phenotypes generalize well to more stringent depression definitions

To investigate the specificity and generalizability of neuroanatomical predictors across depression phenotypes, we implemented a feature transfer approach (**Methods; Supplementary Figures S10-S12**). We first trained random forest classifiers (5-fold repeated cross-validation) on each classification task - a) “deep” (n=5,712) vs. control and b) “shallow” (narrow definition; n=11,321) vs. control - and ranked all 256 candidate features by their permutation-based feature importance. We then selected the top 30% (76/256) from each “outcome-specific” classifier and used these feature sets to train new models for the opposite classification task. Specifically, the 76 most important features identified from the shallow vs control random forest classifier were used to train a new deep vs control random forest classifier, and conversely, the 76 most important features from the deep vs control classifier were used to train a shallow vs control classifier. Importantly, we transferred model features but not weights.

As summarized in **Table 3**, feature transfer generally yielded performance metrics comparable to the outcome-specific models. For the deep vs control task, transferring top features derived from the shallow phenotype resulted in an AUC of 0.843 (95% CI: 0.831-0.855) and an AUPRC of 0.773 (95% CI: 0.756-0.789), compared to an AUC of 0.840 (95% CI: 0.828-0.852) and an AUPRC of 0.745 (95% CI: 0.726-0.763) achieved by models trained exclusively on deep depression and controls. In the shallow vs. control task, applying deep-derived features yielded an AUC of 0.764 (95% CI: 0.745-0.775) with an F1 score nearly identical to that of the outcome-specific shallow classifier (0.628 vs 0.629; sensitivity 0.799, specificity 0.502, and PPV 0.518), while the outcome-specific shallow classifier attained an AUC of 0.779 (95% CI: 0.769-0.790).

**Table 3.** Feature Transfer Performance Metrics. This table presents a comparison of performance metrics between outcome-specific and feature transfer models for both deep vs control and shallow vs control classification tasks. The outcome-specific models utilize the top 30% of features (76 features) trained to discriminate the respective task (i.e., shallow vs control features for the shallow task, deep vs control features for the deep task), while the feature transfer models apply the top 30% of features trained on the alternative task. For the shallow vs control analysis, only cases meeting the narrow definition of shallow depression (i.e., exclusively meeting criteria for shallow depression without deeper phenotypes) were included. Performance metrics include F1 Score, Sensitivity, Specificity, Positive Predictive Value (PPV), Negative Predictive Value (NPV), Area Under the Receiver Operating Characteristic Curve (AUC), and Area Under the Precision-Recall Curve (AUPRC). Values in parentheses reflect the 95% confidence intervals around the point estimates for each metric, providing a measure of statistical uncertainty. Confidence intervals were estimated using a bootstrap resampling method with 10,000 iterations. For the deep vs control task, the outcome-specific and feature transfer models yielded similar F1 scores (0.585 [95% CI: 0.568-0.602] vs 0.579 [95% CI: 0.563-0.596], respectively) and identical sensitivity (0.799). However, the feature transfer model exhibited marginal improvements in AUC point estimates (0.842 vs 0.840) and AUPRC (0.773 vs 0.745), alongside slight reductions in specificity (0.580 vs 0.689) and PPV (0.455 vs 0.462). In the shallow vs control task, both models achieved similar F1 scores (0.629 [95% CI: 0.617-0.641] vs 0.632 [95% CI: 0.620-0.644], and similar specificity (0.504 vs 0.512) and PPV (0.519 vs 0.522). The outcome-specific model showed a higher AUC (0.779 [95% CI: 0.769-0.790] vs 0.766 [95% CI: 0.755-0.776]) and AUPRC (0.790 vs 0.756). Stratified bootstrap tests (10,000 iterations) for F1 score and AUC indicated no statistically significant differences between feature transfer and outcome-specific models, for either deep vs control or shallow vs control tasks. McNemar’s chi-squared tests were highly significant for both tasks (deep vs control: χ^2^ = 237.84, p < 2.2e-16; shallow vs control: χ^2^ = 386.68, p < 2.2e-16). These significant McNemar’s test results reveal that despite comparable aggregate performance metrics, the outcome-specific and feature transfer models use distinct classification strategies, making different decisions in individual cases.

| Deep vs Control Task |  |  |  | Shallow vs Control Task |  |  |  |
| --- | --- | --- | --- | --- | --- | --- | --- |
| Metrics | Outcome-Specific<br>(Deep vs Control Features) |  | Feature Transfer<br>(Shallow vs Control Features) | Metrics | Outcome-Specific<br>(Shallow vs Control Features) |  | Feature Transfer<br>(Deep vs Control Features) |
| F1 Score | 0.585 (0.568, 0.602) |  | 0.579 (0.563, 0.596) | F1 Score | 0.629 (0.617, 0.641) |  | 0.632 (0.620, 0.644) |
| Sensitivity | 0.799 (0.780, 0.818) |  | 0.799 (0.780, 0.818) | Sensitivity | 0.799 (0.786, 0.813) |  | 0.799 (0.786, 0.813) |
| Specificity | 0.689 (0.676, 0.702) |  | 0.680 (0.667, 0.693) | Specificity | 0.504 (0.490, 0.518) |  | 0.512 (0.498, 0.526) |
| PPV | 0.462 (0.444, 0.480) |  | 0.455 (0.437, 0.473) | PPV | 0.519 (0.505, 0.532) |  | 0.522 (0.509, 0.536) |
| NPV | 0.912 (0.902, 0.920) |  | 0.911 (0.901, 0.919) | NPV | 0.791 (0.776, 0.804) |  | 0.793 (0.779, 0.806) |
| AUC | 0.840 (0.828, 0.852) |  | 0.843 (0.831, 0.855) | AUC | 0.779 (0.769, 0.790) |  | 0.766 (0.755, 0.776) |
| AUPRC | 0.745 (0.726, 0.763) |  | 0.773 (0.756, 0.789) | AUPRC | 0.790 (0.779, 0.801) |  | 0.756 (0.744, 0.769) |
| Stratified Bootstrap | Observed Difference | Permuted Difference | P-value (FDR | Stratified Bootstrap | Observed Difference | Permuted Difference | P-value (FDR |

**Table 3 | Feature Transfer Performance Metrics.**
| Test (10,000) |  | (95% CI) | corrected) | Test (10,000) |  | (95% CI) | corrected) |
| --- | --- | --- | --- | --- | --- | --- | --- |
| **F1 Score** | 0.211 | 0.188, 0.235 | 0.499 | **F1 Score** | -0.130 | -0.142, -0.118 | 0.497 |
| **AUC** | 0.003 | -0.003, 0.010 | 0.511 | **AUC** | -0.013 | -0.018, -0.009 | 0.513 |
| **McNemar's chi-squared test** |  |  |  | **McNemar's chi-squared test** |  |  |  |
| $\chi^2 = 237.84$ | | df = 1 | < 2.2e-16 | $\chi^2 = 386.68$ | | df = 1 | < 2.2e-16 |

To evaluate whether any nominal performance differences between the feature transfer and outcome-specific models might be influenced by the larger sample size of the shallow cohort, we randomly downsampled the shallow cohort to match the case/control ratio (approximately 1:3 with 5,712 cases) of the deep vs. control classifier. In this downsampled analysis, the shallow vs control classifier (top 30% of features) yielded an F1 score of 0.432 (95% CI: 0.497-0.525), sensitivity of 0.799 (95% CI: 0.780-0.818), specificity of 0.365 (95% CI: 0.352-0.379), PPV of 0.296 (95% CI: 0.283-0.310), NPV of 0.845 (95% CI: 0.830-0.860), AUC of 0.690 (95% CI: 0.674-0.706), and AUPRC of 0.554 (95% CI: 0.531-0.575). When comparing these results to the non-downsampled shallow classifier (n=11,321; see Methods and **Table 2**), the downsampled model showed performance degradation across most metrics, including reduced PPV (ΔPPV = −0.245; 0.296 vs 0.541), AUC (ΔAUC = −0.094; 0.690 vs 0.784), F1 score (ΔF1 score = −0.213), and AUPRC (ΔAUPRC = −0.244). These results indicate that sample size reductions negatively impact classifier performance in the shallow vs control task, although this finding should be interpreted cautiously, as they reflect both the loss of training data (reduced from 11,321 to 5712 cases) and the artificially lowered prevalence, which impacts prevalence-sensitive metrics like PPV.

Furthermore, to characterize the cross-phenotype generalizability at the feature level observed within the feature transfer tasks, we examined the specific neuroanatomical correlates that contribute most to the model’s classification performance. Analysis of the top 76 features from each transfer model revealed approximately 47% feature overlap (36 shared features), indicating that while deep and shallow phenotypes share certain neuroanatomical signatures, each also captures distinct patterns (**Supplementary Figures S11-S12**). Across all models, including outcome-specific, feature transfer, and multi-class classifiers, global volumetric measures were consistently among the top rankings, with normalized gray matter volume and peripheral cortical gray matter volume ranking first and second in both deep and shallow outcome-specific models (**Supplementary Table S10**). When the shallow definition cohort was downsampled to match the deep phenotype case/control ratio, the raw importance scores of these top volumetric features in the shallow model also decreased by 31% and 24% respectively, despite maintaining their leading ranks. These results suggest that deep and shallow depression phenotypes share certain core neuroanatomical features while also exhibiting distinct patterns, a finding that remained consistent when controlling for sample size differences.

## Discussion

Efforts to identify the neurobiologic basis of MDD have been complicated by the heterogeneity of clinical presentations and variability in ascertainment and assessment in research cohorts. Many studies have employed shallow (or “minimal”) phenotyping to facilitate the collection of large-scale samples and enhance power. Depth of phenotypic assessment has been shown to impact genomic analyses of MDD[9] but its implications for neuroimaging research have not been systematically studied. Here, we leveraged neuroimaging data from more than 30,000 UK Biobank participants to examine how associations between depression and brain structure vary with phenotypic depth. Three main findings emerged from our analyses. First, regardless of phenotyping depth, the most robust associations with depression case status were found for indices of white matter microstructure (specifically fractional anisotropy and mean diffusivity). Second, neuroanatomical associations were more modest than those observed by the large ENIGMA MDD consortium, possibly reflecting differences in sample composition, depression severity, or measurement approaches. Third—contrary to our hypothesis, neuroanatomical classifier performance was similar regardless of deep vs. shallow case definition. In addition, our feature transfer analyses found that random forest models using the top 30% of features selected from shallow phenotypes performed similarly (by AUC and F1 metrics) in discriminating deep phenotype cases from controls compared to models trained directly on features from deep phenotypes, and vice versa. That said, the ranks of top features did differ for the deep and shallow classifiers, suggesting that biological correlates are not entirely the same (**Supplementary Figure S8**). Of note, the GPNoDep phenotype - help-seeking without depression - showed markedly fewer neuroanatomical associations than any depression phenotype (**Supplementary Table S2**), consistent with genetic evidence that GPNoDep is related to non-specific help-seeking rather than depression-specific biology[9].

As noted, white matter microstructure showed the strongest and most consistent associations with depression status across phenotype definitions, in line with prior large biobank and multi-site studies identifying white matter alterations in depression[36, 41–44]. For example, Nothdurfter et al. (2024)[45] analyzed data from the UKBB across 19,183 individuals (including 5823 with varying levels of depressive symptoms), demonstrating reduced white matter integrity in thalamic and intracortical fiber tracts in depression (corresponding to our “self-reported depression” shallow phenotype), regardless of current symptom level. These findings contrast, however, with an earlier 12-site mega-analysis by Koshiyama et al. (2020)[42], which found no significant differences in DTI indices between MDD patients (n=398) and healthy controls (n=1,506), potentially highlighting the value of large sample sizes in detecting subtle neuroanatomical differences in depression.

The causal relationship of white matter changes in depression remains uncertain. However, longitudinal findings from Flinkenflügel et al. (2024)[46], analyzing diffusion-weighted imaging data from 418 MDD patients and 463 healthy controls over two years, demonstrated that MDD patients show steeper declines in fractional anisotropy in the superior longitudinal fasciculus compared to healthy controls; these changes were associated with cognitive decline. This suggests that white matter integrity may be relevant to depression and related cognitive impairment in at least a subset of individuals. Consistent with this directionality, evidence from Mendelian randomization studies has supported a causal role of depression on white matter integrity [47, 48]. In contrast to the robust white matter findings, cortical thickness and surface area measures showed weaker and more variable associations across depression phenotypes.

Though prior meta-analyses have identified gray matter reductions in regions such as the hippocampus, amygdala, and cingulate cortex in MDD[2, 49, 50], we did not find strong evidence of this. In another study using UKBB data, Harris et al. (2022)[10] similarly reported widespread reductions in fractional anisotropy and increases in mean diffusivity across multiple depression definitions using data from the UKBB; however, they also observed associations between cortical thickness and depression status, particularly in self-reported phenotypes. This discrepancy may reflect differences in analytical approaches. For example, Harris et al.[10] aggregated effects across several cortical regions structures, reducing the burden of correction for multiple comparisons and perhaps enhancing power to detect some associations. Our more stringent multiple comparison approach, combined with greater regional specificity, likely contributed to our more variable cortical associations.

The reproducible pattern of white matter microstructural alterations across independent cohorts and varied analytical approaches points to their potential relevance as therapeutic targets. Consistent with this broader literature[36, 44–46], our analyses revealed increases in fractional anisotropy (FA) and decreases in mean diffusivity (MD) across multiple depression phenotypes, with significant differences and directional consistency in associations across regions including the posterior or anterior limb of the internal capsule, various corpus callosum subregions, and association fibers in the corona radiata. These pathways are related to circuits historically targeted by neurosurgical ablation, such as subcaudate tractotomy, which targeted dysfunctional frontal–subcortical circuits[51]. Modern interventions such as electroconvulsive therapy (ECT), repetitive transcranial magnetic stimulation (rTMS), and deep brain stimulation (DBS), now refine this rationale by leveraging neuroimaging to guide precise, non-ablative modulation of these circuits.

Emerging evidence suggests white matter remodeling as a mechanism of action across neuromodulatory depression treatments, reinforcing the clinical relevance of the white matter alterations we observed. ECT studies have demonstrated both transient increases in right-hemisphere mean diffusivity and progressive increases in fractional anisotropy in dorsal fronto-limbic pathways that correlate with symptom improvement[52, 53]. Similarly, rTMS appears to induce microstructural changes in prefrontal white matter tracts, with studies reporting increased fractional anisotropy in anterior-medial prefrontal bundles following treatment, despite minimal changes in directly stimulated pathways[54, 55]. DBS represents a more invasive approach for treatment-resistant depression (TRD), directly targeting subcortical regions and critical white matter tracts. Studies of subcallosal cingulate DBS have shown that pre-existing structural abnormalities in specific pathways, particularly the forceps minor, uncinate fasciculus, and cingulum bundle, correlate with longer recovery times in depression[56], while supero-lateral medial forebrain bundle stimulation has demonstrated high response rates with sustained effects in TRD[57]. Collectively, these findings suggest that while different neuromodulatory treatments target distinct neural circuits, their therapeutic effects involve white matter remodeling, with evidence that both treatment-induced changes and baseline structural integrity of white matter tracts, measured through fractional anisotropy and mean diffusivity, relate to clinical outcomes in depression.

While ECT, rTMS, and DBS studies suggest white matter pathways mediate treatment responses, our comparison revealed that depression-related alterations in fractional anisotropy and mean diffusivity in UKBB were markedly smaller than those reported by the ENIGMA MDD consortium[36], after converting our odds ratios to Cohen’s d effect sizes, as shown in Figure 3. Notably, this comparison was restricted to 20 white matter tracts common to both datasets, with UKBB hemisphere-averaged values aligned to ENIGMA’s bilateral reporting framework (see Supplementary Methods). The strongest alignment with effect size estimates reported by the ENIGMA MDD consortium were observed in electronic medical record-based depression definitions as shown in Figure 3, though the magnitude of depression-associated neuroanatomical alterations remained smaller than those reported in their meta-analysis. These differences in the strength of association between depression status and neuroanatomical measures between UKBB and ENIGMA likely reflect a combination of sample composition, assessment approaches, and demographic factors rather than phenotyping depth alone—a finding reinforced by the similar performance of shallow and deep phenotyping approaches in our analyses.

Having characterized the pattern and magnitude of neuroanatomical associations across phenotyping approaches, we next evaluated their utility for depression classification through machine learning techniques. Using 256 neuroanatomical features, random forest classifiers performed similarly in distinguishing deep or shallow depression phenotypes from controls. In contrast, super learner ensemble classifiers achieved somewhat better performance for distinguishing deep phenotype cases from controls as opposed to shallow vs. controls on AUC (0.911 vs 0.764), specificity (0.862 vs 0.701), and sensitivity (0.785 vs 0.663), as shown in **Supplementary Table S4**. Nevertheless, the comparable performance of features derived from shallow phenotypes in our feature transfer analyses combined with larger sample sizes facilitated by shallow phenotyping approaches, provides some justification for using simpler operational definitions when developing neuroanatomical classifiers of depression. However, downsampling shallow phenotype cases to match the deep phenotype’s case/control ratio resulted in performance degradation, suggesting that shallow phenotypes required approximately twice the sample size to achieve comparable performance. This reflects the power-specificity tradeoff in psychiatric genetics and neuroimaging, where broader phenotypic definitions capture meaningful biological signals but require substantially larger samples to compensate for lower measurement precision [9, 58–60].

Several important limitations of our study warrant consideration. First, while the UK Biobank provides an unprecedented scale for neuroimaging analyses, it is not representative of the general population[61]. Second, our depression phenotypes, while spanning a broad spectrum, rely heavily on retrospective self-report and may be subject to recall bias. The absence of structured clinical interviews for all participants may have resulted in information bias in our phenotypic classifications. Third, our classification of ICD-10 diagnoses of depression as “deep” phenotypes warrants clarification. While we categorized ICD-10 diagnoses as deep phenotypes because they represent clinician-assigned codes rather than self-report, these codes have inherent limitations (we cannot determine specific clinical characteristics prompting diagnosis, and codes may be assigned for administrative reasons), potentially positioning them as intermediate between self-report measures and CIDI-SF derived phenotypes. Fourth, the UK Biobank sample is relatively homogeneous in terms of race/ethnicity; generalizability of these findings, especially to more diverse populations, may be limited. Fifth, although our multiple “shallow” and “deep” definitions capture different ways to measure or classify depression, these do not necessarily correspond to distinct biological subtypes of depression. Depression is known to be heterogeneous with subtypes that may differ in symptom profiles, course, and neurobiological underpinnings. Our findings speak more to the implications of measurement variability rather than to intrinsic biological subgroups within the disorder. Consequently, while we can compare the predictive utility of different operational definitions (e.g., self-report versus ICD-10 diagnosis), we cannot disentangle all forms of heterogeneity that naturally exist within depression.

In summary, our large-scale analysis reveals that alterations in white matter microstructure are observed across a spectrum of depression phenotype definitions. While the neuroanatomical associations observed in UKBB were of lower magnitude than those reported by the ENIGMA MDD consortium, the directional pattern of white matter changes across different phenotyping approaches (with decreased fractional anisotropy and increased mean diffusivity), aligns with prior studies[36, 43–46], and suggests these alterations represent a replicable biomarker of depression. Although shallow and deep phenotypes achieved comparable classification performance in our machine learning analyses, this was contingent on shallow phenotypes having approximately twice the sample size, underscoring the fundamental power-specificity tradeoff in psychiatric neuroimaging research.

## Data Availability

The datasets analyzed during the current study are available via the UK Biobank data access process (see http://www.ukbiobank.ac.uk/register-apply/). This research was conducted using the UK Biobank Resource under Application Number 32568.

## Acknowledgements

This work was supported by the Tommy Fuss Fund, a foundation established by the Fuss family to promote medical research that furthers our understanding of mental illness and develops more effective means of diagnosing and treating psychopathology.

