## Supplementary Figures for "Mapping heterogeneity in the neuroanatomical correlates of depression": Supplementary Figure S1 _ Effect sizes across imaging-derived phenotypes and the depression spectrum.docx

| **Fractional Anisotropy** | | | 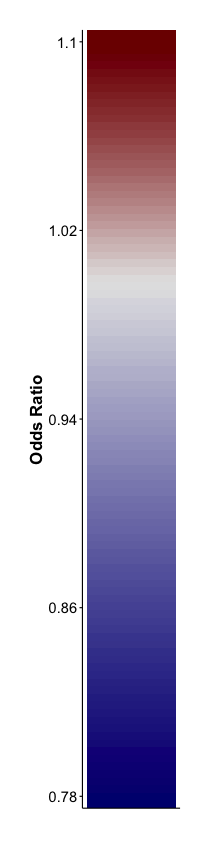 |
| --- | --- | --- | --- |
| **GPNoDep** | **GPpsy** | **Psypsy** |  |
| 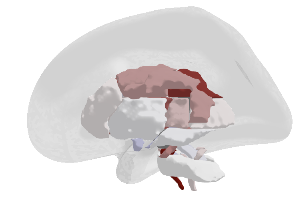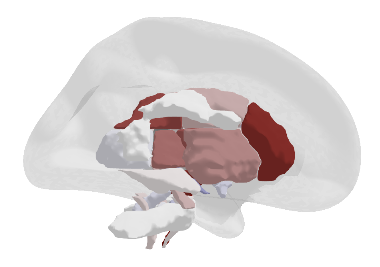 | 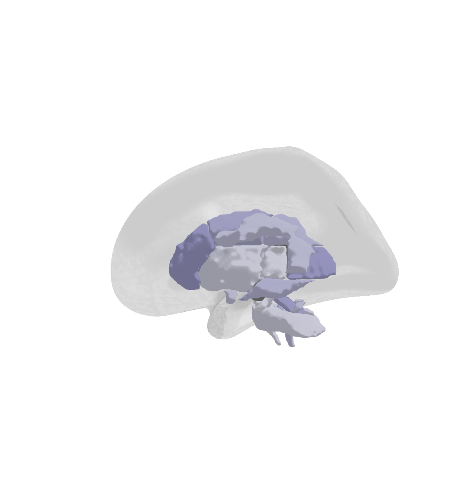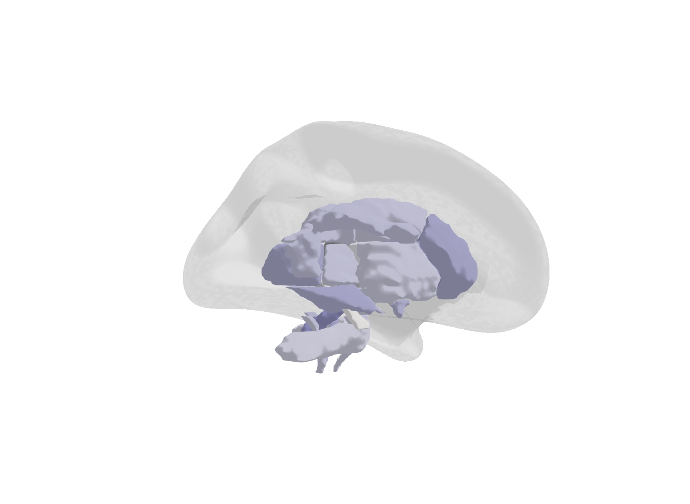 | 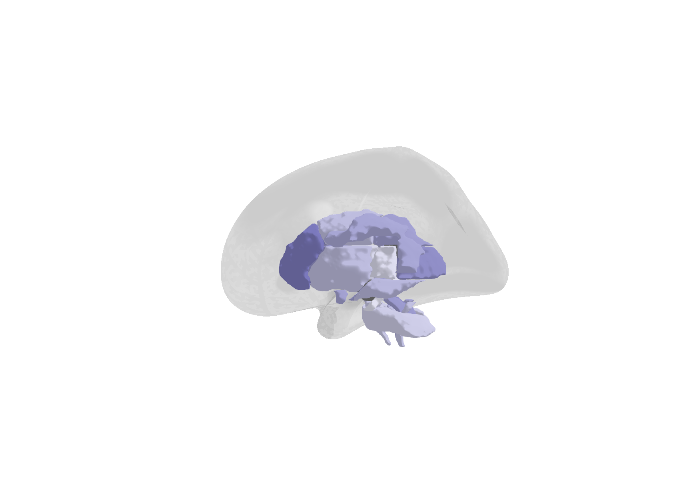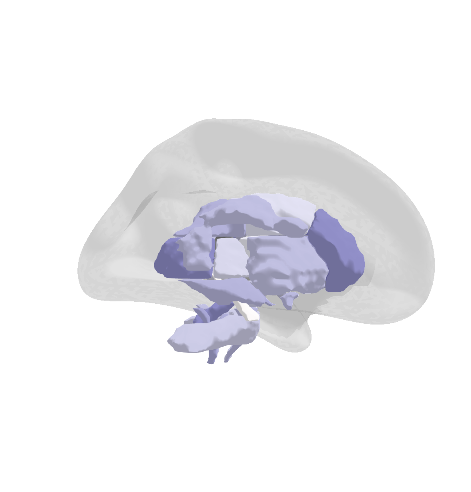 |  |
| **SelfRepDep** | **DepAll** | **ICD10Dep** |  |
| 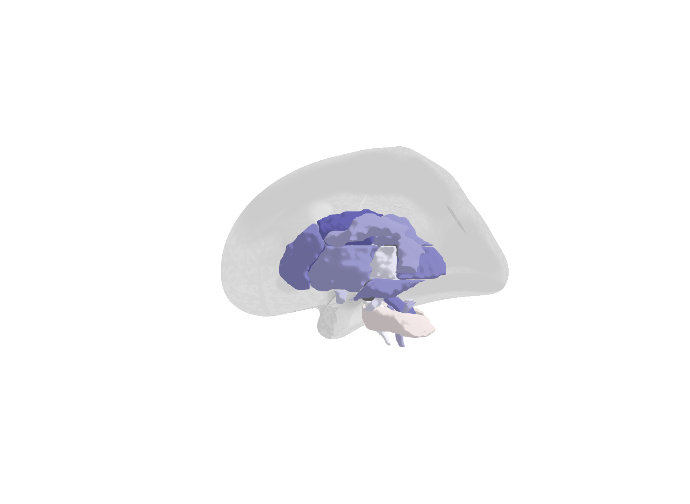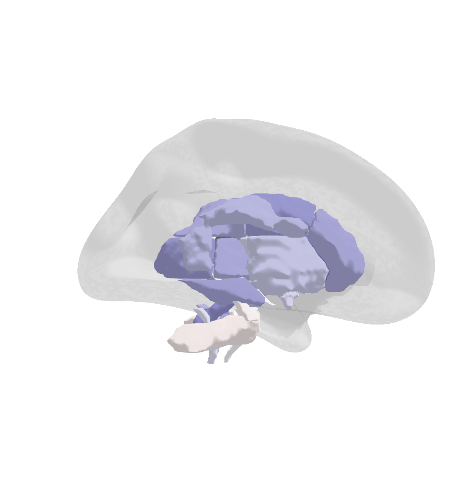 | 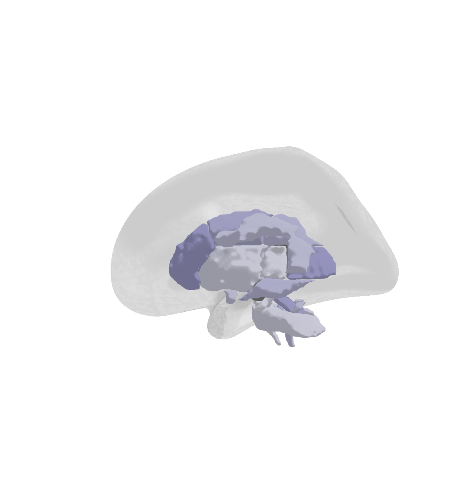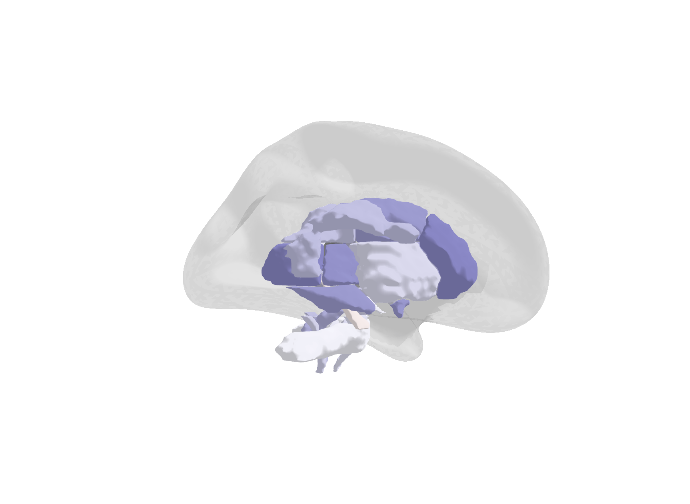 | 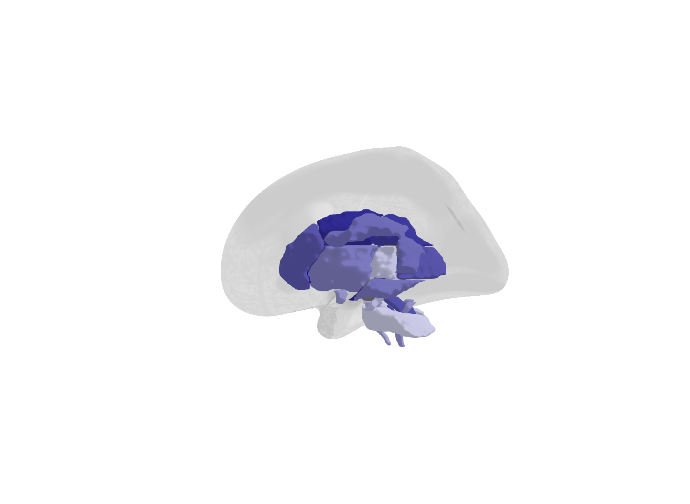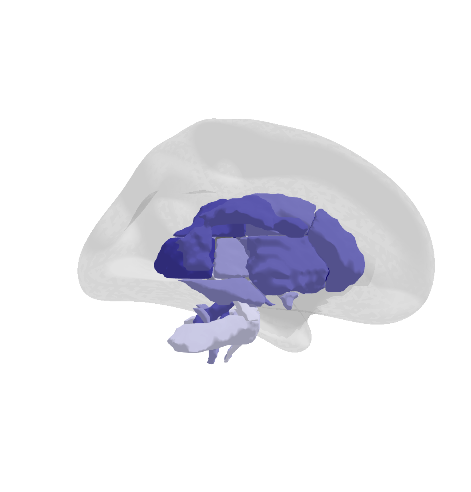 |  |
| **ICD10Dep_exclpsych** | **LifetimeMDD** | **MDDRecur** |  |
| 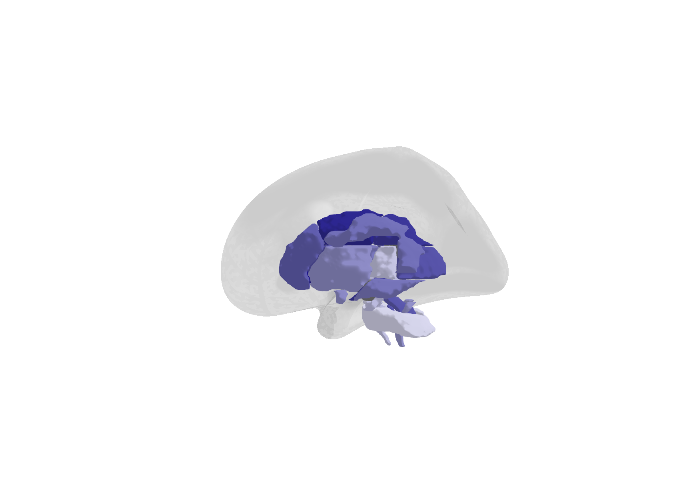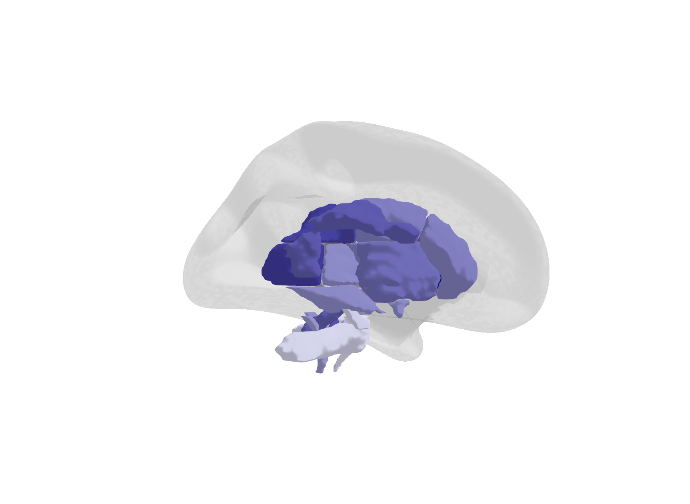 | 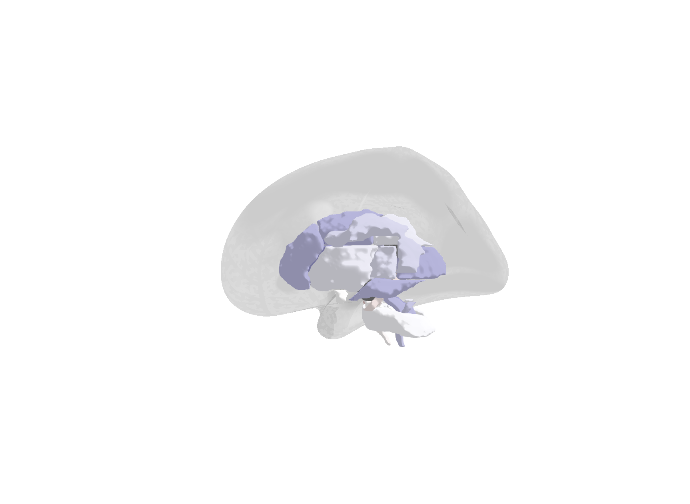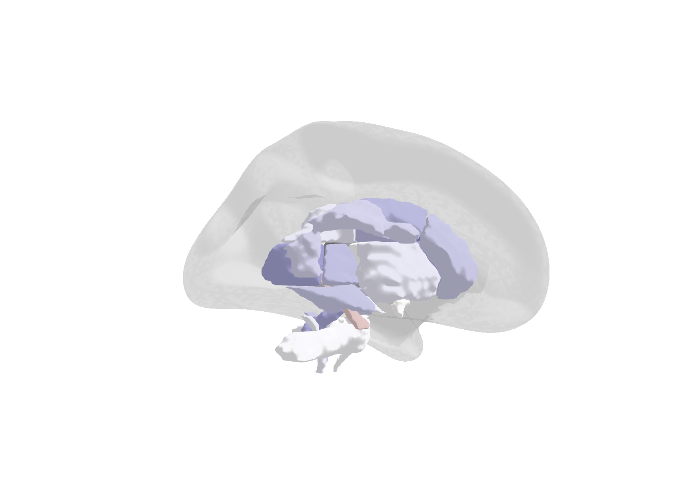 | 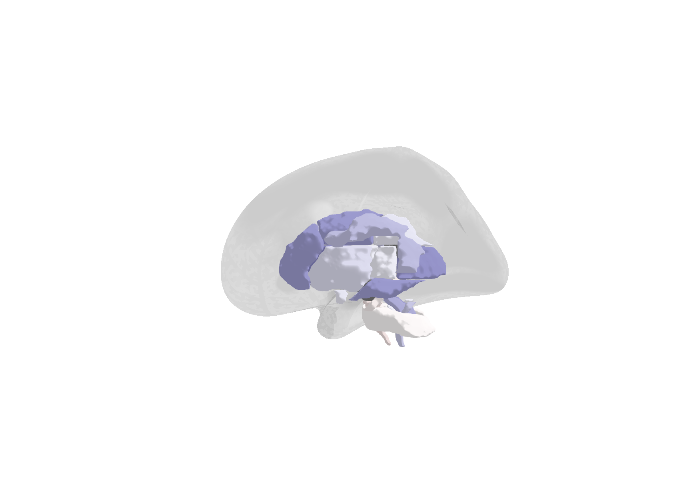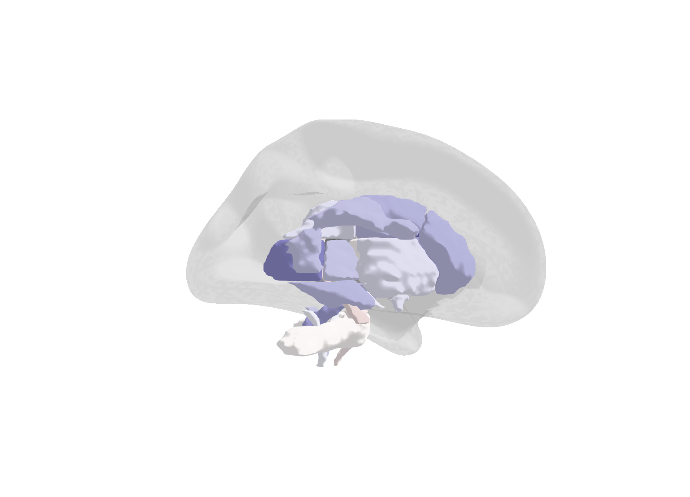 |  |

| **Mean Diffusivity** | | | 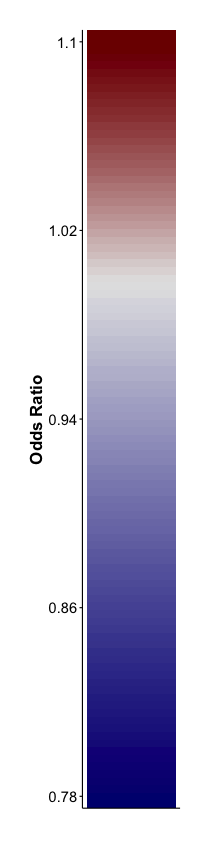 |
| --- | --- | --- | --- |
| **GPNoDep** | **GPpsy** | **Psypsy** |  |
| 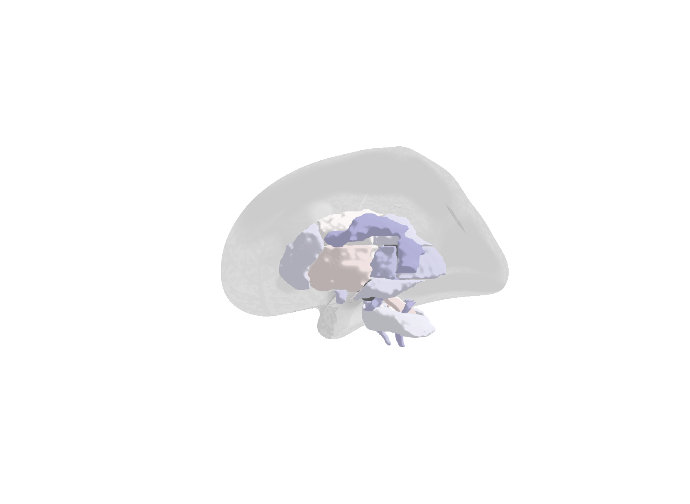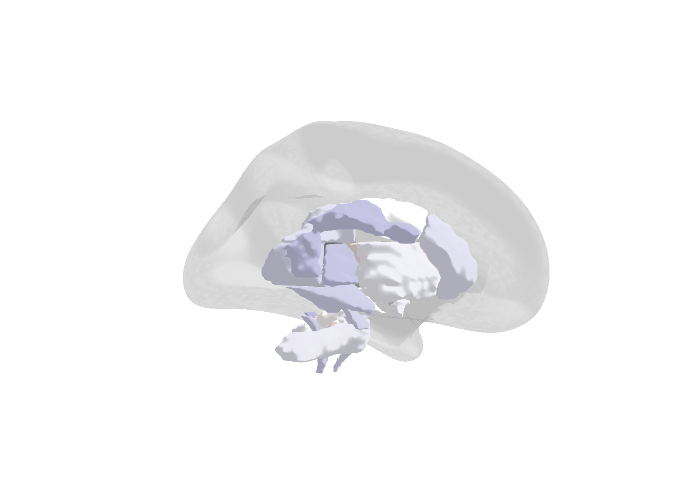 | 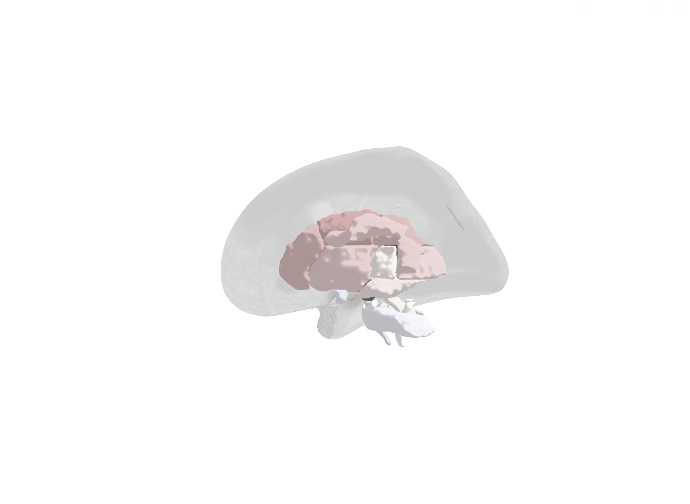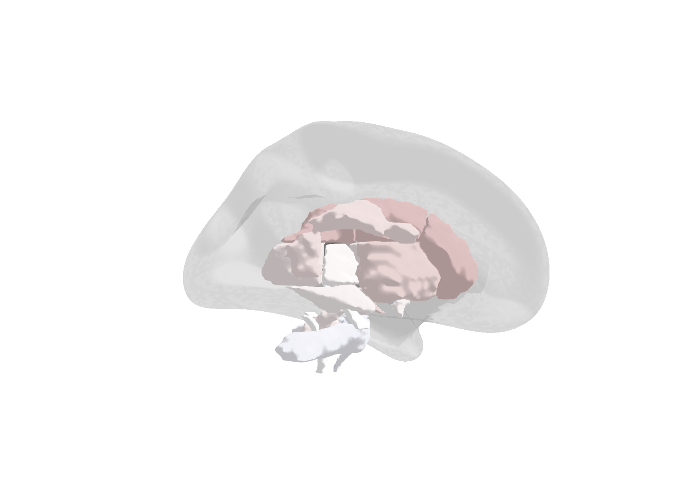 | 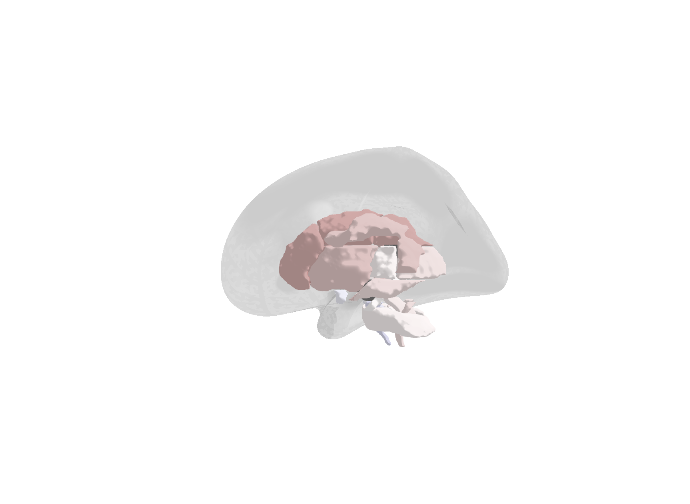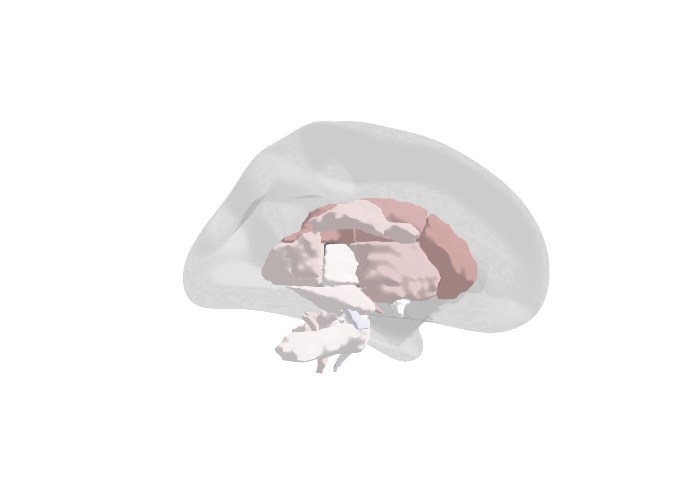 |  |
| **SelfRepDep** | **DepAll** | **ICD10Dep** |  |
| 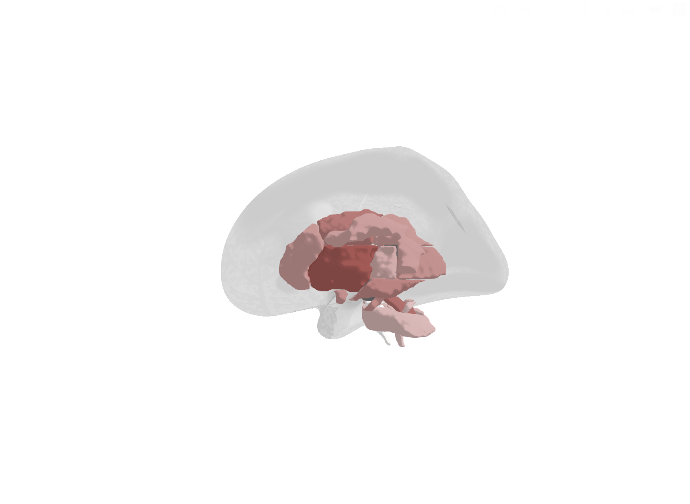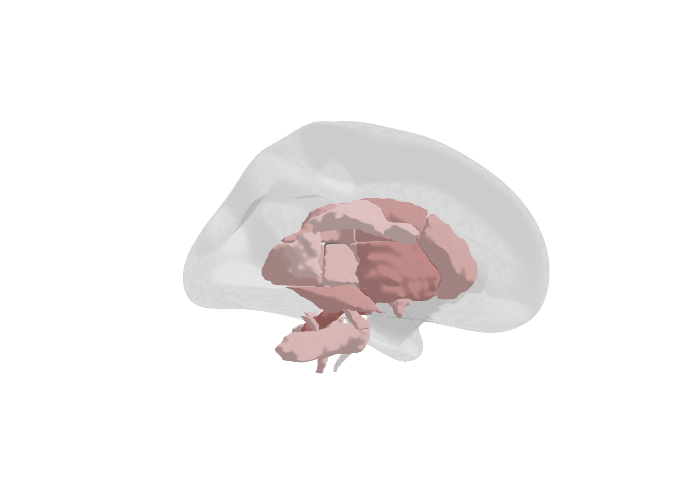 | 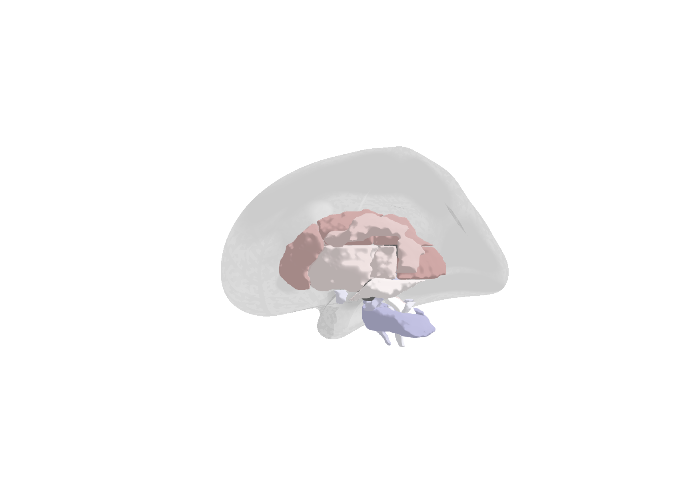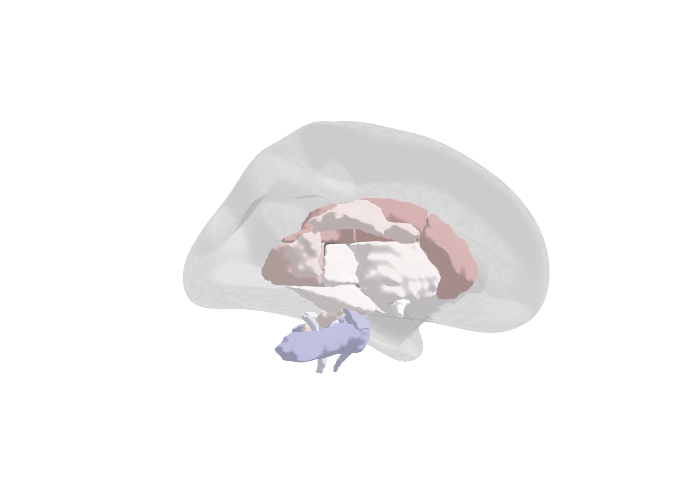 | 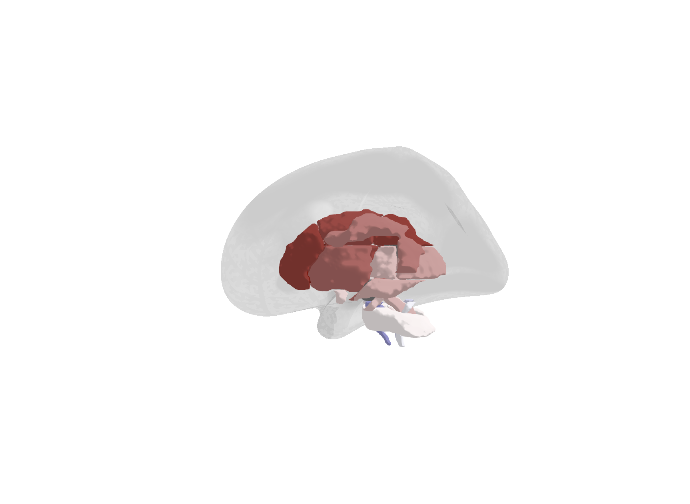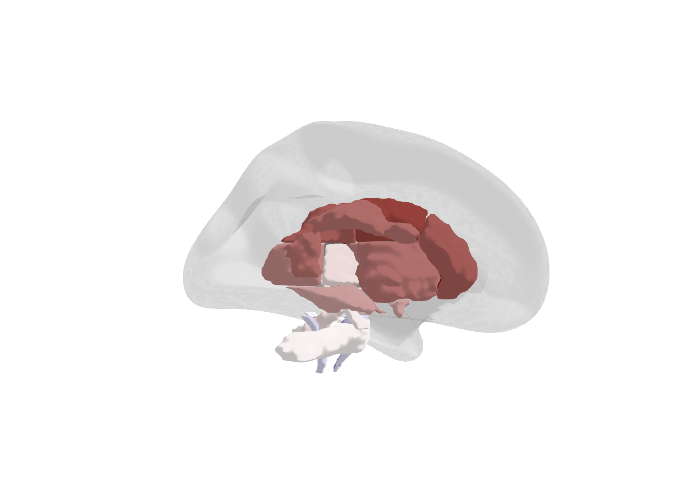 |  |
| **ICD10Dep_exclpsych** | **LifetimeMDD** | **MDDRecur** |  |
| 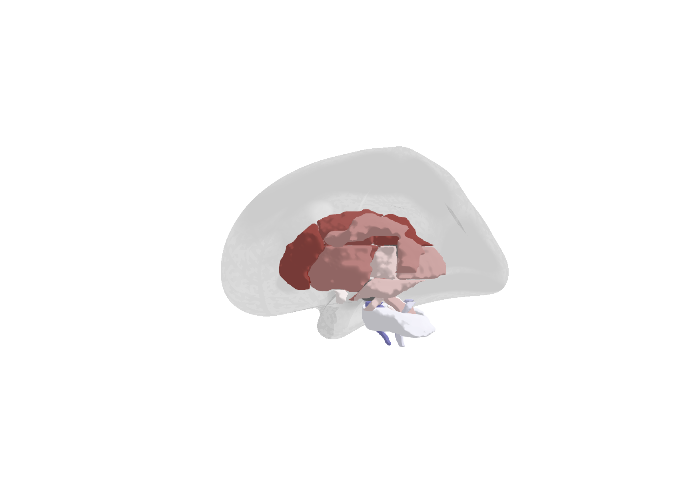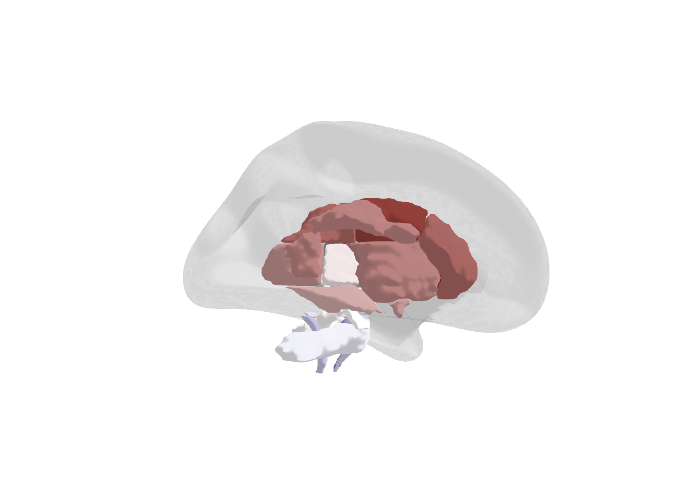 | 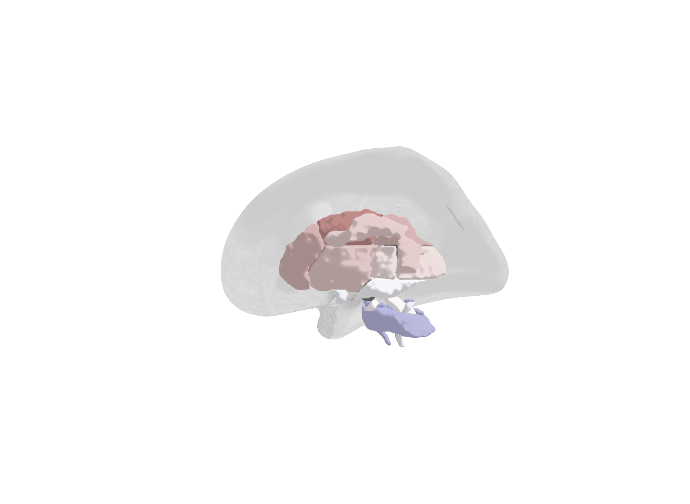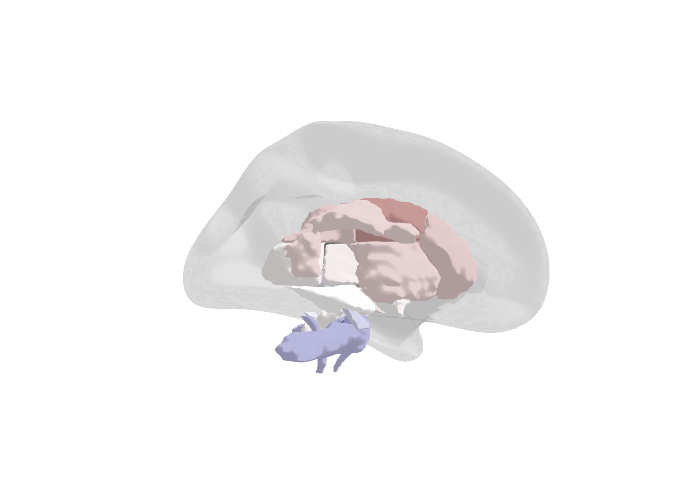 | 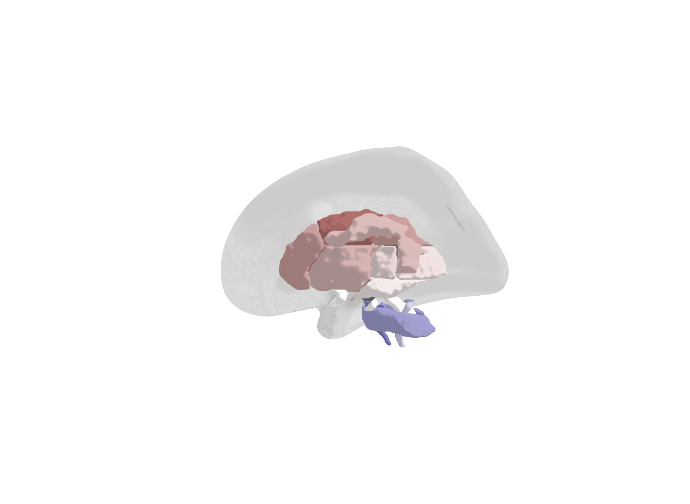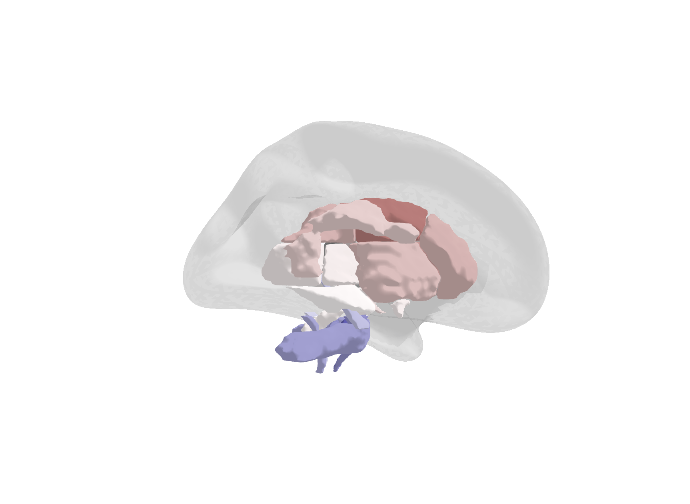 |  |

| **Cortical Thickness** | | | 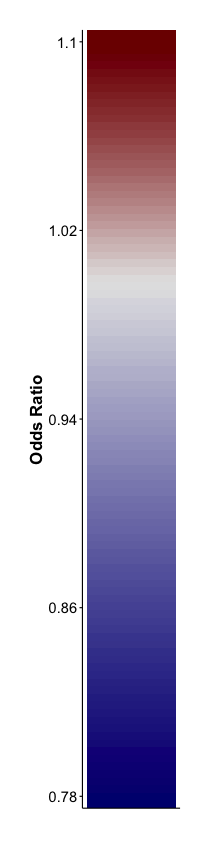 |
| --- | --- | --- | --- |
| **GPNoDep** | **GPpsy** | **Psypsy** |  |
| 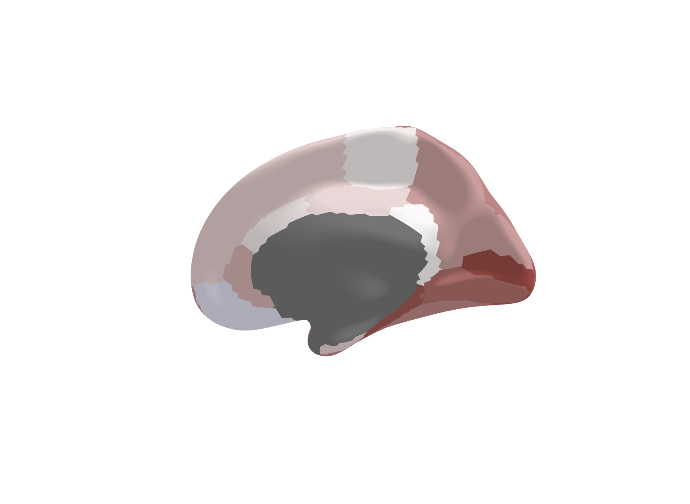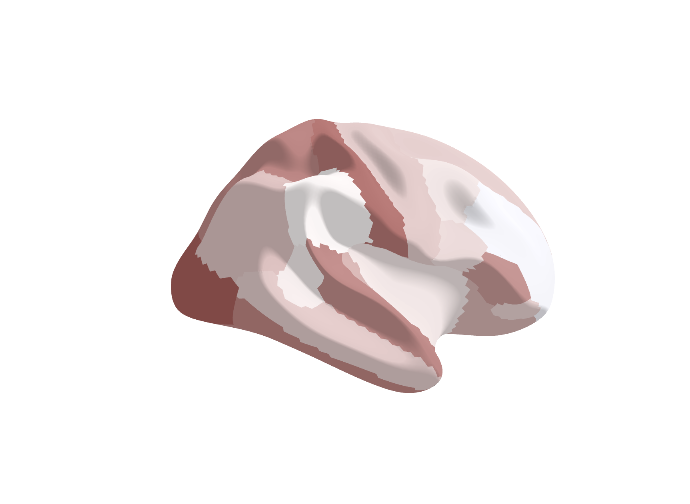 | 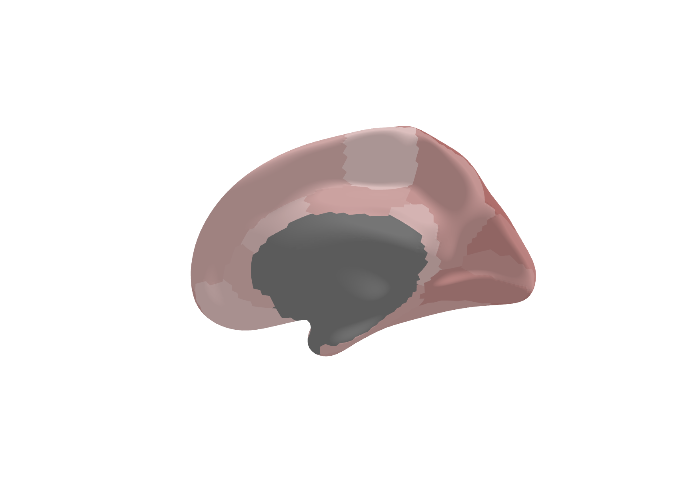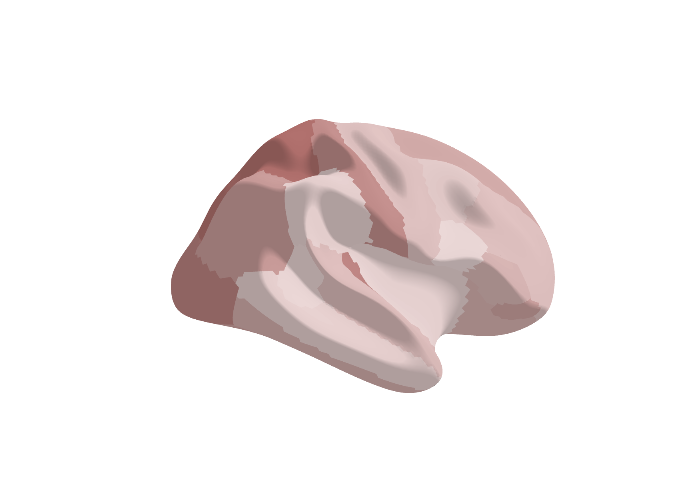 | 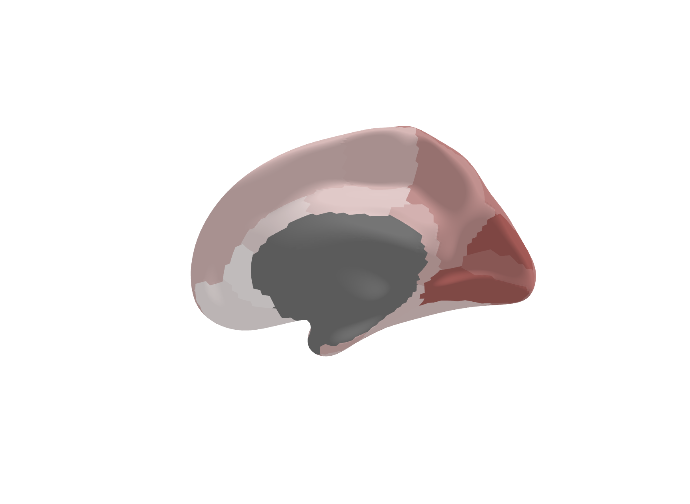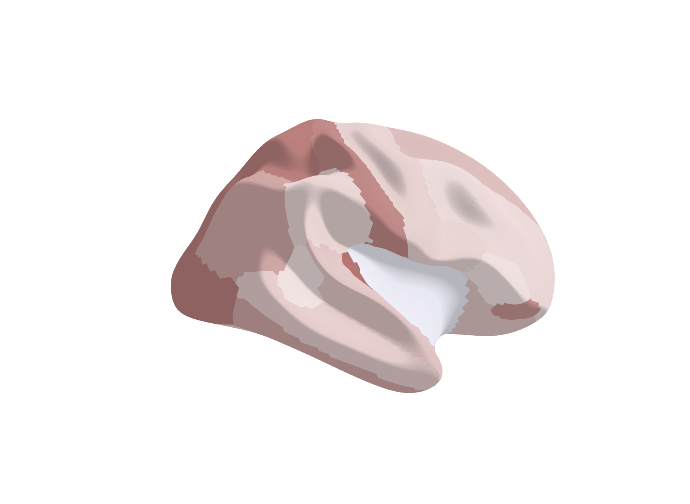 |  |
| **SelfRepDep** | **DepAll** | **ICD10Dep** |  |
| 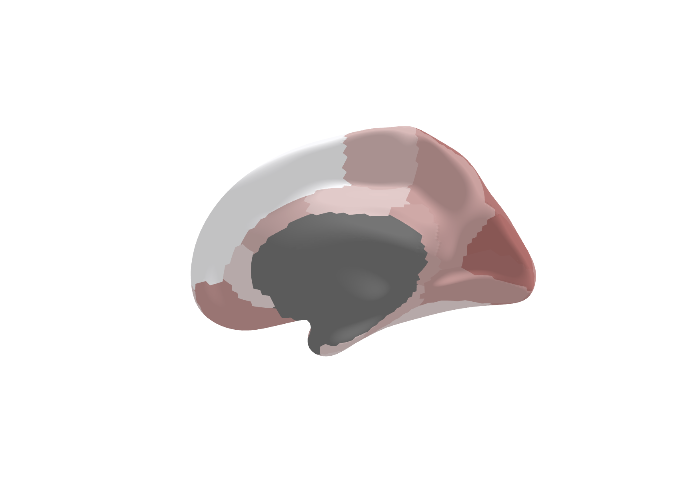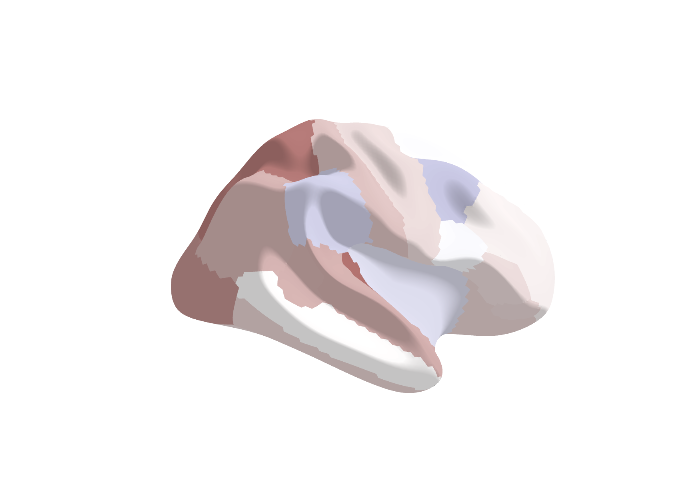 | 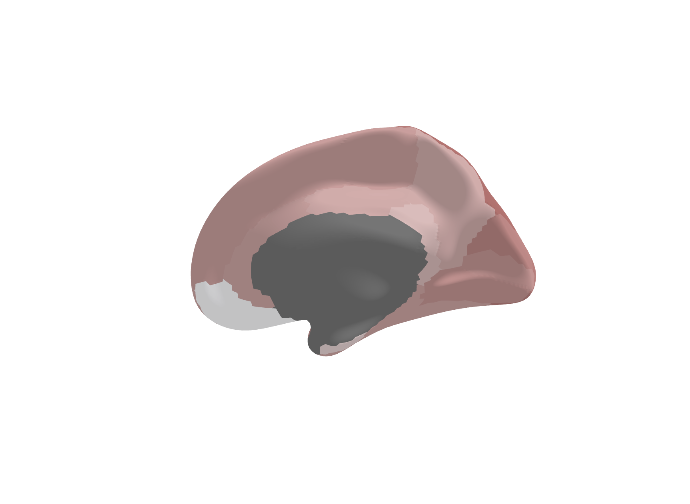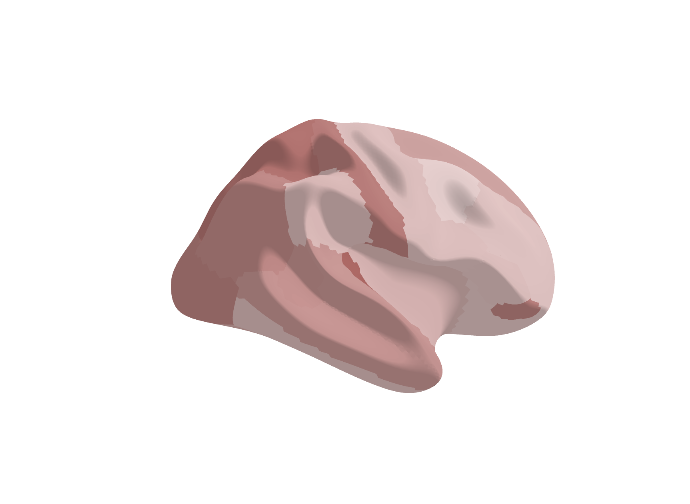 | 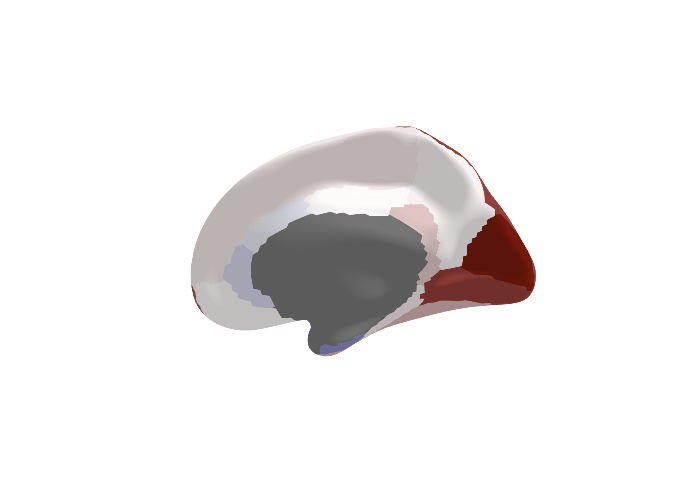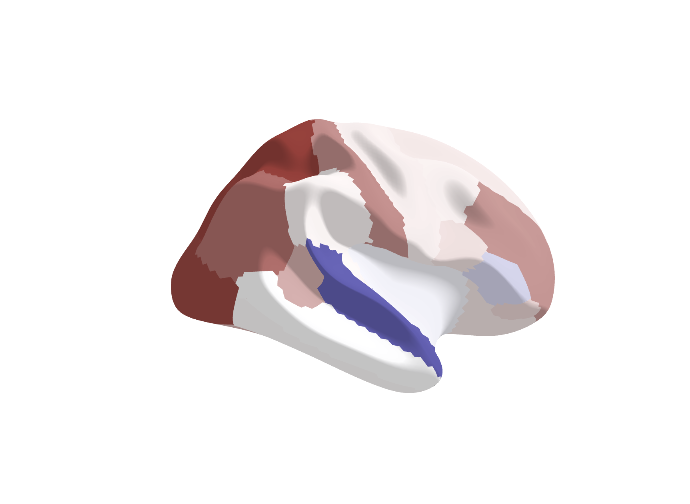 |  |
| **ICD10Dep_exclpsych** | **LifetimeMDD** | **MDDRecur** |  |
| 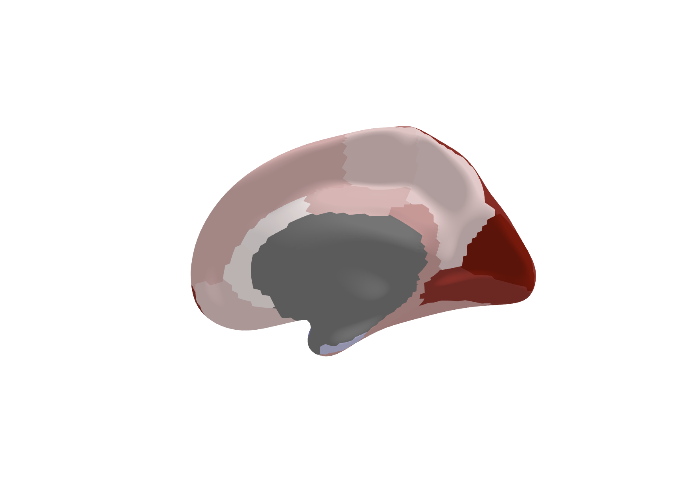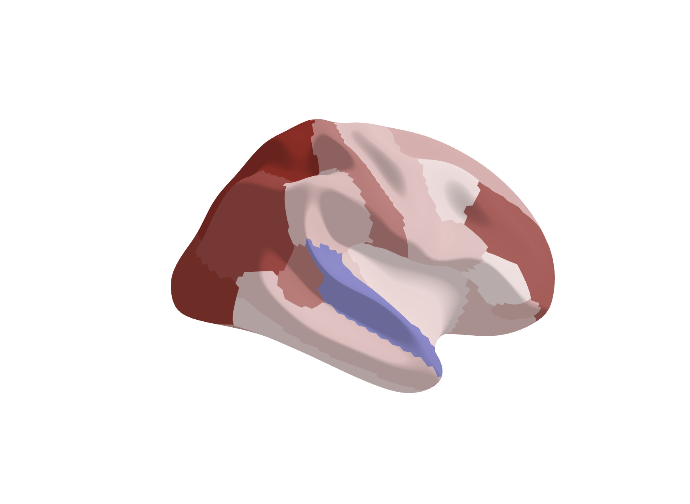 | 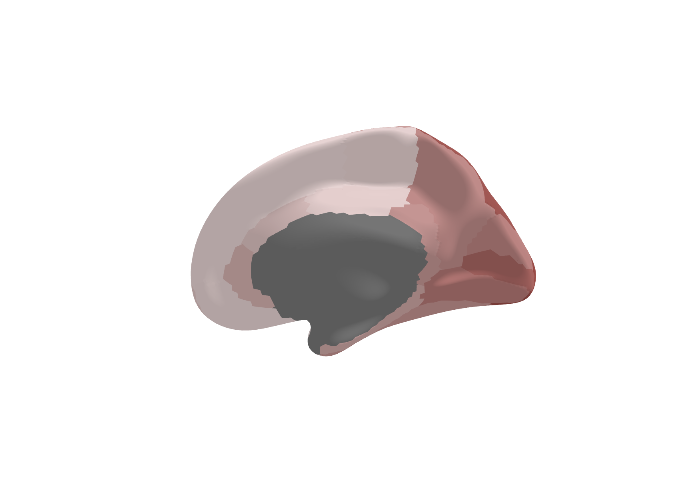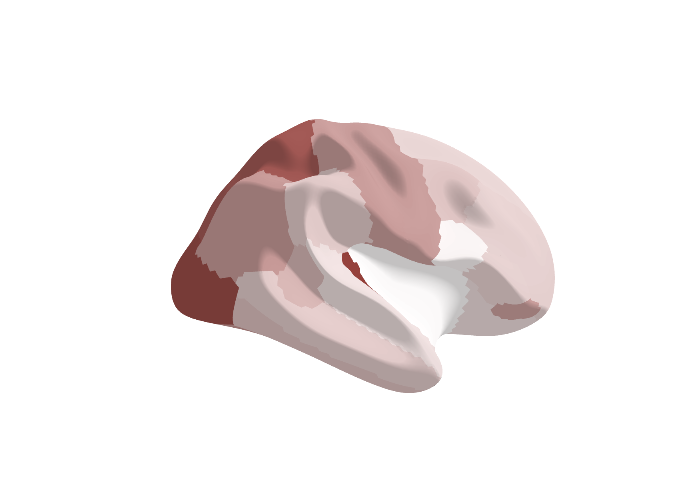 | 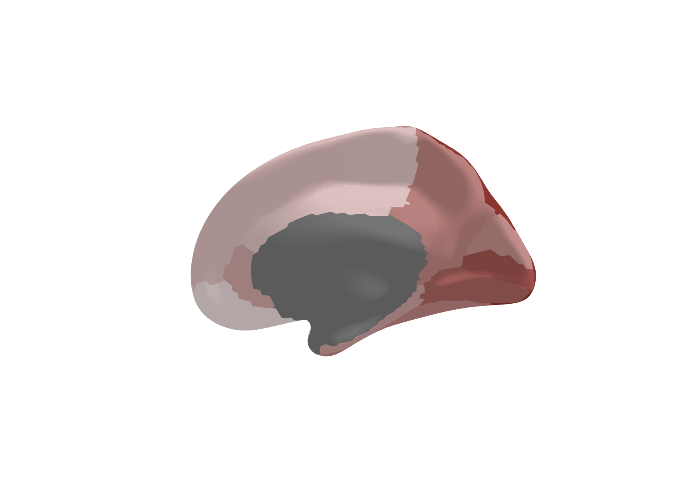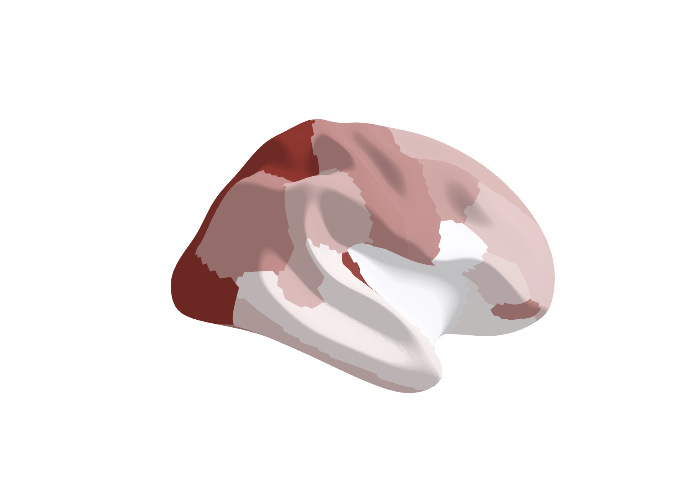 |  |

| **Surface Area** | | | 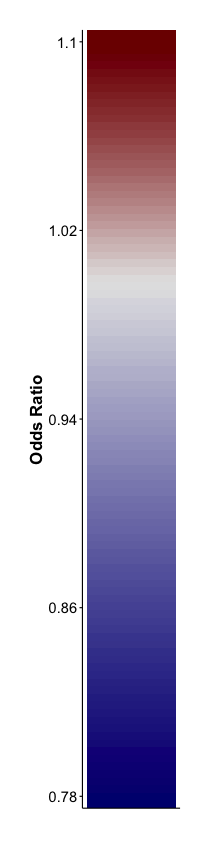 |
| --- | --- | --- | --- |
| **GPNoDep** | **GPpsy** | **Psypsy** |  |
| 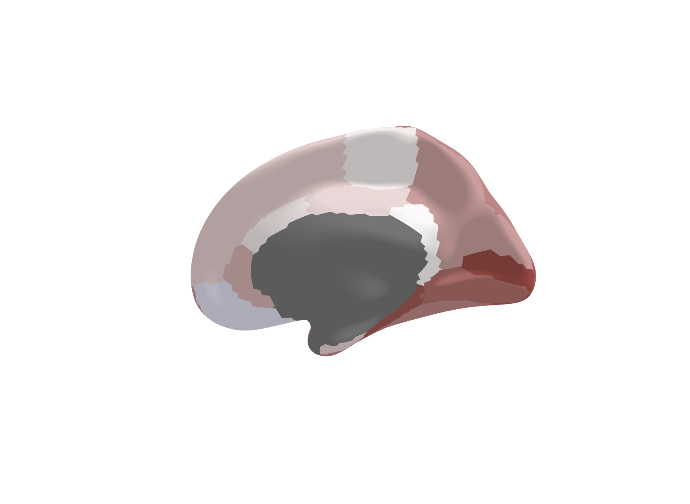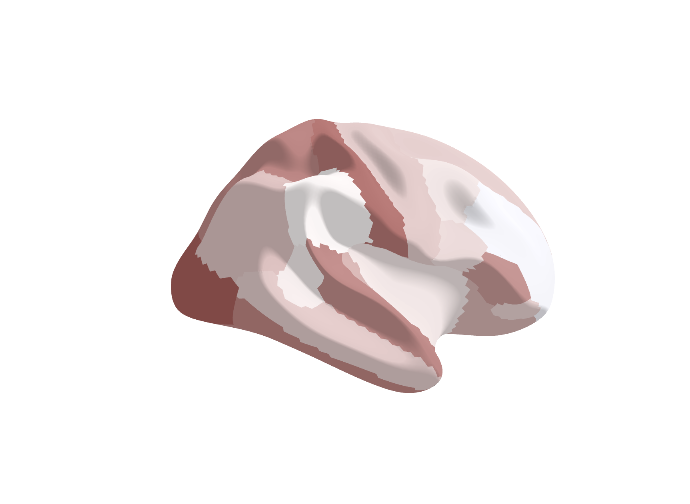 | 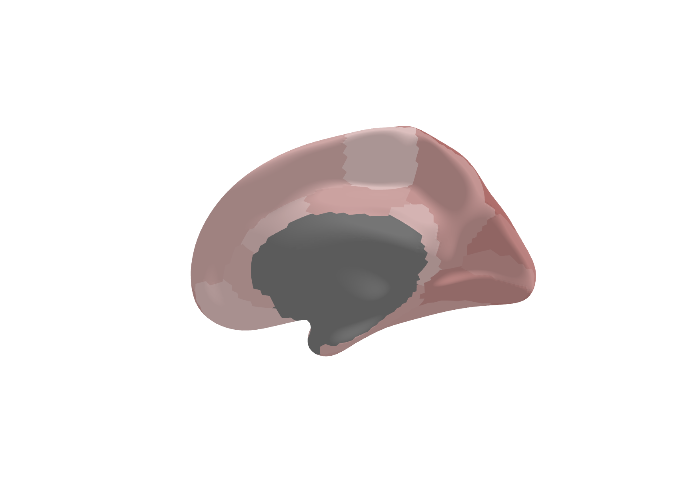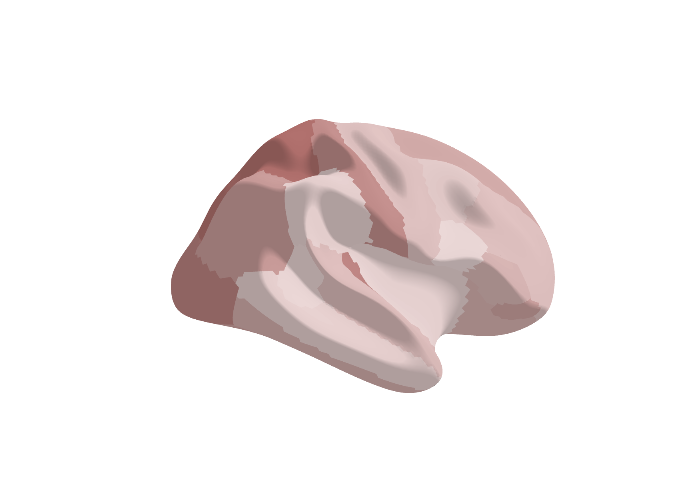 | 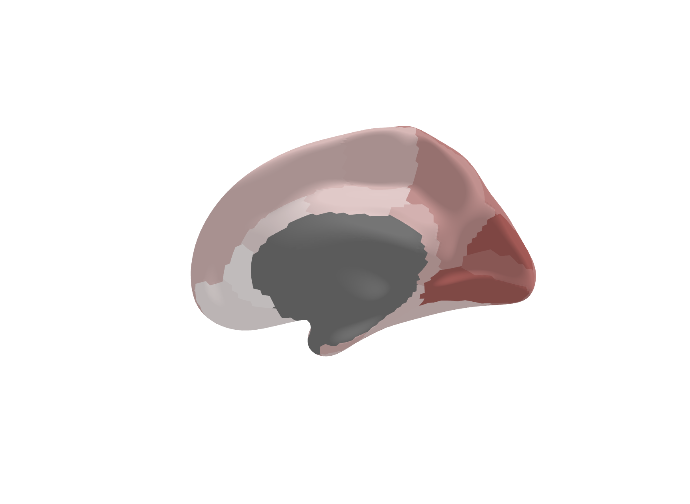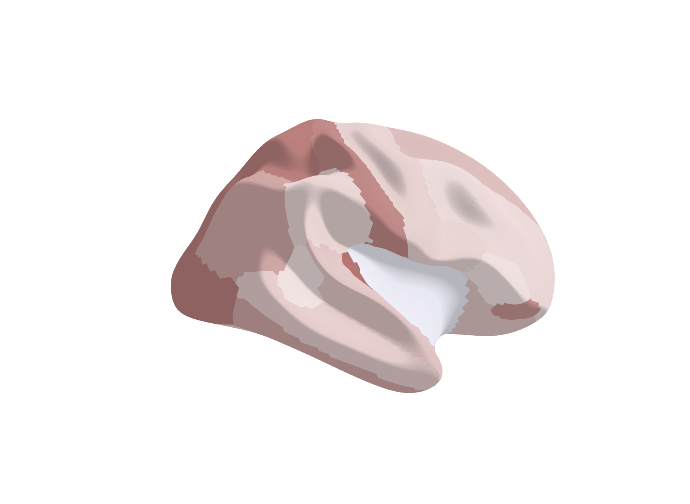 |  |
| **SelfRepDep** | **DepAll** | **ICD10Dep** |  |
| 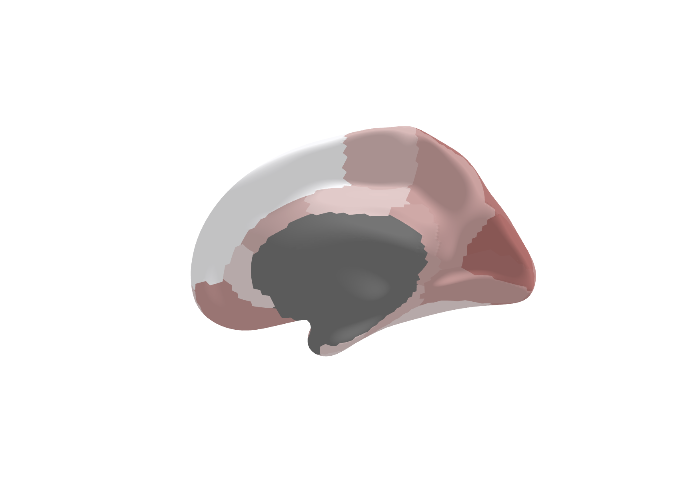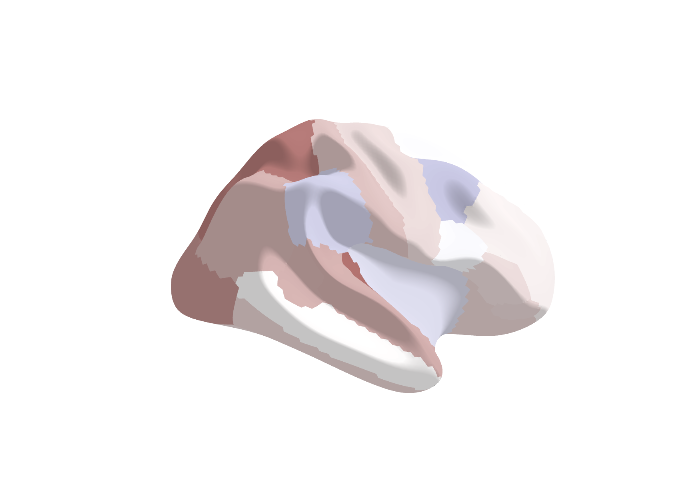 | 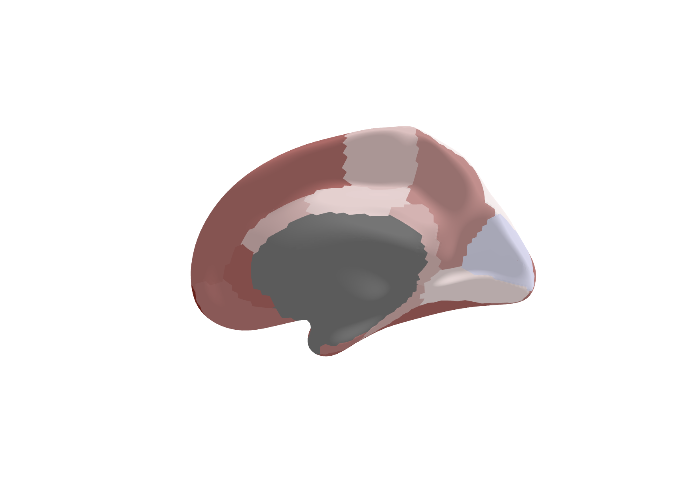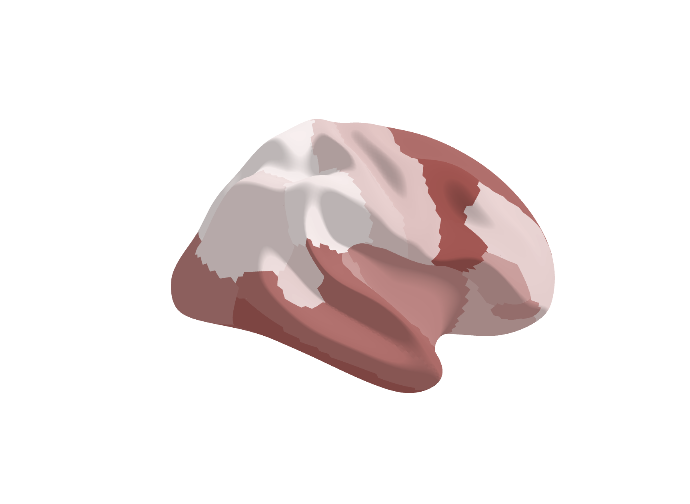 | 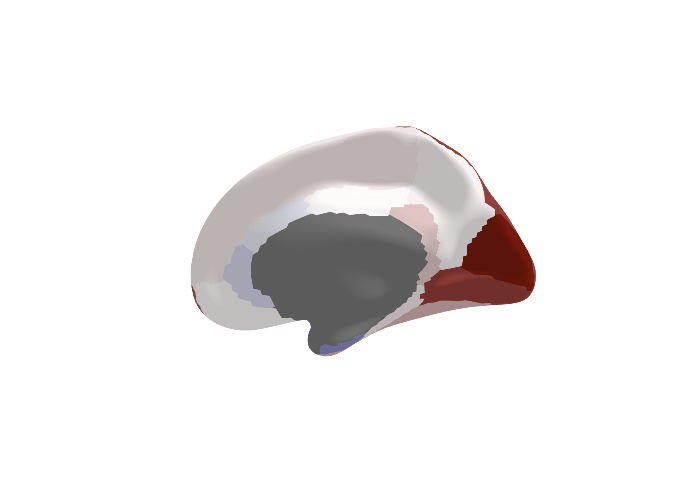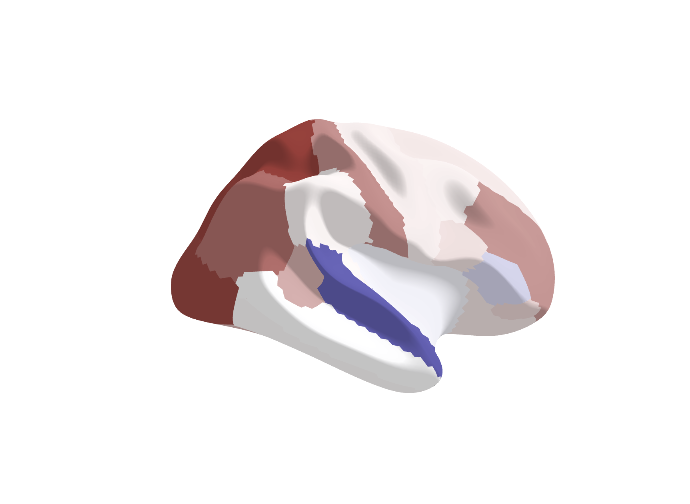 |  |
| **ICD10Dep_exclpsych** | **LifetimeMDD** | **MDDRecur** |  |
| 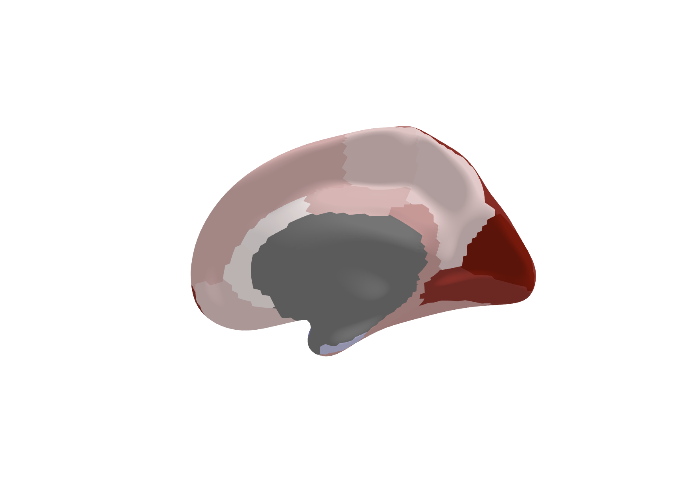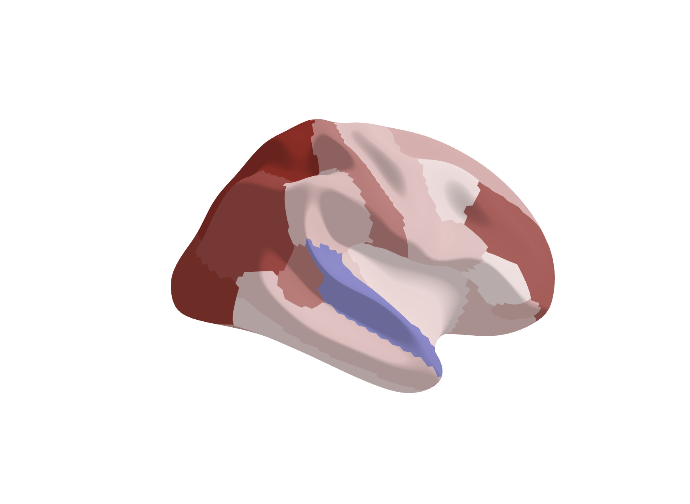 | 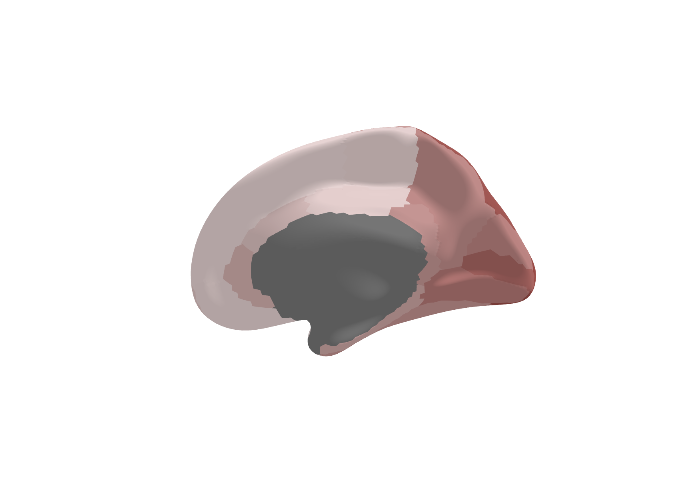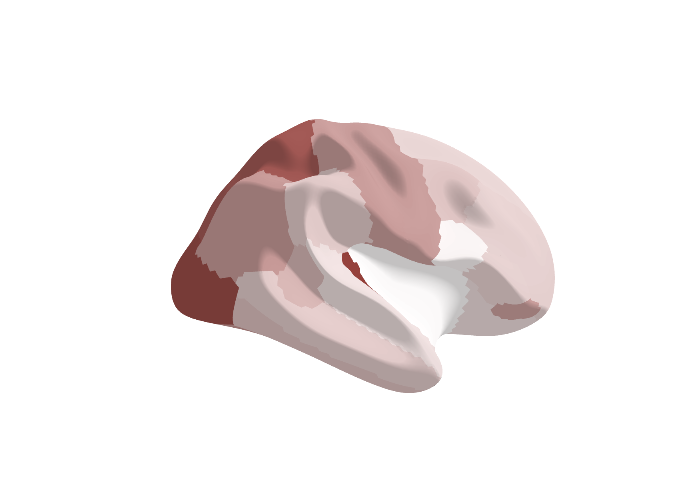 | 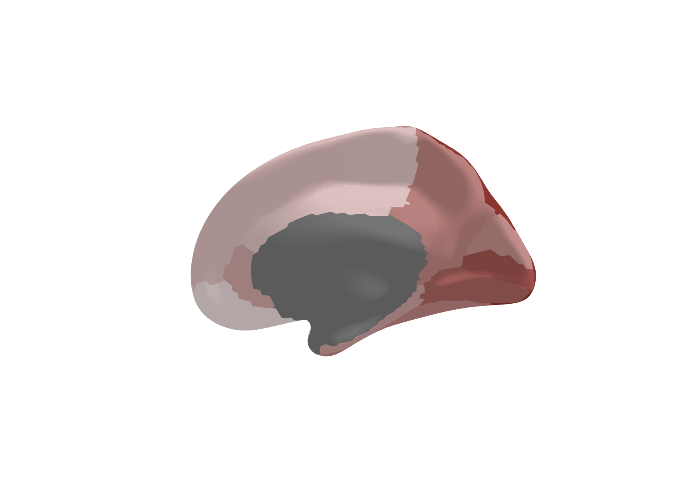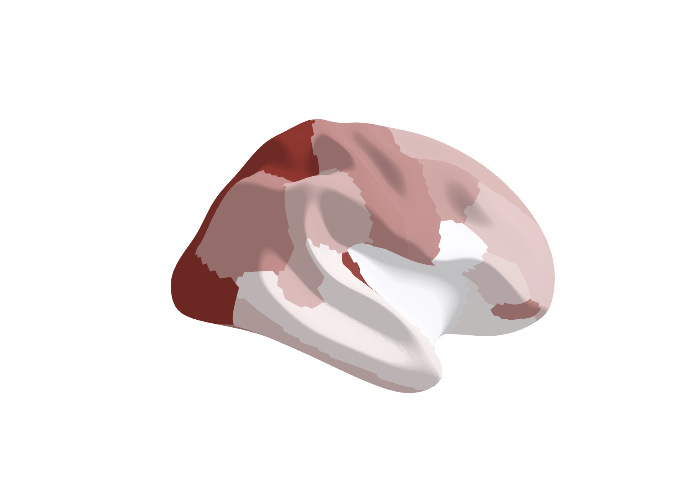 |  |

**Supplementary Figure S1 | Effect sizes across imaging-derived phenotypes and the depression spectrum**

This figure illustrates the relationship between cortical measures and depression across varying phenotyping strategies. The results are derived from multivariable logistic regression models, each including one of 256 z-score transformed neuroanatomical variables as the primary predictor, along with covariates (age, sex, intracranial volume, assessment center, and head motion), with depression status (case/control) as the outcome. This figure focuses on all four main imaging-derived phenotypes (IDP), including Fractional Anisotropy (FA), Mean Diffusivity (MD), Cortical Thickness (CT), and Surface Area (SA), ranging from strictly defined (top left) to minimally defined (bottom right). The 3D brain maps visual Odds Ratios (ORs) across each of the nine depression phenotypes. The color scheme interpretation is consistent across all measures, where cooler colors (blues) indicate regions where increased measures (FA, MD, CT, or SA) are associated with decreased odds of depression, and warmer colors (reds) indicate regions where increased measures are associated with increased odds of depression, respectively. Notably, the area in gray corresponds to the temporal pole, which was not available in data generated using Freesurfer by parcellation of the white surface using Desikan-Killiany parcellation, as part of the UK Biobank brain imaging pipeline for T1 structural brain MRI, due to known segmentation difficulties in this region.
