## Supplementary Figures for "Mapping heterogeneity in the neuroanatomical correlates of depression": Supplementary Figure S2 _ Comparison of cases and controls across the depression phenotyping spectrum.docx

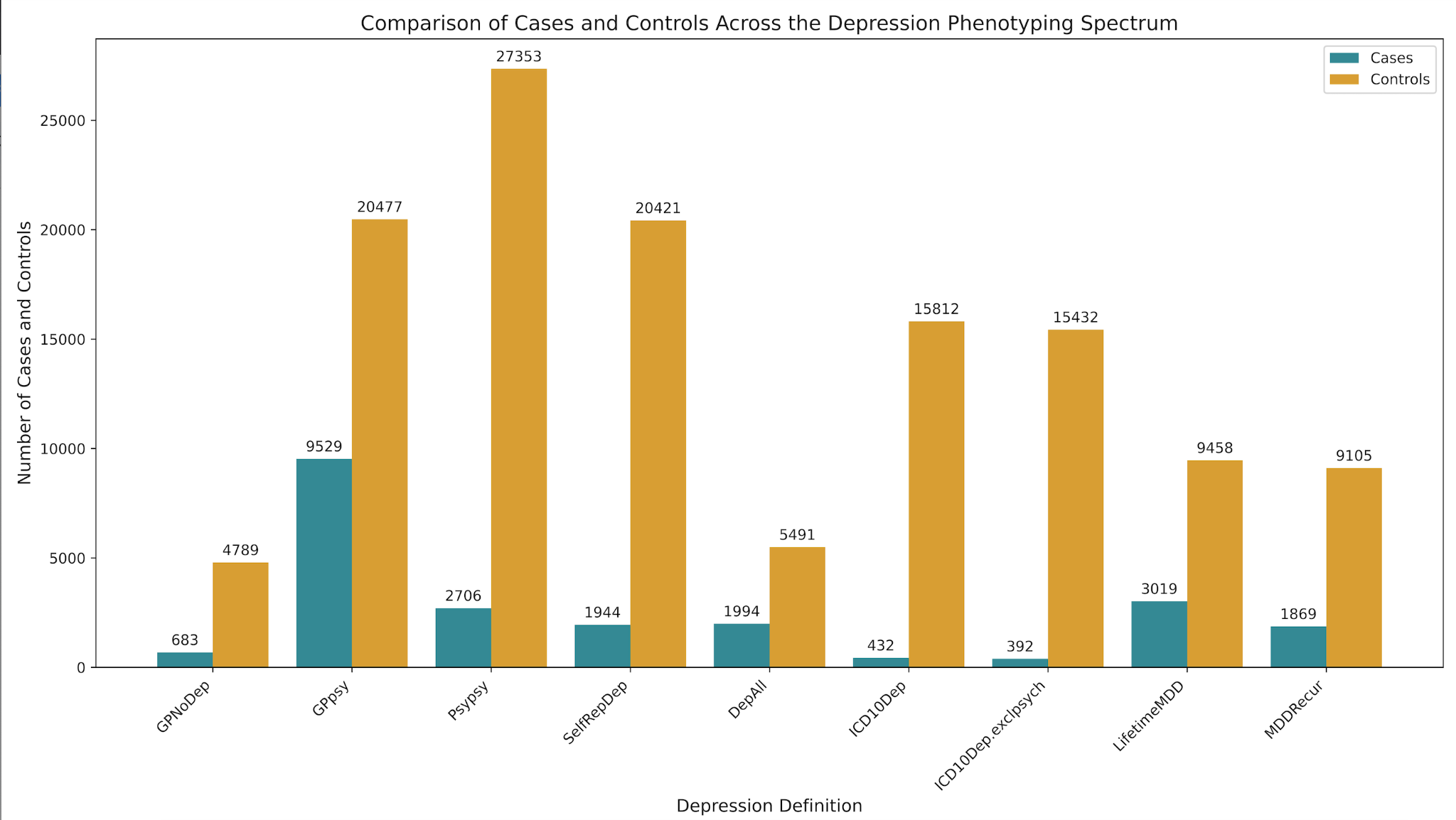

**Supplementary Figure S2 | Comparison of cases and controls across the depression phenotyping spectrum**
