## Supplementary Figures for "Mapping heterogeneity in the neuroanatomical correlates of depression": Supplementary Figure S3 _ Association between imaging-derived cortical phenotypes and depression definitions._.docx

A) Surface Area
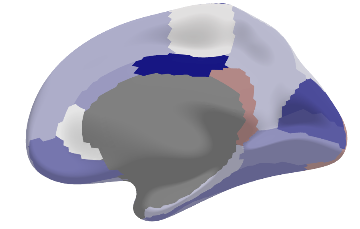

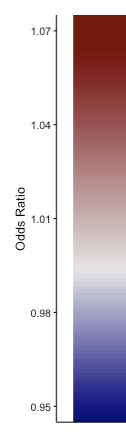

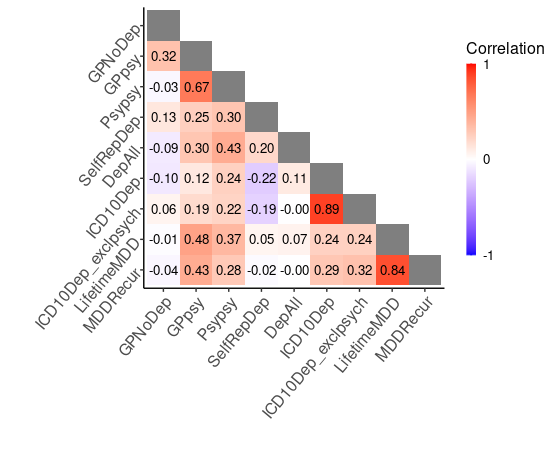

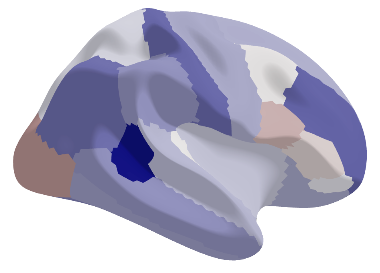

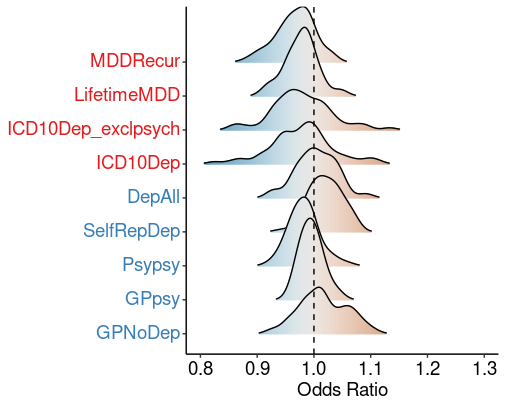

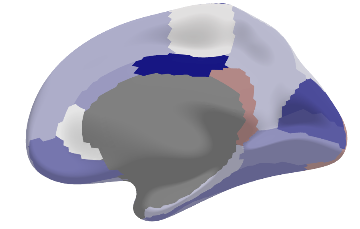

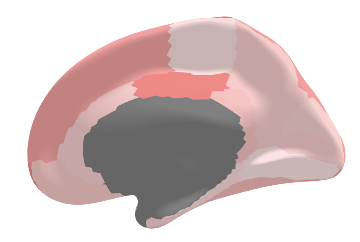

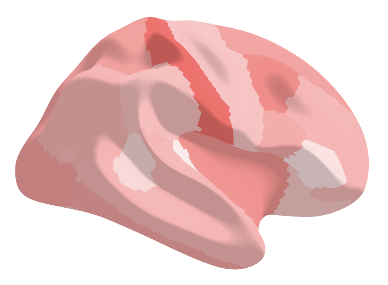

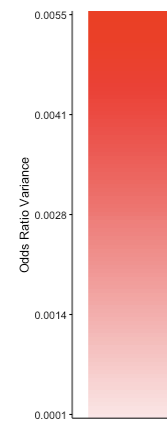

B) Cortical Thickness
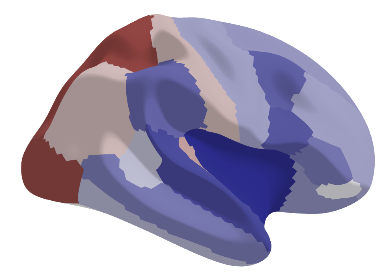

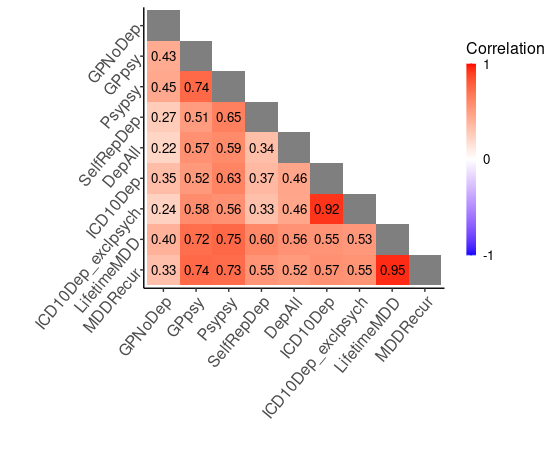

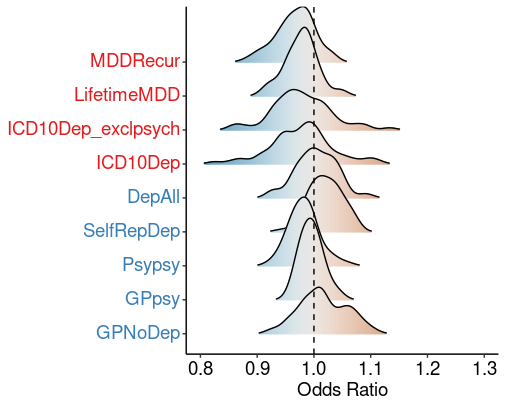

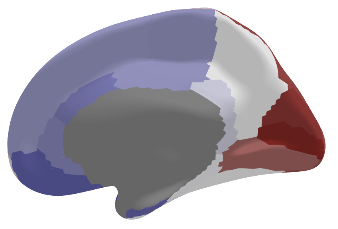

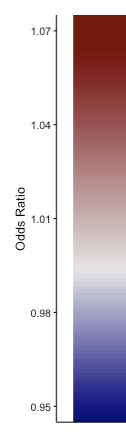

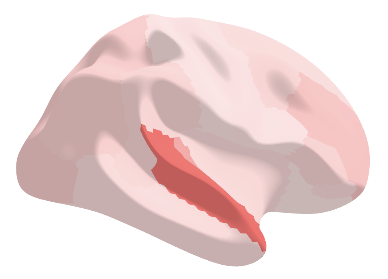

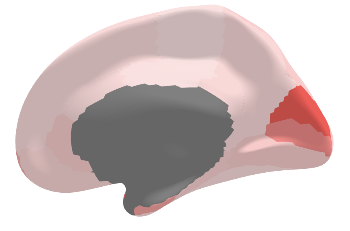

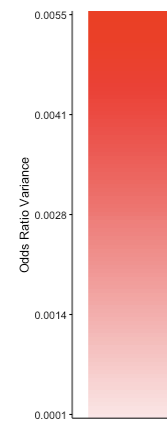

**Supplementary Figure S3 | Association between imaging-derived cortical phenotypes and depression definitions.** This figure illustrates the relationship between cortical measures and depression across varying phenotyping strategies. The results are derived from multivariable logistic regression models, each including one of 256 z-score transformed neuroanatomical variables as the primary predictor, along with covariates (age, sex, intracranial volume, assessment center, and head motion), with depression status (case/control) as the outcome. This figure focuses on two key cortical measures, namely (A) Surface Area (SA) and (B) Cortical Thickness (CT). **(A)** SA and (B) CT associations are presented through density plots, 3D brain maps, and correlation heatmaps. Density plots (left) display odds ratios (ORs) for SA and CT across depression phenotypes, ranging from strictly defined (top) to minimally defined (bottom). Deeper phenotypes are depicted in red, shallower in blue. ORs < 1 are shown in light blue, >1 in light brown. The 3D brain maps (center) visualize mean ORs (top) and standard deviations (bottom) across all nine depression definitions. For both measures, cooler colors (blues) indicate regions where increased SA or CT is associated with decreased odds of depression, whereas warmer colors (red) indicate regions where increased SA or CT is associated with increased odds of depression, respectively. Standard deviation maps highlight regions with the most variance in effects across definitions. Heatmaps (right) display Spearman's correlations of ORs across depression definitions, ranging from -1 (perfect negative correlation) to 1 (perfect positive correlation). In contrast to the white matter microstructure findings, both Surface Area and Cortical Thickness demonstrate limited associations with depression status. The density plots show narrow ranges of ORs centered close to 1, indicating minimal effect sizes across most cortical regions. The 3D brain maps reveal mostly neutral colors, suggesting weak associations between these cortical measures and depression across phenotypes. The standard deviation maps show low variability, further emphasizing the consistency of these weak associations across different depression definitions. Notably, the correlation heatmaps reveal distinct patterns between Surface Area and Cortical Thickness. These heatmaps display the correlations of odds ratios across depression definitions, providing insight into the consistency of effect patterns across phenotypes. The Surface Area heatmap displays a heterogeneous mix of correlations, with varying intensities of colors, indicating that the patterns of odds ratios vary considerably across different depression definitions. This suggests that the relationship between Surface Area and depression is sensitive to how depression is defined. In contrast, the Cortical Thickness heatmap shows predominantly darker colors, indicating higher correlations of odds ratios across definitions. This suggests that the pattern of associations between Cortical Thickness and depression is more consistent, regardless of how depression is defined. This difference highlights that while both measures show weak overall associations with depression, the pattern of Cortical Thickness associations remains more stable across phenotyping strategies compared to Surface Area associations.
