## Supplementary Figures for "Mapping heterogeneity in the neuroanatomical correlates of depression": Supplementary Figure S4 _ Relationships between brain-psychopathology associations in UK Biobank and the ENIGMA BD and SCZ consortia in fractional anisotropy_.docx

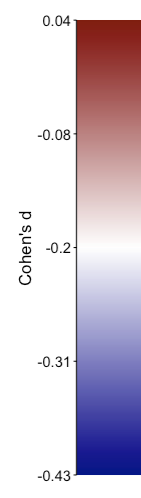

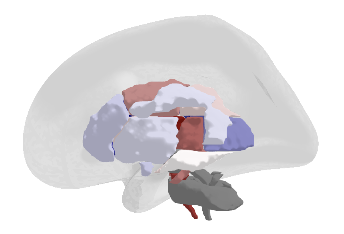

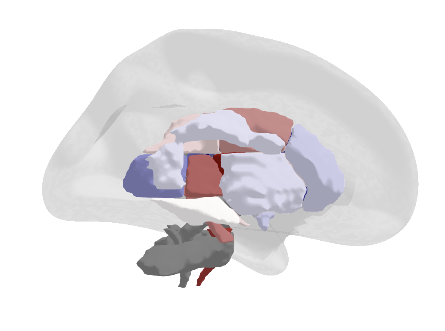

**Bipolar Disorder**

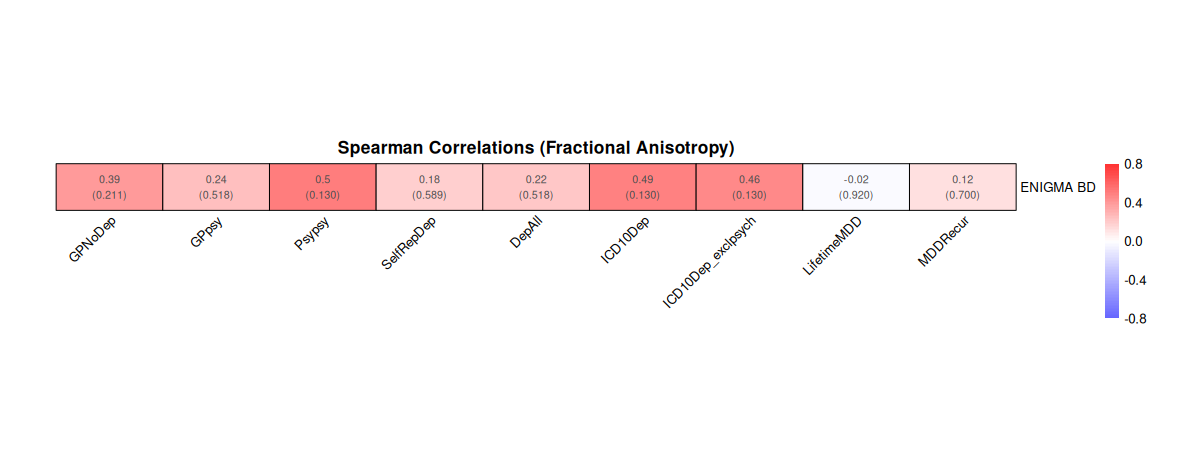

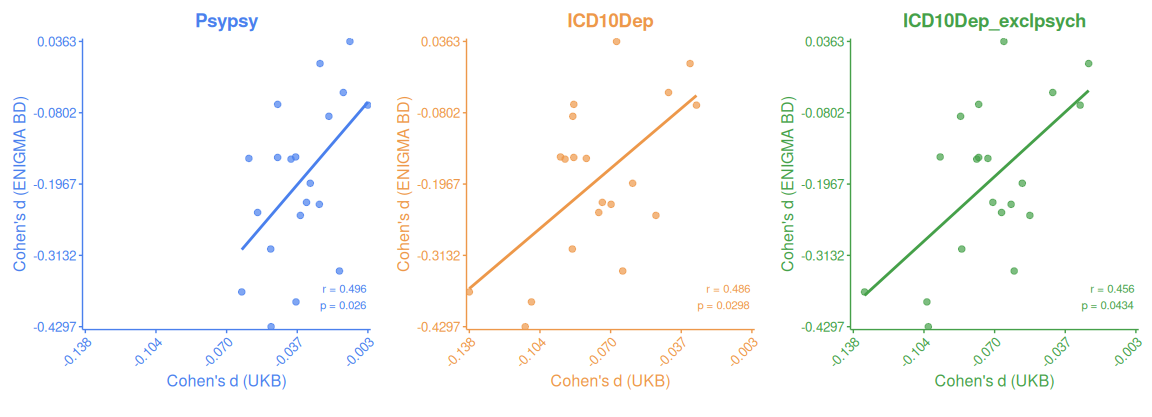

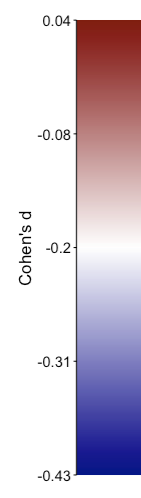

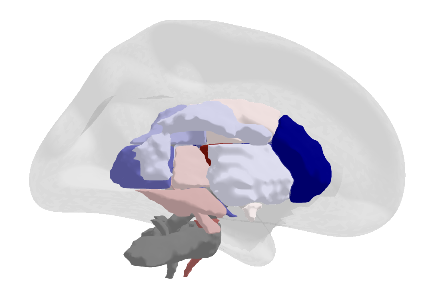

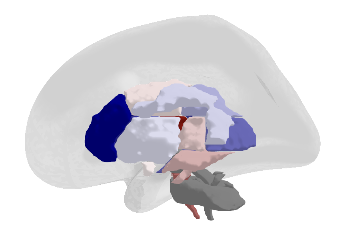

**Schizophrenia**

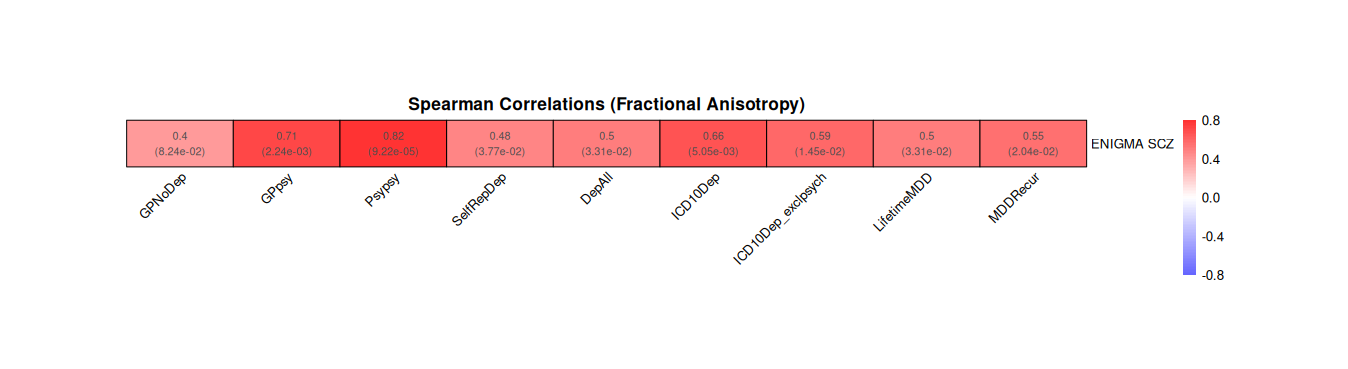

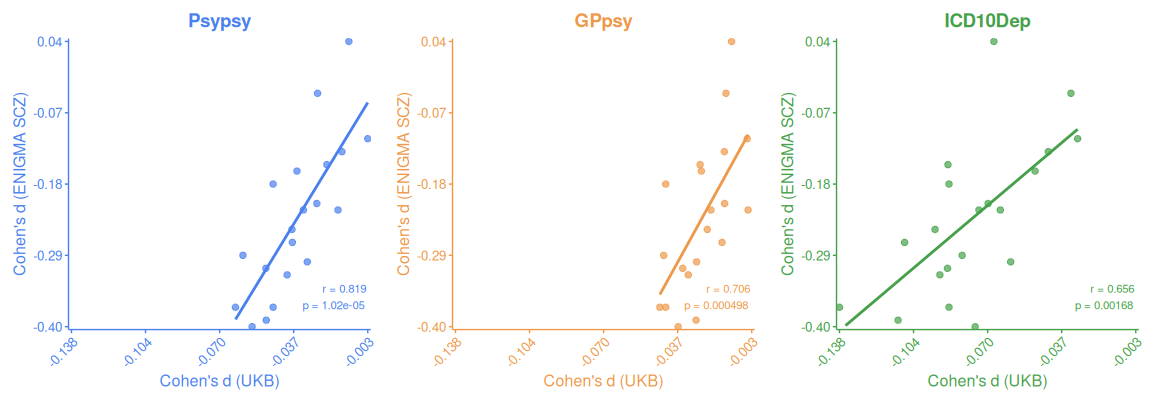

**Supplementary Figure S4 | Relationships between brain-psychopathology associations in UK Biobank and the ENIGMA BD and SCZ consortia in fractional anisotropy.** This figure examines convergence between white matter microstructure alterations, specifically fractional anisotropy (FA), in psychotic disorders and depression phenotypes. The analysis compares effect sizes (Cohen's d) between ENIGMA consortium FA findings and UK Biobank depression phenotypes across white matter tracts, as FA was the only diffusion measure available across both meta-analyses. For Bipolar Disorder (**BD**), the brain maps display ENIGMA BD working group effect size estimates, showing heterogeneous alterations with both decreased (blue) and increased (red) FA values relative to controls (d=-0.43 to d=0.04). The largest FA decreases were observed in the body of corpus callosum (d=-0.43), cingulum (d=-0.39), genu (d=-0.37), splenium (d=-0.34), and posterior thalamic radiation (d=-0.30). Correlations between ENIGMA BD FA effects and UKBB depression phenotypes were modest, with highest associations in Psypsy (r=0.50, p=0.026), ICD10Dep (r=0.49, p=0.030), and ICD10Dep excluding psychiatric comorbidities (r=0.46, p=0.043). For Schizophrenia (**SCZ**), FA maps showed consistently lower values in cases versus controls, with largest differences in anterior corona radiata (d=-0.40), corpus callosum (d=-0.40), body of corpus callosum (d=-0.39), anterior limb of internal capsule (d=-0.37), and genu (d=-0.37). ENIGMA SCZ FA effects showed stronger correlations with depression phenotypes compared to BD, particularly for Psypsy (r=0.82, p=1.02e-05), GPpsy (r=0.71, p=0.000498), and ICD10Dep (r=0.66, p=0.00168). For both disorders, strongest correlations emerged with phenotypes capturing psychiatric care-seeking behavior and EMR-based phenotypes rather than broader or more stringent depression definitions.
