## Supplementary Figures for "Mapping heterogeneity in the neuroanatomical correlates of depression": Supplementary Figure S5 _ Relationships between brain-psychopathology associations in UK Biobank and the ENIGMA MDD consortium in gray matter morphometry.docx

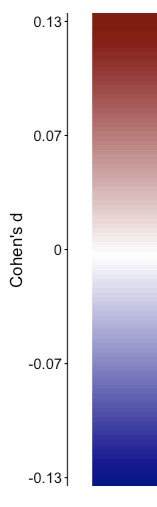

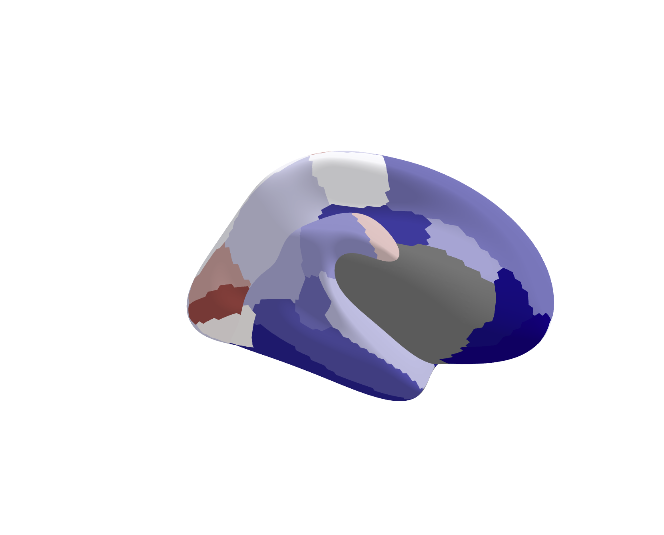

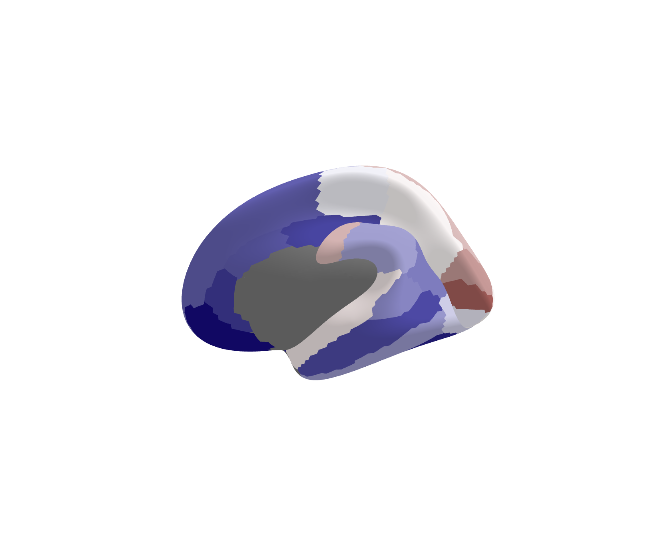

**Cortical Thickness**
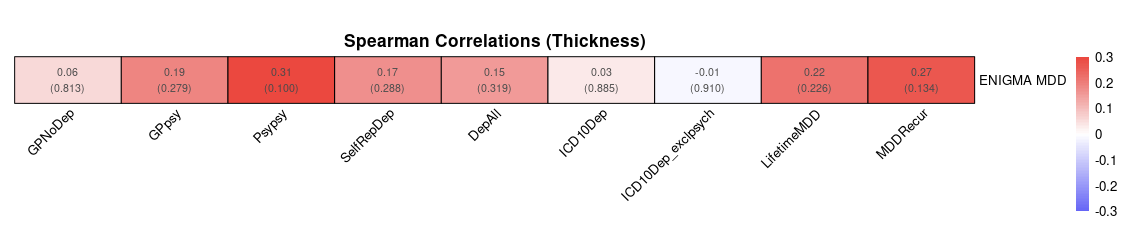

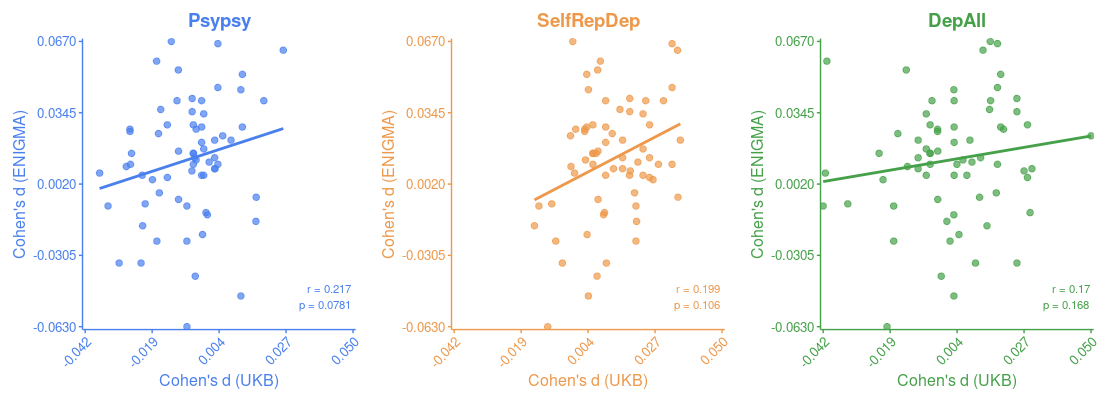

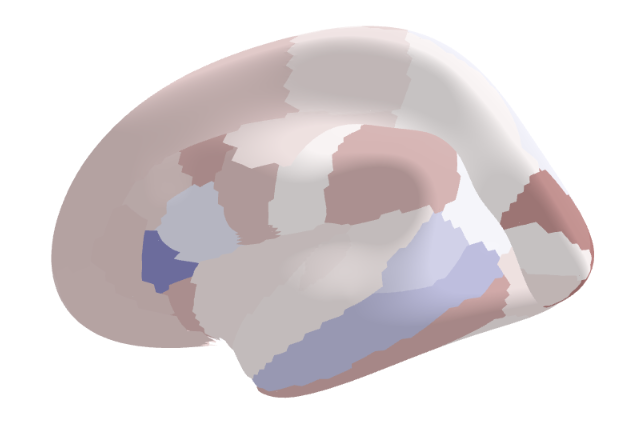

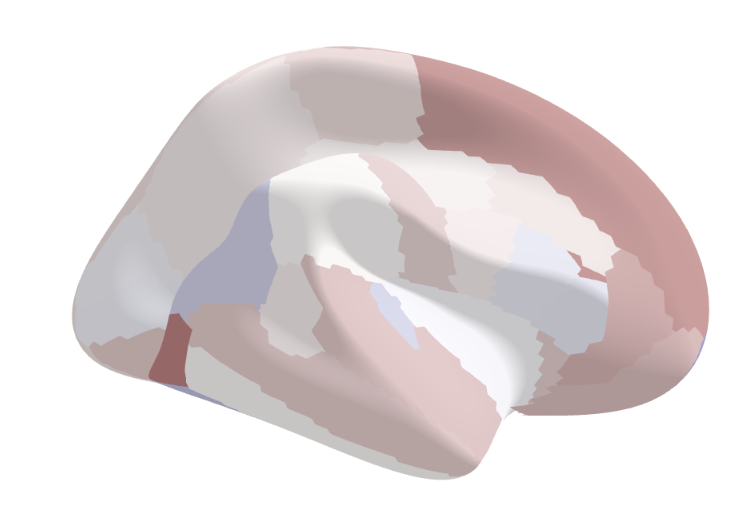

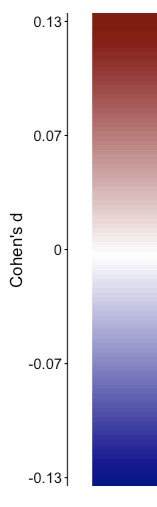

**Surface Area**
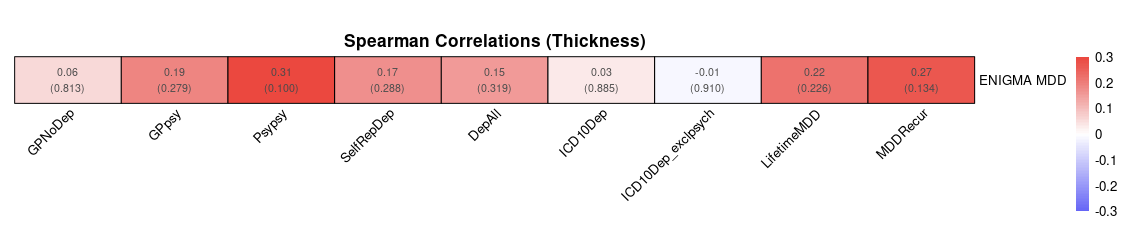

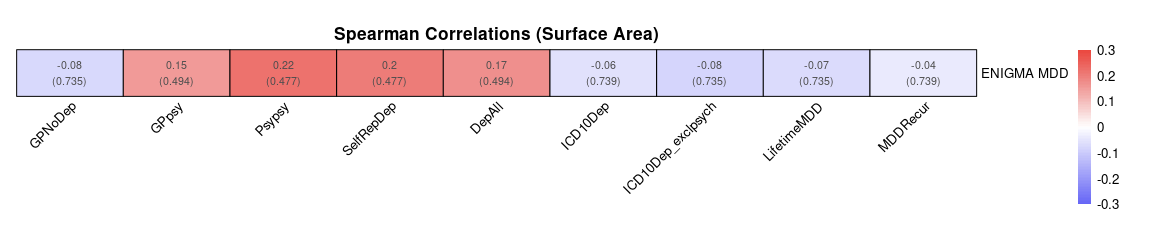

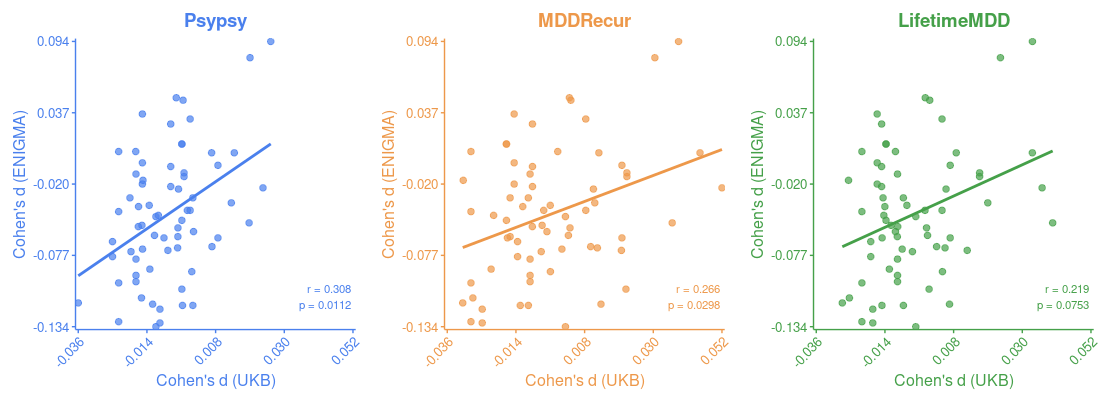

**Supplementary Figure S5 | Relationships between brain-psychopathology associations in UK Biobank and ENIGMA MDD in gray matter morphometry.** This figure compares cortical thickness (CT) and surface area (SA) effect sizes between the UK Biobank (UKBB) depression phenotypes and effect size estimates from the most recent ENIGMA MDD Working Group meta-analysis. For both CT and SA, the figure presents heatmap vectors of Spearman’s rank correlation between ENIGMA MDD and UKBB results across depression phenotypes, with no correlations reaching statistical significance. The central brain maps visualize the ENIGMA MDD meta-analysis effect size estimates (Cohen’s d) for each cortical region. Scatter plots display Cohen’s d values from ENIGMA MDD versus UKBB for three phenotypes showing highest nominal correlations. For CT, these phenotypes are Psypsy (r = 0.217), MDDRecur (r = 0.199), and LifetimeMDD (r = 0.17), while for SA they are Psypsy (r = 0.308), MDDRecur (r = 0.266), and LifetimeMDD (r = 0.219). Each point represents a cortical region. The modest correlation coefficients and absence of significant concordance between UKBB and ENIGMA MDD findings suggests heterogeneity in brain-psychopathology associations across cohorts and analytic approaches.
