## Supplementary Figures for "Mapping heterogeneity in the neuroanatomical correlates of depression": Supplementary Figure S6 _ Relationships between brain-psychopathology associations in UK Biobank and the ENIGMA BD consortium in gray matter morphometry.docx

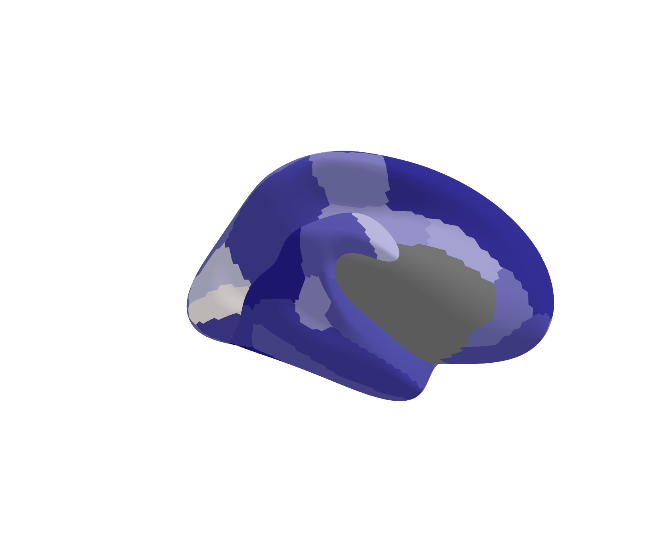

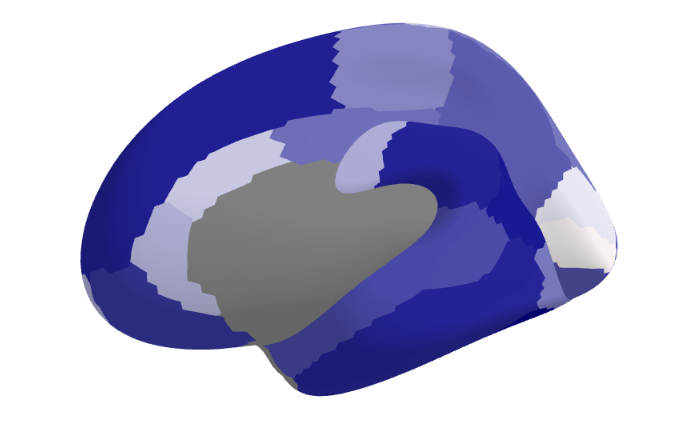

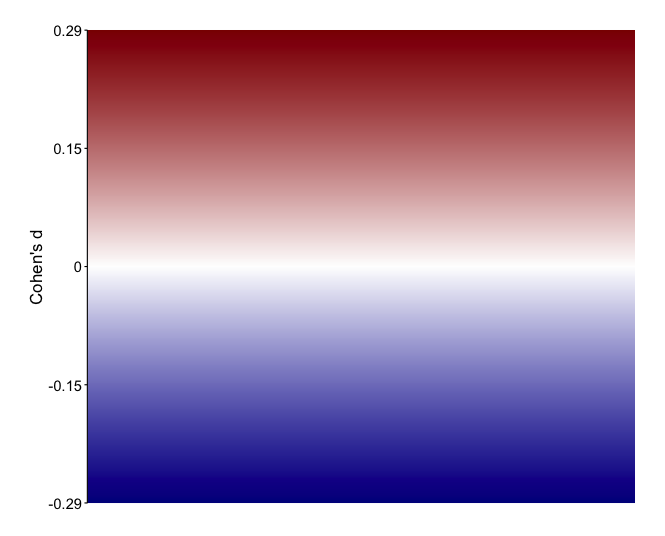

A)

**Cortical Thickness**
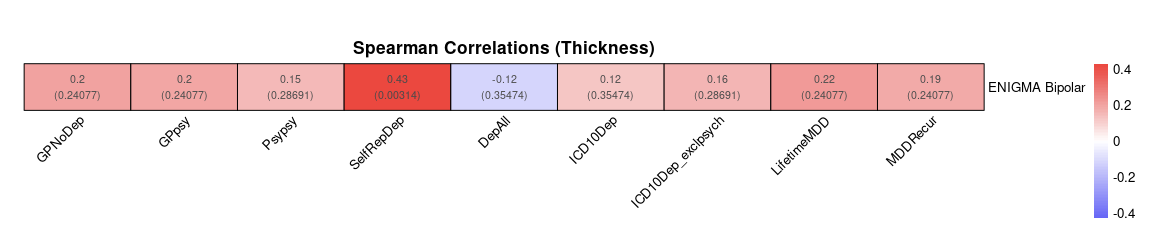

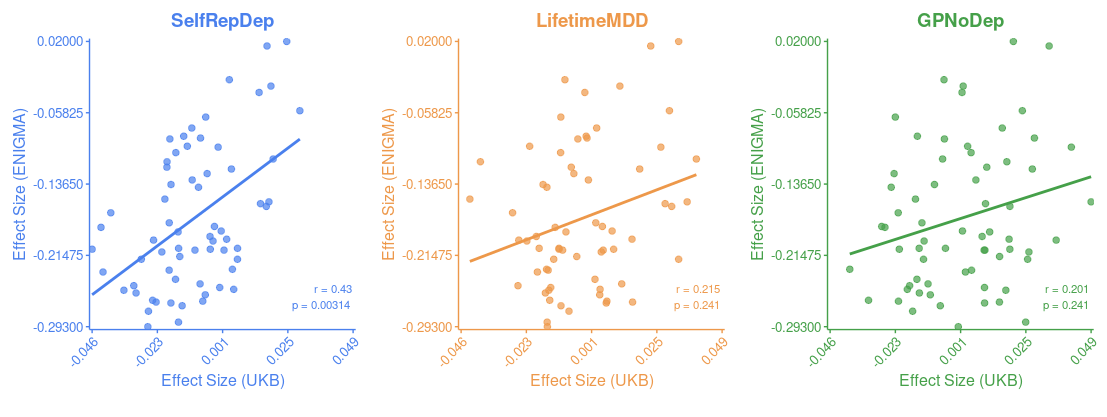

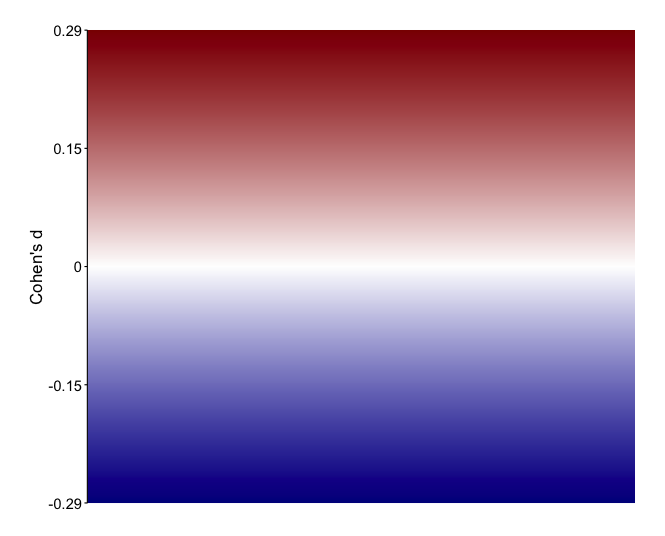

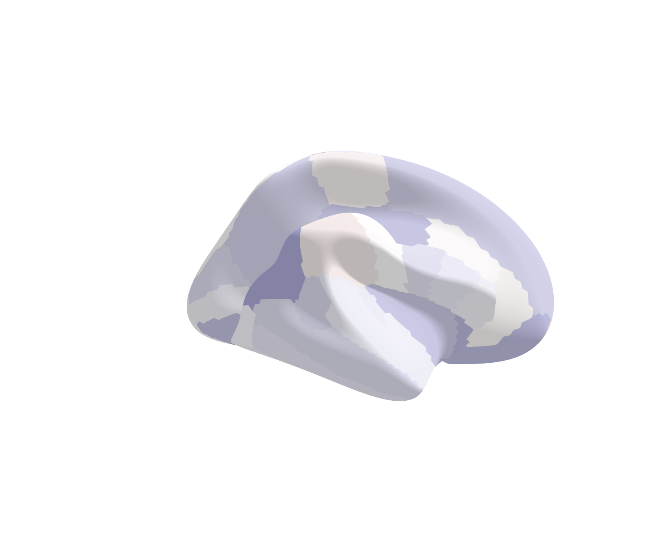

B)
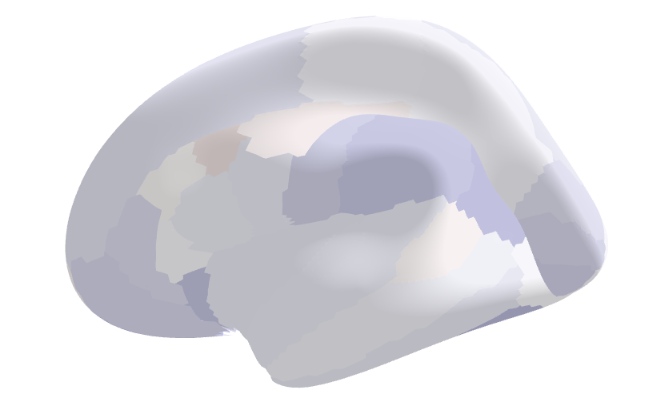

**Surface Area**

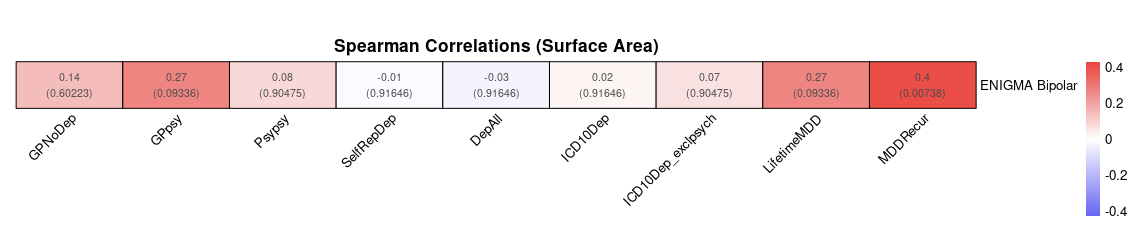

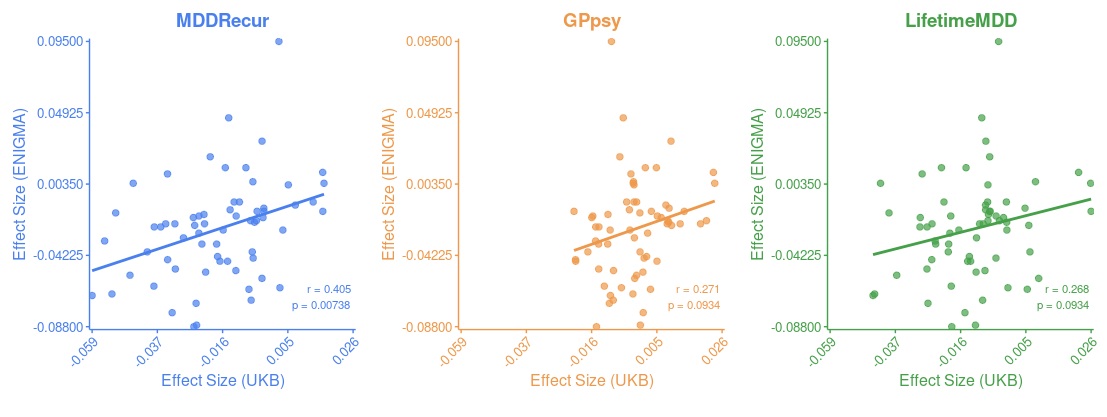

**Supplementary Figure S6 | Relationships between brain-psychopathology associations in UK Biobank and the ENIGMA BD consortium in gray matter morphometry.** This figure illustrates the convergence between gray matter morphology (specifically surface area (SA) and cortical thickness (CT)) in Bipolar Disorder (BD) using meta-analytic effect size estimates (reported in Cohen’s d) from the ENIGMA Bipolar working group, and depression ascertained across various phenotyping strategies in the UKBB. The brain maps display ENIGMA BD working group effect size estimates, where cooler colors (blues) indicate regions where increased measures are associated with decreased odds of BD, while warmer colors (red) indicate regions where increased measures are associated with increased odds of BD. These maps reveal widespread lower CT in BD cases compared to controls, while SA differences between BD cases and controls were modest to negligible. When assessing associations between ENIGMA BD meta-analytic effect size estimates and findings across UKBB depression phenotypes using Spearman correlations, most phenotypes did not show statistically significant associations. The top three depression phenotypes demonstrated low to moderate correlations ranging from r = 0.201-0.43 in CT, and r = 0.268-0.405 in SA, respectively.
