## Supplementary Figures for "Mapping heterogeneity in the neuroanatomical correlates of depression": Supplementary Figure S7 _ Relationships between brain-psychopathology associations in UK Biobank and the ENIGMA SCZ consortium in gray matter morphometry.docx

A)
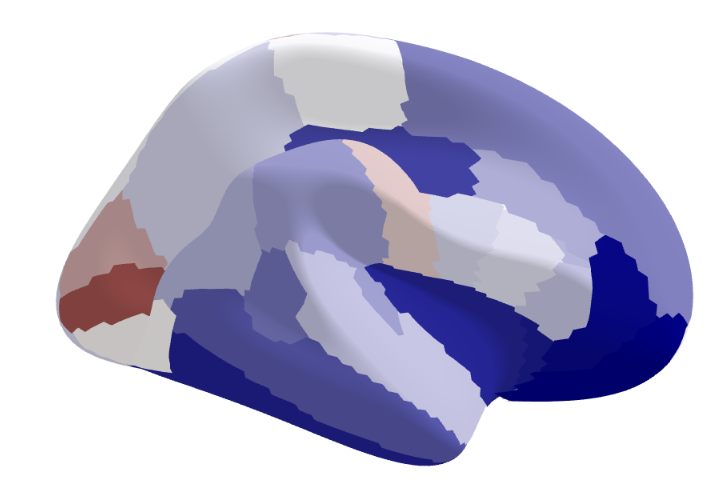

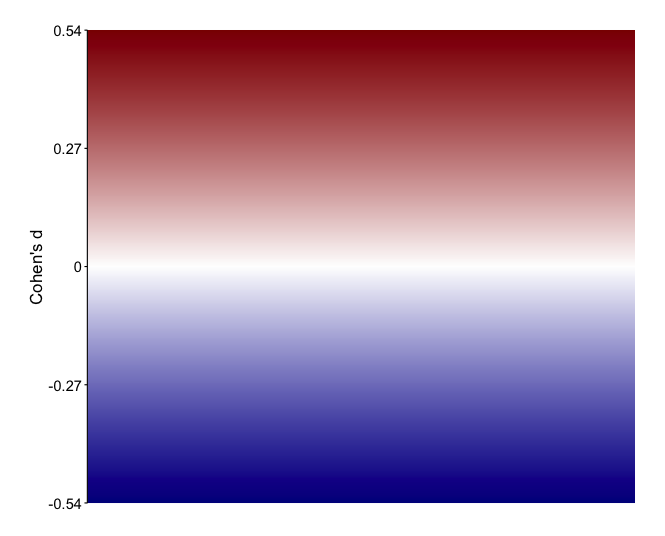

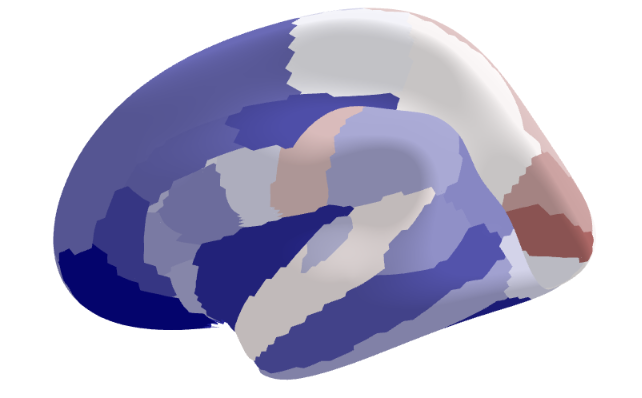

**Fractional Anisotropy**

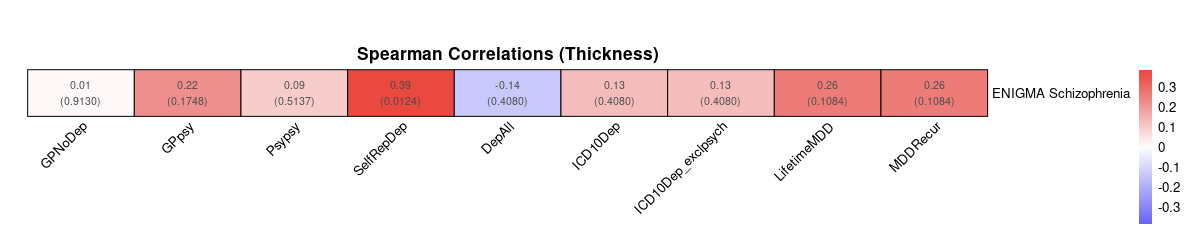

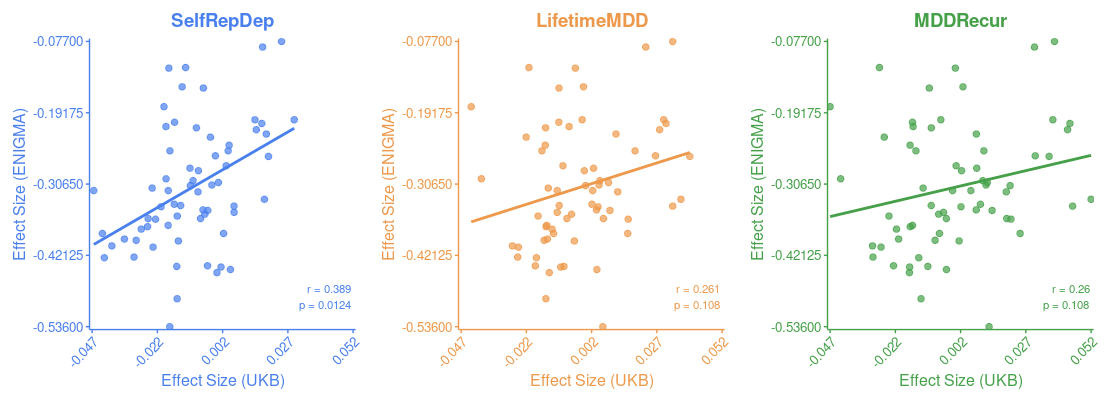

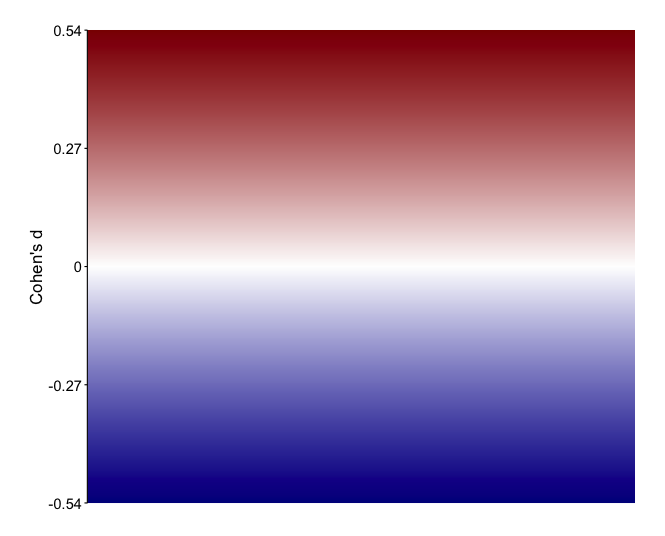

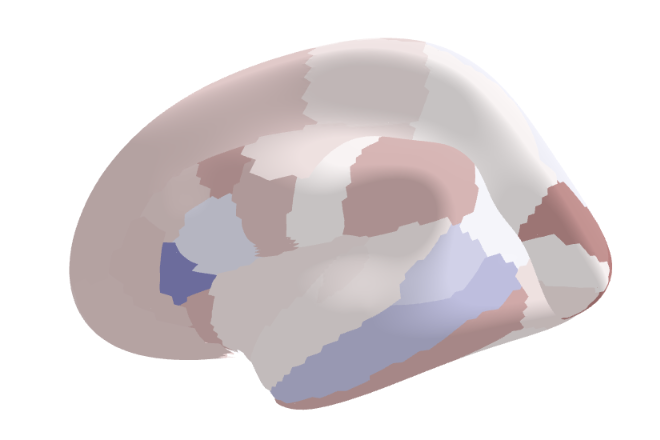

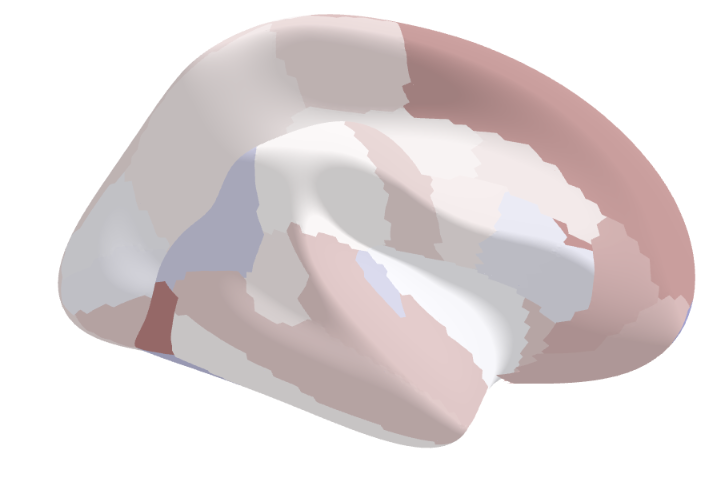

B)	**Mean Diffusivity**

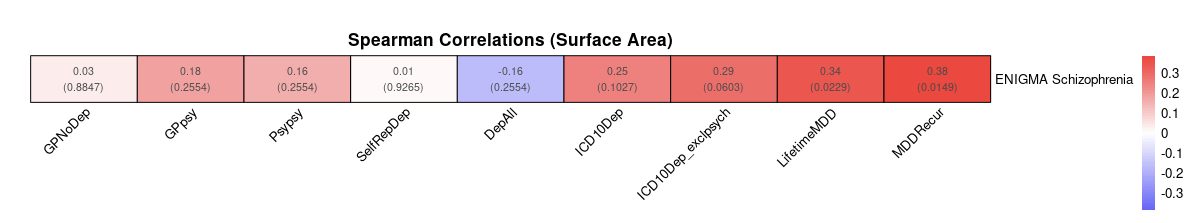

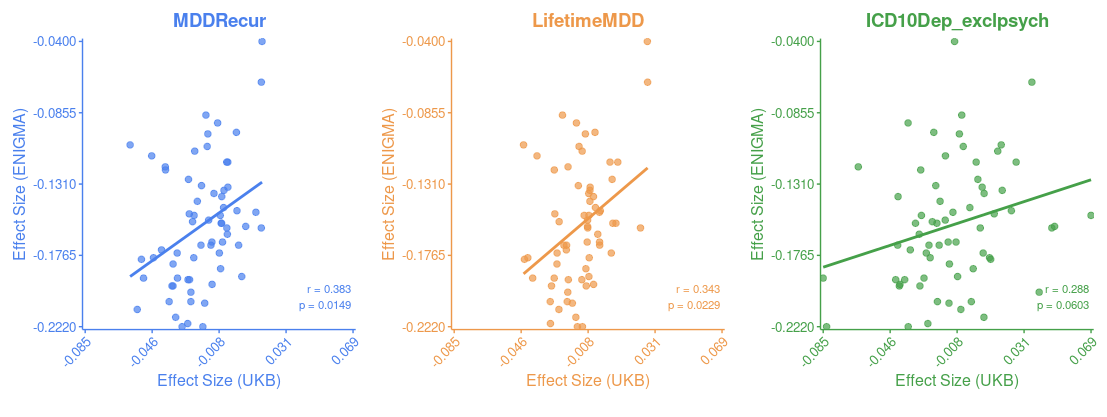

**Supplementary Figure S7 | Relationships between brain-psychopathology associations in UK Biobank and the ENIGMA SCZ consortium in gray matter morphometry.** This figure examines the convergence between gray matter morphology (specifically surface area (SA) and cortical thickness (CT)) in Schizophrenia (SCZ) using meta-analytic effect size estimates (Cohen’s d) from the ENIGMA Schizophrenia working group, and depression ascertained across various phenotyping strategies in the UKBB. The brain maps display ENIGMA SCZ working group effect size estimates, where cooler colors (blues) indicate regions where increased measures are associated with decreased odds of SCZ, while warmer colors (red) indicate regions where increased measures are associated with increased odds of SCZ. For CT **(A)**, these maps reveal lower CT in SCZ cases in occipital regions. For SA **(B)**, the pattern was reversed in frontal regions, with higher SA in SCZ cases compared to controls, though these standardized differences were more modest. When assessing associations between ENIGMA SCZ meta-analytic effect size estimates and findings across UKBB depression phenotypes using Spearman correlation, no phenotypes showed statistically significant associations. The top three depression phenotypes demonstrated low to moderate correlations ranging from r = 0.260-0.389 in CT, and r = 0.288-0.383 in SA, respectively.
