## Supplementary Figures for "Mapping heterogeneity in the neuroanatomical correlates of depression": Supplementary Figure S8 _ Total feature importance ranking across shallow and deep depression models._.docx

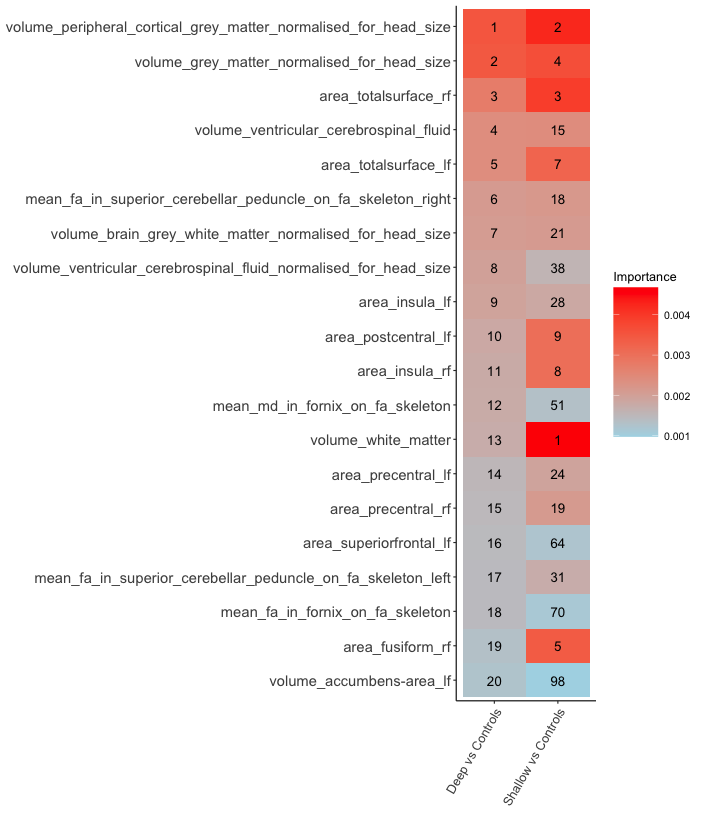

**Supplementary Figure S8 | Total feature importance ranking across shallow and deep depression models.** This figure illustrates the relative importance of the top 20 neuroimaging features in discriminating deep depression from controls, and shallow depression from controls using random forest classifiers using all 256 neuroimaging features. Feature importance was calculated using permutation-based methods implemented in the Ranger package in R, following Altmann et al.’s (2010) approach. For each feature, the decrease in out-of-bag (OOB) prediction accuracy after random permutation of the feature’s values was measured, with larger decreases indicating greater feature importance. Warmer colors (red) indicate higher importance, while cooler colors (blue) represent lower importance. Overall, the deep depression model prioritizes volume measurements, with normalized volume of peripheral cortical gray matter and normalized gray matter volume ranking 1st and 2nd respectively (2nd and 4th in shallow model). Five of the top eight features in the deep depression model are volume-related. In contrast, the shallow depression model shows different prioritization, with volume of white matter as the top feature (ranked 13th in the deep model). The area measurements show varying importance between models - total surface area right ranks 3rd in both models, while total surface area left ranks 5th in deep but 7th in shallow. Some features show marked differences in importance - volume of ventricular cerebrospinal fluid ranks 4th in deep but 15th in shallow depression, and area fusiform right ranks 19th in deep but 5th in shallow. The volume of accumbens area left shows the most dramatic difference, ranking 20th in deep but 98th in shallow depression.
