## Supplementary Figures for "Mapping heterogeneity in the neuroanatomical correlates of depression": Supplementary Figure S9 _ Comparison of feature importance rankings between multi-class and deep vs shallow depression classifiers_.docx

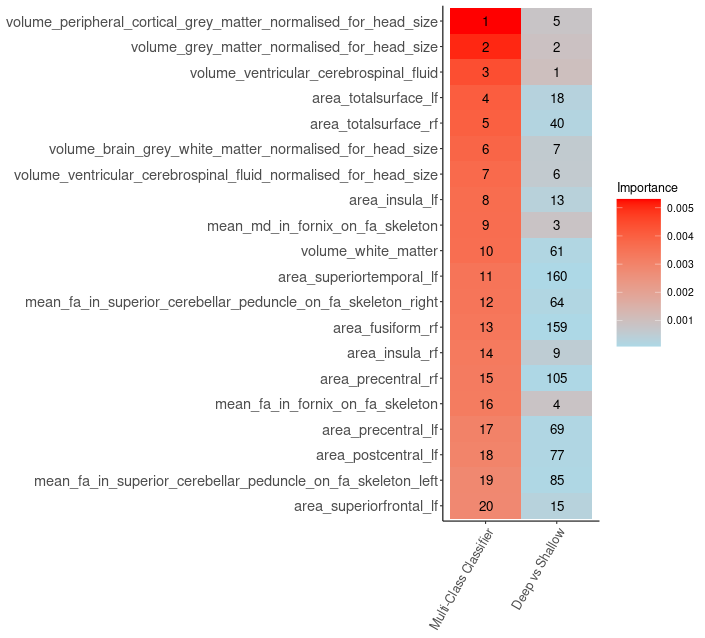

**Supplementary Figure S9 | Comparison of feature importance rankings between multi-class and deep vs shallow depression classifiers.** This figure compares the relative importance of neuroimaging features between two random forest models: a multi-class classifier distinguishing between controls, shallow depression, and deep depression (left column), and a binary classifier differentiating deep from shallow depression (right column). Feature importance was calculated using permutation-based methods, where larger decreases in model accuracy after feature permutation indicates greater importance, represented by warmer colors in the heatmap. The visualization reveals both consistencies and divergences is how these models prioritize neuroanatomical features. Volume-based measurements demonstrate high relative importance in the multi-class classifier, with peripheral cortical gray matter, total gray matter, and ventricular cerebrospinal fluid volumes occupying the top three positions. Surface area measurements show notably different rankings between models - for example, total surface area (left) ranks 4th in the multi-class classifier, but 18th in the deep vs shallow comparison. White matter measurements, including both volumetric and microstructural (mean FA/MD) features, appear consistently important but with varying rankings. This pattern suggests that while volume-based measurements are strongly associated with distinguishing depression from healthy controls, the differentiation between ‘shallower’ and ‘deeper’ depression phenotypes relies on a broader range of neuroanatomical characteristics.
