## Supplementary Figures for "Mapping heterogeneity in the neuroanatomical correlates of depression": Supplementary Figure S10 _ Neuroimaging-based machine learning pipeline and feature transfer framework for depression classification.docx

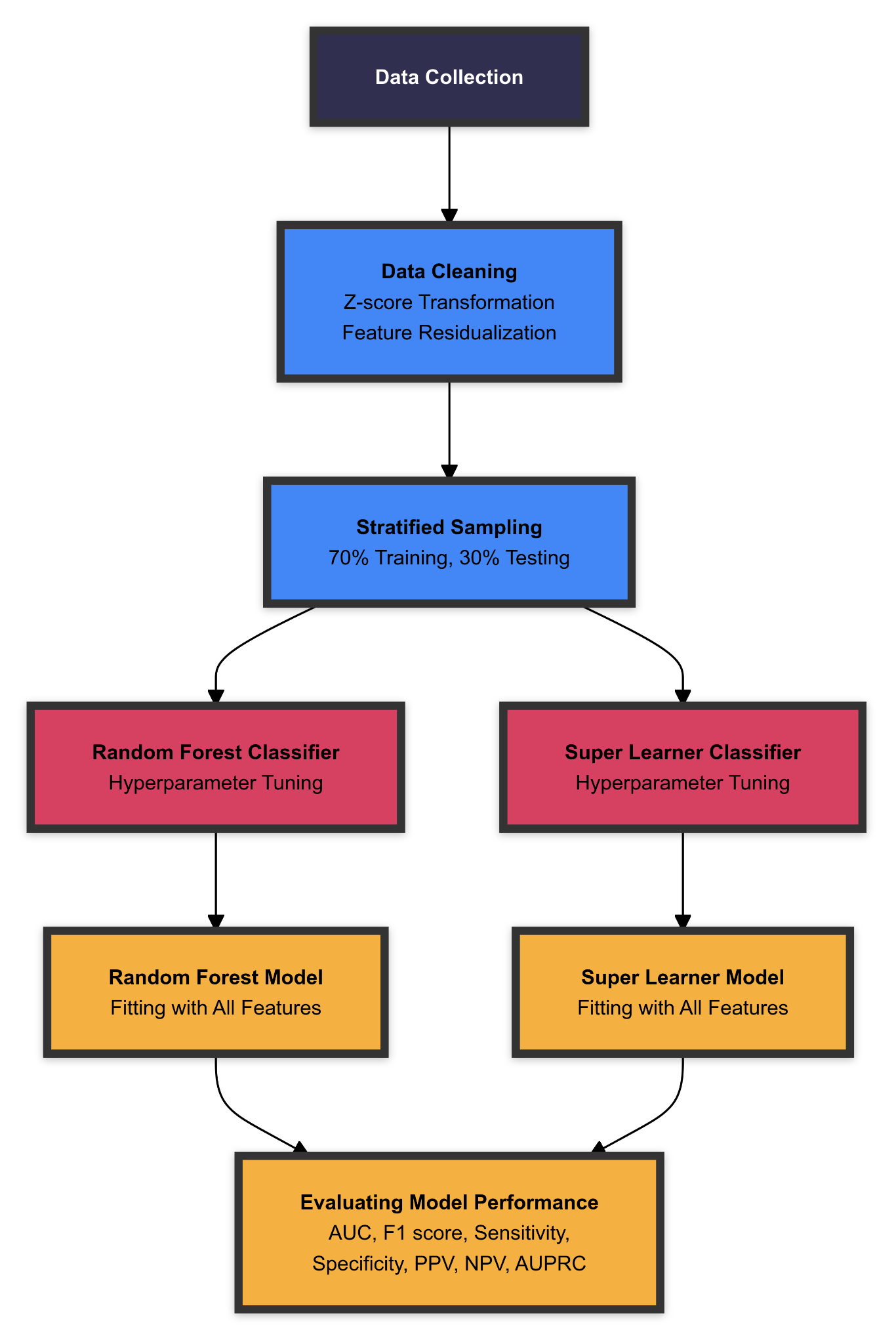

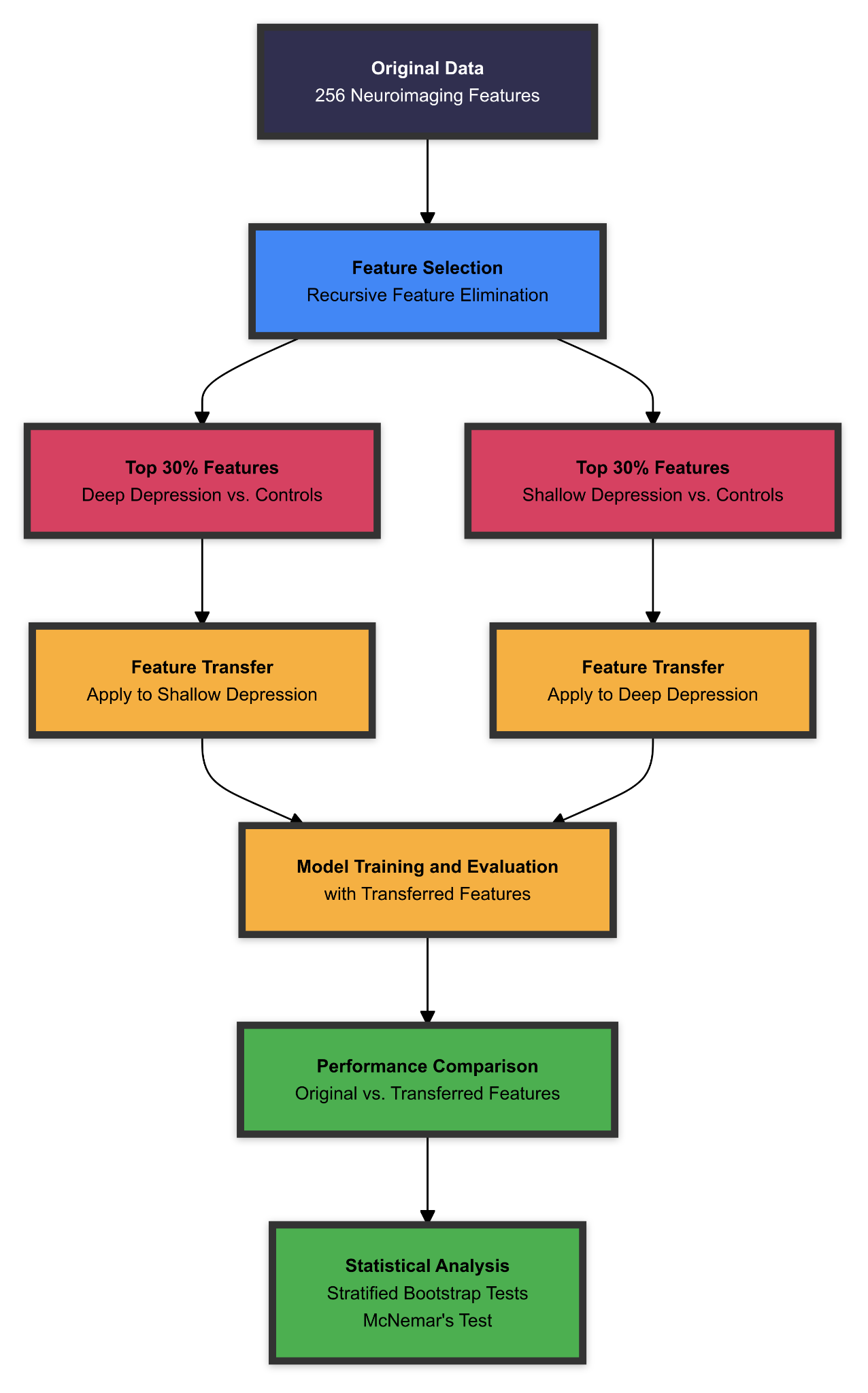

**Supplementary Figure S10 | Neuroimaging-based machine learning pipeline and feature transfer framework for depression classification.** An illustration of the machine learning pipeline used to investigate the predictive power and generalizability of neuroanatomical features across various depression phenotypes. **a)** The primary machine learning pipeline begins with using preprocessed neuroanatomical data collected by the UK Biobank (detailed in Supplementary Table 1). After data cleaning through z-score transformation and feature residualization, stratified sampling splits the data into training (70%) and testing (30%) sets. Two classifiers are implemented - Random Forest and Super Learner - both using Bayesian Optimization for hyperparameter tuning. Model performance is evaluated using multiple metrics, including AUC, F1 Score (harmonic mean of precision and recall), sensitivity, specificity, PPV, NPV, and AUPRC. **b)** The feature transfer framework assesses neuroanatomical feature generalizability across depression phenotypes. From 256 neuroimaging features, recursive feature elimination selects the top 30% (76) features for both deep and shallow depression versus controls models. These features are then applied reciprocally: deep depression features classify shallow depression and vice versa. Specifically, the 76 features identified from the shallow versus controls classifier are used to train a new deep versus controls model, while the 76 features from the deep versus controls classifier are used to train a new shallow versus controls model. Performance comparisons between original and transferred feature models undergo statistical testing via stratified bootstrap and McNemar’s tests. This approach evaluates both predictive performance metrics and feature importance rankings across depression phenotypes, identifying which neuroanatomical correlates best predict different phenotypes and their cross-phenotype generalizability.
