## Supplementary Figures for "Mapping heterogeneity in the neuroanatomical correlates of depression": Supplementary Figure S11 _ Feature transfer framework using top shallow features and importance comparison across depression models_.docx

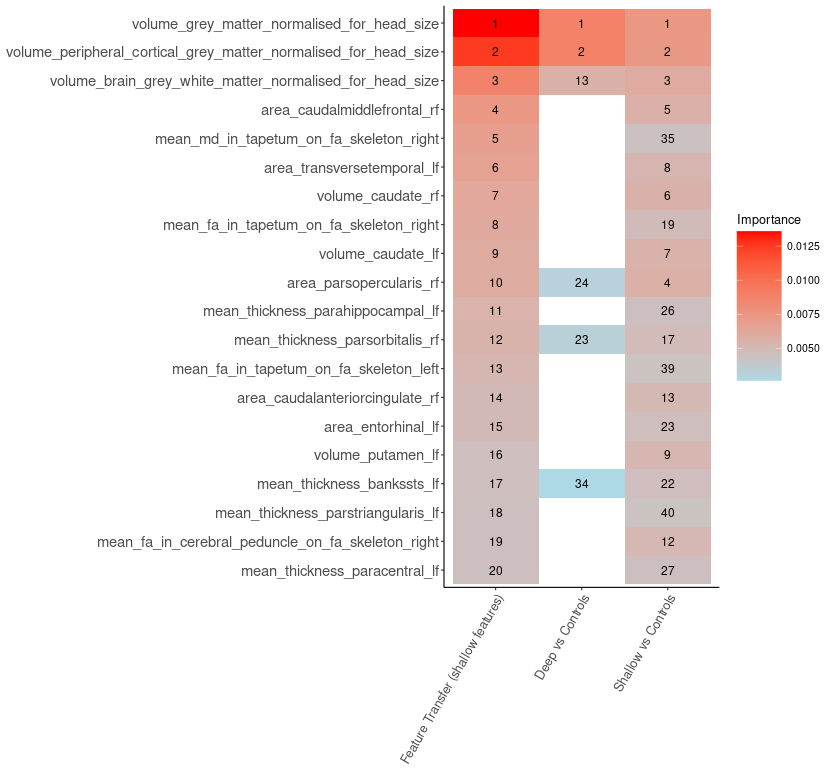

**Supplementary Figure S11 | Feature transfer framework using top shallow features and importance comparison across depression models.** This figure illustrates the feature transfer approach comparing three models: shallow depression vs controls, deep depression vs controls, and a feature transfer model that applies shallow depression-derived features to deep depression classification. The analysis focuses on the top 30% of features (76 total) identified through recursive feature elimination (RFE) in the training set. Feature importance was calculated using permutation-based methods in the Ranger R package. This approach measures each feature’s importance by first computing the model’s prediction accuracy on out-of-bag samples, then randomly permutes the feature’s values to break any association with the outcome. The resulting decrease in prediction accuracy after permutation, averaged across all trees in the forest, represents the feature’s importance - where larger decreases indicate greater importance. The heatmap visualizes feature rankings and relative importance (red indicating higher importance, blue lower) across all three models. While the shallow depression and feature transfer models share the same feature set, the deep depression model shows limited overlap, evidenced by numerous blank spaces indicating features absent from its top 30% (76 features in total). Notable ranking differences exist even between models sharing feature sets - for example, the 5th-ranked feature in the feature transfer model (mean md in tapetum on fa skeleton right) ranks 35th in the shallow depression model, and the 8th-ranked feature (mean fa in tapetum on fa skeleton right) ranks 19th. However, the top two features maintain consistent importance across all models.
