## Supplementary Figures for "Mapping heterogeneity in the neuroanatomical correlates of depression": Supplementary Figure S12 _ Deep feature transfer framework and importance comparison across depression models_.docx

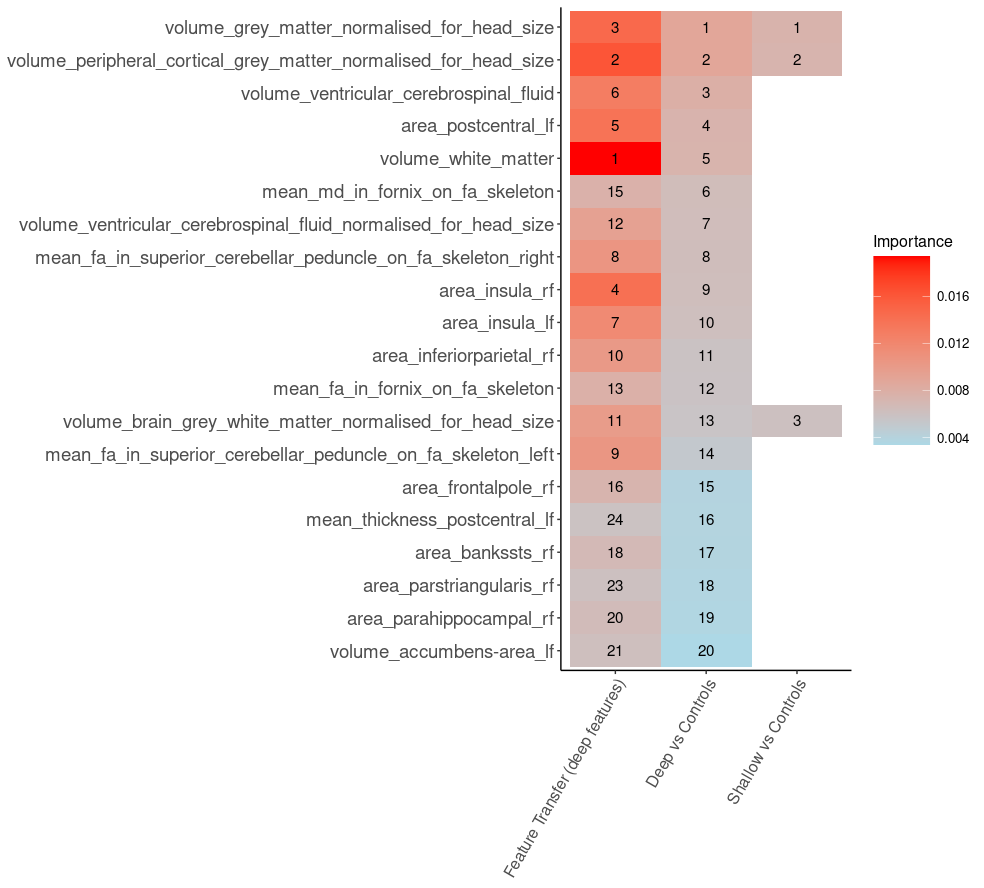

**Supplementary Figure S12 | Deep feature transfer framework and importance comparison across depression models.** This figure examines a feature transfer approach comparing three models: feature transfer (deep), deep depression vs controls, and shallow vs controls. The analysis focuses on top features identified through recursive feature elimination (RFE) in discriminating deep depression from controls in the training set. Feature importance was calculated using permutation-based methods in the Ranger R package, where importance scores reflect the decrease in model AUC when feature values are randomly permuted across out-of-bag samples. The heatmap visualizes feature rankings and relative importance (red indicating higher importance, blue lower) across all three models. Empty cells in the “Shallow vs Controls” column indicate features that were not selected during the initial feature selection process, which retained only the top 30% of features most predictive of distinguishing shallow depression from controls in the training set (70%). Volume-based measurements show high importance across models, with normalized gray matter and peripheral cortical gray matter volumes ranking consistently in the top positions. However, notable differences emerge in other features' rankings. For example, ventricular cerebrospinal fluid volume ranks 6th in the feature transfer model and 3rd in deep vs controls, but is absent from the shallow vs controls top rankings. Similarly, the right insular area ranks 7th and 10th in feature transfer and deep vs controls respectively, but does not appear among top features for shallow vs controls. The numerous blank spaces in the "Shallow vs Controls" column indicate limited overlap between features predictive of deep versus shallow depression, suggesting distinct neuroanatomical signatures between depression subtypes.
