## Supplementary Figures for "Mapping heterogeneity in the neuroanatomical correlates of depression": Supplementary Figure S13 _ Overlap in depression case definitions across phenotypes_.docx

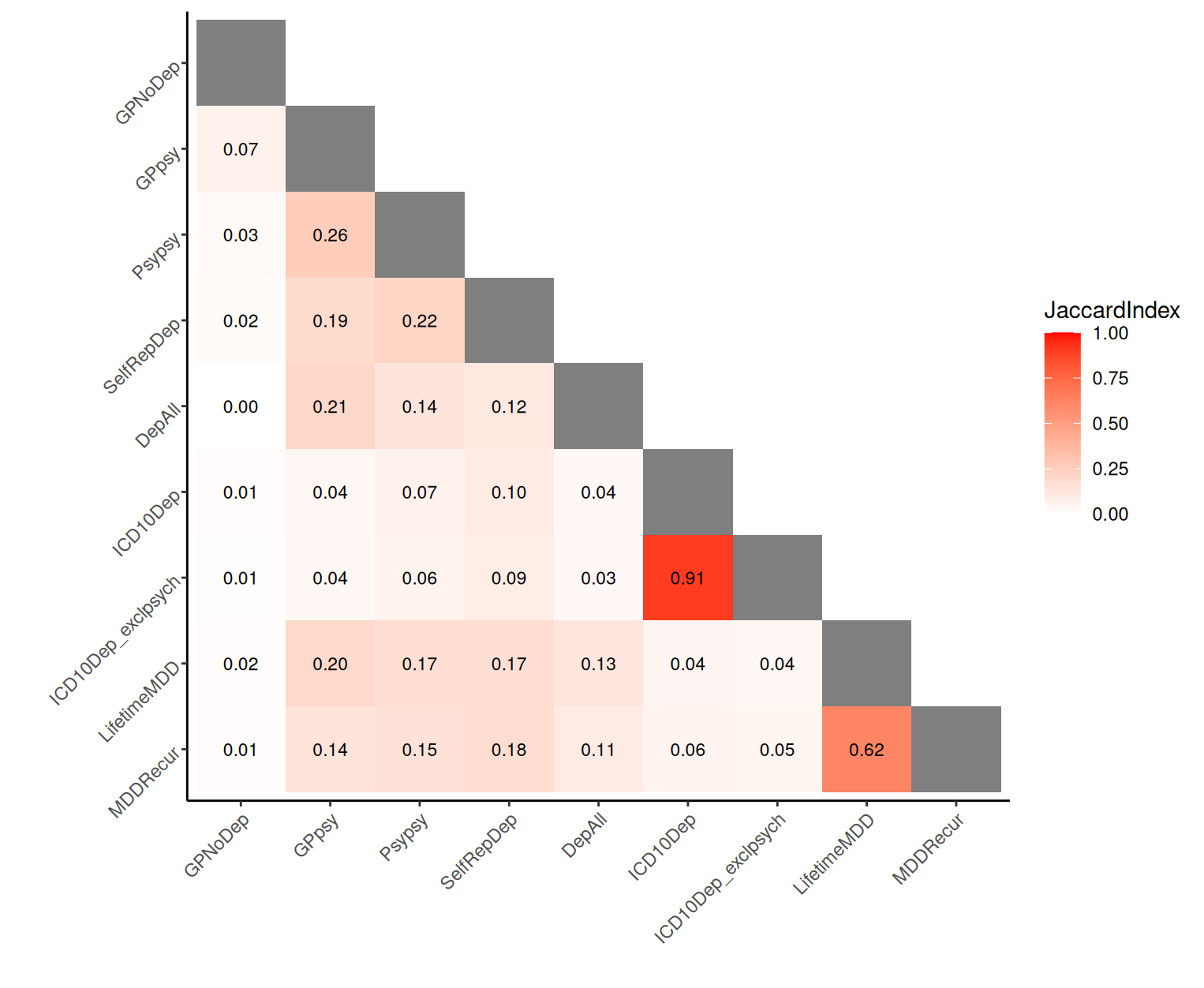

**Supplementary Figure S13 | Overlap in depression case definitions across phenotypes.**

The index ranges from 0 (no overlap) to 1 (complete overlap), providing a normalized measure of similarity calculated by dividing the size of the intersection (common elements) by the size of the union (total unique elements) between two sets of subjects for different depression definitions. These are displayed in a symmetric matrix where diagonal elements are 1 (self-comparison) and off-diagonal elements show the pairwise overlap between definitions. Full definitions and case/control counts across depression phenotypes are provided in Table 1.
